# Cost-effectiveness of a government rebate program for air cleaners in preventing asthma and related adverse health outcomes

**DOI:** 10.1101/2025.10.03.25337202

**Authors:** Spencer Lee, Amin Adibi, Amanda Giang, Chris Carlsten, Naman Paul, Emily Brigham, Kate M. Johnson

## Abstract

**Background:** Wildfire-derived fine particulate matter (PM_2.5_) is an increasing contributor to air pollution in British Columbia (BC), Canada, and is linked to asthma development and asthma-related adverse outcomes including exacerbations. Portable high-efficiency particulate air (HEPA) cleaners can reduce indoor PM_2.5_, but evidence on their long-term, population-level cost-effectiveness is limited.

**Methods:** We developed a monthly, time-varying Markov cohort model (2010–2035; healthcare payer perspective; 1.5% annual discounting; $50,000/QALY willingness-to-pay) for two BC cohorts (age 5 and age 25 at baseline) across 16 Health Service Delivery Areas (HSDAs). The model simulated monthly transitions between health states (well-controlled asthma, not well-controlled asthma, exacerbations, and death) over a 25-year time horizon (2010–2035). The target population was children age 5 and adults aged 25 (separate cohorts). We combined historical total PM_2.5_ (2010–2022) from provincial data sources with projected PM_2.5_ (2023–2035) combining anthropogenic emissions with wildfire-derived PM_2.5_. Wildfire PM_2.5_ projections were calculated by multiplying monthly PM_2.5_ averages from 2018–2022 by 0%, 5.5%, and 11% cumulative PM_2.5_ increase scenarios, informed by climate modeling. The base-case rebate was $150, and varied rebates between $50 to $200. We assumed continuous use with unit replacement every 5 years and filter replacement every 9 months. We calculated asthma incidence attributable to wildfire PM_2.5_ and incremental cost-effectiveness ratios (ICERs) across BC’s 16 Health Service Delivery Areas (HSDAs). We conducted the analysis from a health payer perspective with a 1.5% discount rate and $50,000/QALY willingness to pay threshold. Costs are expressed in 2024 CAD.

**Results:** From 2023–2036, wildfire-derived PM_2.5_ was associated with 13–14 incident asthma cases per 100,000 person-years annually across BC. Over 25 years, air cleaners prevented 444 moderate exacerbations, 55 emergency department visits, and 42 hospitalizations (combined cohorts), but the base-case program was not cost-effective in any HSDA (ICER range: $149,408–$154,749/QALY). A $50 rebate was cost-effective province-wide and $100 was cost-effective in three HSDAs. Results were most sensitive to concentration-response functions for PM2.5 (incidence, control, exacerbations) and to HEPA air cleaner and asthma care costs. Cost-effectiveness was most sensitive to air cleaner costs and the concentration-response function between PM_2.5_ and asthma incidence.

**Conclusions:** Wildfire-related PM_2.5_ contributes meaningfully to asthma incidence in BC. A universal $150 HEPA rebate program was not cost-effective for primary prevention under base-case assumptions, whereas lower rebates ($50 province-wide, $100 in three HSDAs) may offer better value. Future evaluations should co-benefits across multiple disease outcomes to better support policies to reduce the health impacts of increasing wildfires.

## Introduction

Asthma is a chronic disease characterized by airway inflammation and hyperresponsiveness. With an annual incidence of 600 cases per 100,000 person years and a prevalence of 12%^1^, asthma is one of the most prevalent chronic conditions in Canada. Environmental exposures, including air pollution, play a role in asthma development, symptom severity, and exacerbation risk^2–6^.

Ambient (outdoor) air pollution remains one of the largest contributors to global disease burden including asthma^7^. Ambient air pollution is comprised of gaseous (e.g., NO_2_, O_3_, SO_2_) and particulate components, with fine particulate matter (PM_2.5_) being of high concern due to its consistently demonstrated health effects and ability to penetrate deep into the lung. PM_2.5_ is emitted from anthropogenic activity (e.g. traffic, industry) and biomass burning (including wildfires)^8^. Nearly one-third of global asthma cases are attributable to PM ^9^, and exposure to PM_2.5_ is consistently associated with higher asthma incidence^2,5^, exacerbation risk, and healthcare utilization across the life course^10,11^.

Recent global analyses have highlighted the disproportionate and growing health burden of wildfire PM_2.5_, and in Canada wildfire smoke accounts for a substantial and growing proportion of annual PM_2.5_ exposure^8^. Wildfire activity has intensified over recent decades, driven by climate change, and is reversing decades of improving air quality in cities across North America through emission reduction measures and air quality policies^12^. British Columbia (BC) is uniquely vulnerable to deteriorating air quality due to climate change because of its regular exposure to wildfires^13^. Since the start of the 21^st^ century, BC has experienced a dramatic increase in wildfire activity with a nearly tenfold increase in area burned from 2017-2021 compared to late 20^th^ century averages, and four of the five worst wildfire years (2017, 2018, 2021, and 2023) occurring in the past decade alone^14^. This makes BC a critical setting for examining interventions aimed at reducing the impact of wildfire smoke on asthma and other respiratory outcomes.

Environmental health interventions, including climate mitigation strategies for wildfire PM_2.5,_ often face structural barriers: healthcare funding is typically allocated within disease areas, and the value of prevention strategies is not captured within short time frames, making them less attractive to decision makers with short-term budget cycles^15^. Yet, in population-based BC estimates, approximately 42% of outdoor PM_2.5_ infiltrated indoors^16^, where individuals spend an estimated 90% of their time^17,18^. Indoor PM_2.5_ is therefore an important, individually modifiable risk factor for asthma incidence and burden. Portable air cleaners with high-efficiency particulate air (HEPA) filters can substantially reduce indoor PM_2.5_ concentrations^19^, and are an opportunity for personalized intervention. However, mitigation strategies in indoor environments are inequitable and often inaccessible without public investment^20^.

Given the growing burden of wildfire PM_2.5_ exposure and the known health impacts of PM_2.5_ on asthma incidence and burden, quantifying the future health risks of PM_2.5_ and the related cost-effectiveness of climate mitigation strategies is critical for informing public health and environmental policy. We therefore aimed to: (1) estimate the future asthma burden attributable to wildfire-related PM_2.5_ and (2) evaluate the potential health and economic value of a portable air cleaner rebate program in BC.

## Methods

The findings of this study are reported in line with the Consolidated Health Economic Evaluation Reporting Standard (CHEERS) 2022 statement^21^. Our target population was the general BC population at age 5 years and 25 years (separate cohorts), to represent differences in natural history and economic burden of asthma among children and adults. Within these populations, we evaluated the cost-effectiveness of a government funded rebate program for portable HEPA air cleaners in preventing asthma and reducing asthma-related morbidity over a 25-year time horizon (2010-2035). The comparator strategy was no HEPA air cleaners. The time horizon included both a historical exposure period (2010-2022) and projected period (2023-2035) using future wildfire and anthropogenic PM_2.5_ estimates. We developed PM_2.5_ projections from 2023-2036 and used this longer-time span to estimate asthma incidence attributable to wildfire PM_2.5_ (Objective 1). However, the cost-effectiveness analysis (Objective 2) ended in 2035 to avoid purchasing a replacement air cleaner in the final year of the simulation with no opportunity for benefits to accrue over its 5-year lifespan. We used monthly time cycles to capture seasonal variation in air pollution.

Costs are presented in 2024 Canadian dollars with an annual discount rate of 1.5% applied to costs and effects, with scenario analysis of 0% and 3% discount rates, consistent with Canada’s Drug Agency guidelines^22^. The geographic unit of analysis was Health Service Delivery Areas (HSDAs), which are administrative units used for healthcare delivery, and the base case analysis was from the health care payer perspective. We used a willingness-to-pay (WTP) threshold of $50,000 per quality-adjusted life-year (QALY)^23^.

### Model development

We developed a Markov cohort model with health states for: (1) general BC population (non-asthma), (2) well-controlled asthma, (3) not well-controlled asthma (includes both partly controlled and uncontrolled asthma) defined using Global Initiative for Asthma criteria^24^, (4) moderate asthma exacerbation requiring oral systemic corticosteroids (OCS), (5) severe asthma exacerbation leading to emergency room (ER) visit or (6) hospitalization, and (7) death (**Figure 1**). We parameterized the model by using BC-specific estimates when available, followed by Canadian, North American, and global data sources.

**Figure 1.**
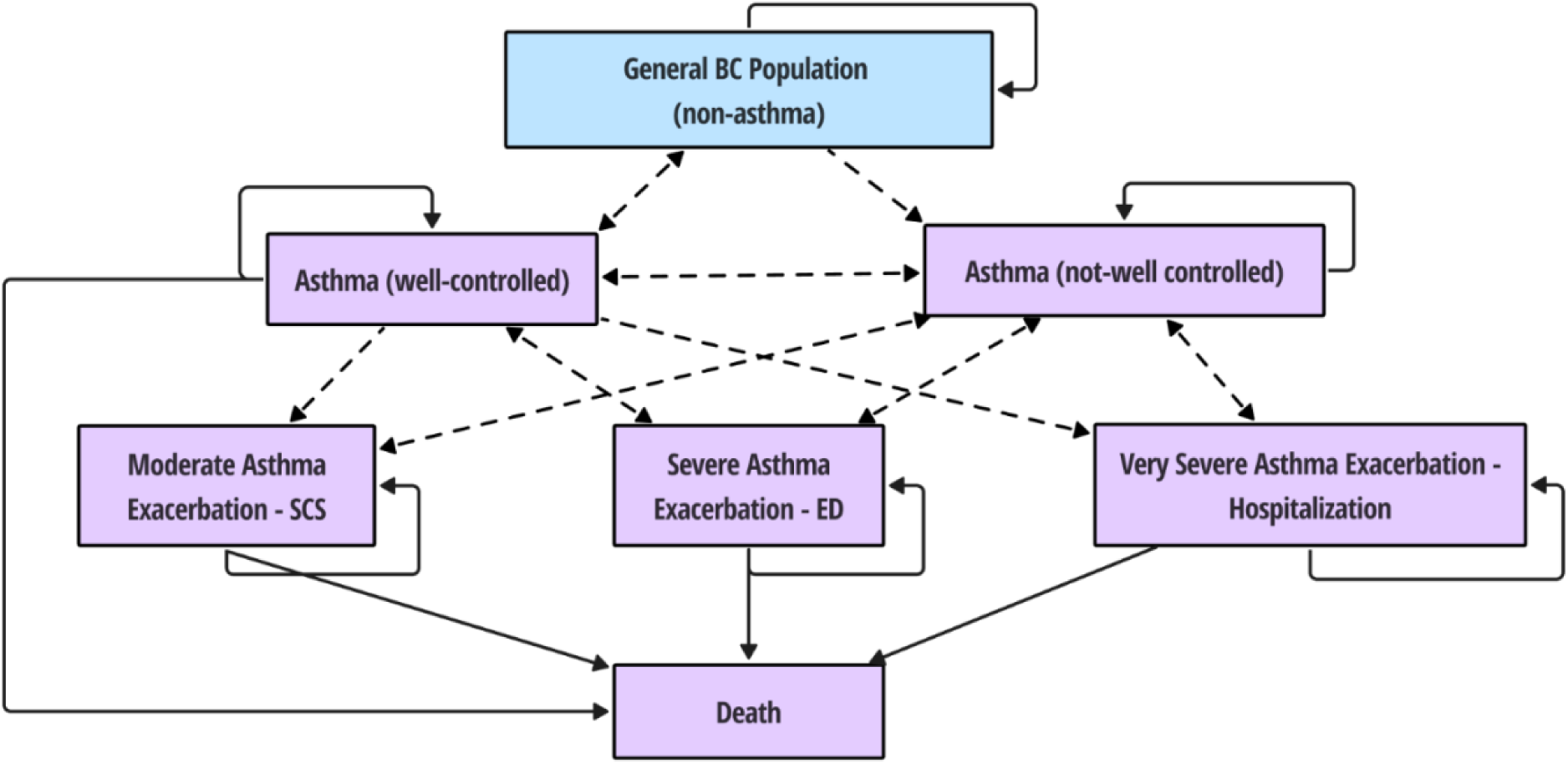
Markov model diagram (health states and health state transitions). OCS = Oral systemic corticosteroids; ED: emergency department. Dashed arrows indicate transition probabilities modified by exposure to PM_2.5._

### Asthma-related parameters

#### Asthma Incidence

Annual asthma incidence and prevalence rates by age group were derived from provincial data sources (**Appendix 2 and 3**)^25^. We averaged rates across three-year periods (2010–2012, 2013– 2015, and 2016–2019) and excluded data from 2020-2023 due to the impact of the COVID-19 pandemic on healthcare utilization and asthma diagnoses. The most recent estimates (2016–2019) were used for the projected period (2023–2036) in the model. The transition from well-controlled asthma back to non-asthma (BC general population) was predicted by a remission probability that was equal to asthma prevalence minus the sum of asthma incidence probability and asthma-related mortality probability. As a result, asthma prevalence remained stable within each three-year period and HSDA, assuming a constant PM_2.5_ concentration.

#### Asthma control

The proportion of individuals with asthma in the well-controlled asthma state at baseline and following an OCS exacerbation was 49% in children (≤17 years), informed by a United Kingdom (UK) study on school aged children^26^, and 56% in adults (≥18 years), informed by a *post-hoc* analysis of the Symbicort Given as Needed in Mild Asthma (SYGMA)-1 trial^27^. We calculated transition probabilities for well-controlled to not well-controlled asthma and vice versa using three-month transitions between control categories in the Economic Burden of Asthma (EBA) study, which was conducted among children and adults with mild-moderate asthma in BC, converted to monthly transition probabilities^28^.

#### Asthma exacerbations

Following previous economic evaluations in SYGMA-2, we assumed an average exacerbation length of 6 days for hospitalizations, 4 days for ED visits, and 2 days for OCS exacerbations^29^. The annual risk of moderate to severe exacerbations in children was derived from a US-based cohort of children with mild to moderate asthma^30^, and the as-needed budesonide-formoterol arm in the SYGMA-2 trial among adults, both converted into monthly transition probabilities^27^. We applied a risk ratio of 1.20 for the increased risk of exacerbations in children and adults with not well-controlled asthma, which was based on a large US commercial claims study^31^.

The probability of death following an asthma exacerbation of each severity was based on rates of in-hospital mortality in a UK administrative database analysis^32^; estimates were age-independent. Annual all-cause mortality was determined from age-specific life tables for BC, converted to a monthly probability^33^.

### Air pollution Exposure

Multiple air pollutant models informed PM_2.5_ exposure estimates over the historical (2010-2022) and projected (2023-2036) periods (**Figure 2**). For the historical period from 2010-2022, we used daily PM_2.5_ concentration estimates from the Canadian Optimized Statistical Smoke Model (CanOSSEM) at 5x5 km spatial resolution, aggregated at the monthly and HSDA level^34^. CanOSSEM is a random-forest machine learning model that estimates total ambient PM_2.5_ concentrations across populated regions in Canada and is optimized to capture biomass sources of PM_2.5_ including wildfire emissions. We estimated monthly wildfire-attributable PM_2.5_ from 2018-2022 for each HSDA by subtracting PM_2.5_ concentration from anthropogenic sources provided by the Regional Air Quality Deterministic Prediction System (RAQDPS) from PM_2.5_ concentrations from all sources (including wildfires) using the RAQDPS-FireWork (RAQDPS-FW) model^35^.

For 2023 to 2036, we used projections of PM_2.5_ concentrations with present-day (2019) climate conditions, but with future anthropogenic emissions under business-as-usual (BAU) scenario via the Global Environment Multiscale – Modelling Air CHemistry (GEM-MACH) model^36^. The BAU scenario reflects expected emissions trajectories in the absence of new major policy interventions and has been used in prior regulatory and health impact assessment conducted by the Government of Canada^36^. Our approach focuses on changes in anthropogenic and wildfire emissions under a scenario with limited climate action, but does not account for climate-driven changes in atmospheric chemistry or meteorological conditions that may also affect air quality. Further details are provided in **Appendix 1.**

**Figure 2.**
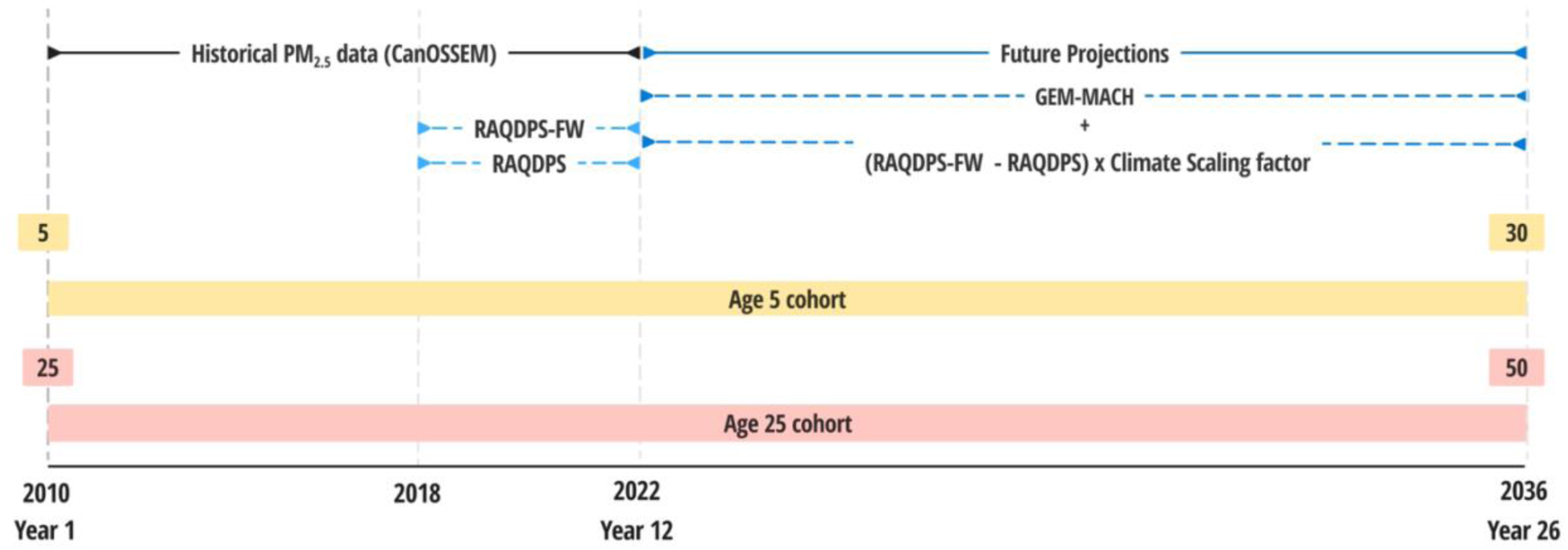
Data sources used to inform historical (2010-2022) and future (2023-2036) PM_2.5_ concentrations. CanOSSEM = Canadian Optimized Statistical Smoke Exposure Model, RAQDPS = Regional Air Quality Deterministic Prediction System; RAQDPS-FW = RAQDPS-FireWork; GEM-MACH = Global Environmental Multiscale – Modelling Air quality and CHemistry.

To account for increases in wildfire PM_2.5_ with time^12^, we applied a scaling factor (*S* in eq. 1) to monthly wildfire PM_2.5_ concentrations from 2023-2036 representing the following scenarios: no increase (0%), moderate increase (5.5%), and high increase (11%) in average wildfire PM_2.5_ concentrations from 2018-2022 to 2036 (**Table 1**). The high increase scenario was derived from an estimated baseline 49.2% increase in wildfire PM_2.5_ from 2001 to 2059 in the western United States, under the Representative Concentration Pathway (RCP) 8.5 scenario^37^, scaled proportionally to the 14-year projection period (2023–2036).

**Table 1.** Scenarios for increasing wildfire PM_2.5_ from 2023 to 2036.

| Future wildfire scenario | Annual increase in wildfire PM<sub>2.5</sub> | Total increase in wildfire PM<sub>2.5</sub> from 2023-2036 |
| --- | --- | --- |
| No increase | 0% | 0% |
| Moderate increase | 0.42% | 5.5% |
| High increase | 0.84% | 11% |

Therefore, total PM_2.5_ for each month (*i*) and HSDA (*j*) from 2023-2036, was calculated as shown in *equation 1*:

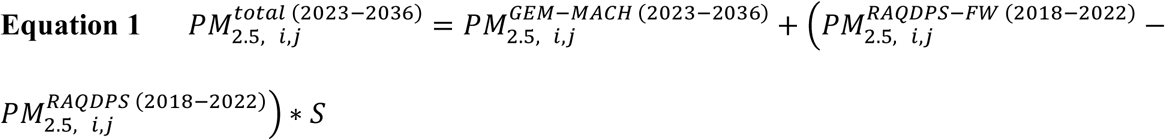

#### Impact of air pollution

We used concentration response functions (CRFs) to model the effect of increasing PM_2.5._ concentrations on transition probabilities for asthma incidence, worsened asthma control, and asthma exacerbations. For asthma incidence, the CRFs were derived from recent meta-analyses demonstrating a risk ratio (RR) of 1.16 per 5 μg/m^3^ increase in PM_2.5_ in children^5^, and a RR of 1.07 per 5 μg/m^3^ increase in PM_2.5_ in adults^2^. For worsened asthma control, we applied a RR of 1.04 per 10 μg/m^3^ increase in PM_2.5_ from a study of the association between salbutamol dispensations and daily PM_2.5_ in BC^11^. From the same study, we used a RR of 1.06 per 10 μg/m^3^ increase in PM_2.5_ for asthma-related outpatient visits, which was used as a proxy for OCS exacerbations^11^. The RR of asthma-related ED visits and hospitalizations was 1.07 and 1.06 per 10 μg/m^3^ increase in PM_2.5_, respectively^10^. RRs were applied to transition probabilities assuming a log-linear relationship, as shown in equation 2, where *p_1_* is the transition probability at a given PM_2.5_ concentration, *p_0_* is the baseline probability from the literature and assumed to occur at a PM_2.5_ of 0 μg/m^3^, RR is the relative risk estimate, and ΔPM_2.5_ is the PM_2.5_ concentration expressed in 5-10 µg/m^3^ increments, depending on the parameter.

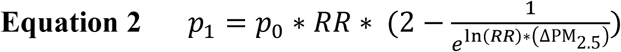

### Air pollution infiltration and air cleaner effectiveness

To estimate total monthly PM_2.5_ exposure we adjusted outdoor concentrations to reflect both infiltration into indoor environments and the amount of time individuals spend indoors. We applied an infiltration efficiency of 61% based on a study of households in BC^16^, and multiplied this by 89.1% or 88.2% indoors average time spent indoors among individuals aged <12 and ≥12 years, respectively, from a Canadian time use survey^17^. Total exposure was calculated as a weighted average of indoor and outdoor PM_2.5_ exposure. Indoor sources of PM_2.5_ (e.g., cooking, smoking, indoor residential wood burning) were not included in the model.

We modelled a government-funded rebate program for the purchase of portable HEPA air cleaners, assuming a base-case rebate of $150 applied at the point of sale. This amount is consistent with existing rebate programs in BC, such as FortisBC’s Connect Thermostat rebate^38^. We assumed a new air cleaner of the same model was purchased every 5 years (the average lifespan of the unit)^39^. Filter replacement costs were paid out-of-pocket by individuals, assuming a $30 purchase cost every nine months, with no rebate applied. We assumed an air cleaner rebate was provided to each person regardless of whether they were in a shared household, and that the air cleaners operated continuously with a filtration efficiency of 0.48 based on a recent pooled estimate of air cleaner effectiveness under wildfire smoke conditions^40^.

### Asthma-related costs and utilities

The direct costs of well-controlled and not well-controlled asthma for children and adults were derived from the EBA study, including only the costs of medications while outpatient visits as exacerbation costs were counted separately^28^. We calculated the direct costs of asthma exacerbations by combining healthcare resource use from the SYGMA-2 trial with Canadian unit costs^28,41^. We calculated indirect costs of exacerbations by multiplying the exacerbation length (in days) by the 2024 Canadian median weekly wage^42^.

We considered the indirect costs of well-controlled and not well-controlled asthma due to workplace absenteeism and presenteeism for both patients and caregivers. In the EBA study, adults reported 7.6 and 8.2 hours per week of productivity loss for well-controlled and not well-controlled asthma, respectively^28,41,42^. In children, we considered caregiver productivity loss from paid work of 36 hours per school year, stratified by control status using the lower (8 hours, well-controlled) and upper (36 hours, not well-controlled) interquartile values. We valued productivity loss for children using the same 2024 Canadian median weekly wage as adults.

General population utilities, which represent health-related quality of life where 0 represents death and 1 represents perfect health, were derived from a Canadian population-based survey using the EQ-5D-5L instrument, stratified by age groups (<18, 18–24, 25–34, 45–54 years)^43^. The utility of well-controlled asthma was 0.867 using the EQ-5D-5L reported in SYGMA-2. We converted an annual disutility of 0.2 for not well-controlled asthma from the 2011 US National Health and Wellness Survey into a monthly disutility. Exacerbation disutilities were based on a UK asthma cohort and applied during the cycle in which the exacerbation occurred, adjusted for exacerbation length^44^.

### Analysis of projected wildfire attributable PM_2.5_ on asthma incidence

To evaluate the impact of projected wildfire PM_2.5_ on future asthma incidence, we calculated the average annual incidence of asthma per 100,000 population in each HSDA during the projected period (2023-2036), under the no wildfire increase scenario. To estimate the average incidence of asthma over the 2023-2036 period, we calculated the total number of individuals transitioning into an asthma-related health state over one year (12 monthly model cycles) and divided by the total person-time at risk in that year. We reported annual incidence per 100,000 population.

### Sensitivity analysis

We conducted one-way deterministic sensitivity analysis (DSA) and probabilistic sensitivity analysis (PSA) with Monte Carlo simulation and 1,000 repetitions (**Appendix 7 & 8**). Parameter bounds for the DSA were set at ±20% of the base case value unless otherwise specified. In addition, we conducted the following scenario analyses: (1) societal perspective scenario incorporating indirect costs due to workplace productive loss, out of pocket costs of filter replacement, and electricity costs for running air cleaners; (2) discount rates of 0% and 3% (aligned with Canda’s Drug Agency guidelines^23^), (3) an alternative WTP threshold of $100,000 and $150,000 per QALY, (4) an alternative outdoor-to-indoor infiltration efficiency of 0.28, based on a large North American household study^45^, and (5) alternative rebate levels of $50, $100, and $200 (**Appendix 8-13**).

We performed the analysis using R (version 4.3.1) and *heemod* package (version 0.15.1)^46^.

**Table 2.** Model Parameters.

| Parameter | Base case | DSA | PSA | Source |
| --- | --- | --- | --- | --- |
| Fixed model inputs |  |  |  |  |
| Start year | 2010 | - | - |  |
| Childhood cohort age at start, yr | 5 | 5, 10 | - |  |
| Adult cohort age at start, yr | 25 | 20, 30 | - |  |
| Discounting (annually) | 1.5% | - | - |  |
| Air cleaner unit lifespan (years) | 5 | - | - |  |
| Air cleaner filter lifespan (months) | 9 | - | - |  |
| Baseline mortality | Varies by age (years) | - | - | Statistics Canada life tables<sup>33</sup> |
| Exposure inputs |  |  |  |  |
| Infiltration efficiency | 0.61 | ± 20% | Normal | Barn et al (2008)<sup>16</sup> |
| Filter effect | 0.48 | ± 20% | Beta | Huff et al. (2025)<sup>40</sup> |
| Proportion of time spent at home | 0.891(≤12 years)<br>0.903 (>12 years) | ± 20% | Beta | Matz et al (2014)<sup>17</sup> |
| Rates, Probabilities, and Risk |  |  |  |  |
| *Asthma control* |  |  |  |  |
| Proportion of well-controlled asthma at baseline | 0.506 (≤18 years)<br>0.560 (>18 years) | ± 20% | Beta | Kennedy et al (2024), Sadatsafavi et al (2015), O'Bryne et al (2021)<sup>26–28</sup> |
| Proportion of not-well controlled asthma at baseline | 0.494 (≤18 years)<br>0.440 (>18) | ± 20% | Beta |  |
| Probability of well-controlled asthma to not-well controlled asthma (monthly) | 0.244 | ± 20% | Beta | Sadatsafavi et al (2015)<sup>28</sup> |
| Probability of not well-controlled asthma to controlled asthma (monthly) | 0.140 | ± 20% | Beta |  |
| *Asthma exacerbations* |  |  |  |  |
| Relative risk of exacerbations in not well-controlled asthma (vs. well-controlled) | 1.20 | ± 20% | Log-normal | Pollack et al. (2022)<sup>31</sup> |
| Rate of OCS exacerbation (annually) | 0.10 (≤18 years)<br>0.090 (>18 years) | ± 20% | Beta | Adams et al (2001), Bateman et al (2018), Pollack et al (2022)<sup>30,31,47</sup> |
| Rate of ED visit exacerbation (annually) | 0.068 (≤18 years)<br>0.011 (>18 years) | ± 20% | Beta |  |
| Rate of hospitalization exacerbation (annually) | 0.019 (≤18 years)<br>0.009 (age>18) | ± 20% | Beta |  |
| Risk of death due to OCS exacerbation (per event) | 0.00027 | ± 20% | Beta | Watson et al (2007)<sup>32</sup> |
| Risk of death due to ED visit exacerbation (per event) | 0.001733 | ± 20% | Beta |  |
| Risk of death due to hospitalization exacerbation (per event) | 0.001801 | ± 20% | Beta |  |
| *Effects of PM<sub>2.5</sub>* |  |  |  |  |
| RR incident asthma (per 5 ug/m<sup>3</sup> PM<sub>2.5</sub>) | 1.16 (≤18 years)<br>1.07 (>18 years) | 1, 1.20 | Log-normal | Khreis et al (2017),<br>Lee et al (2024)<sup>2,5</sup> |
| RR for not well-controlled asthma (per 10 ug/m<sup>3</sup> PM<sub>2.5</sub>) | 1.04 | 1, 1.20 | Log-normal | Yao et al (2016)<sup>11</sup> |
| RR for OCS exacerbation (per 10 ug/m<sup>3</sup> PM<sub>2.5</sub>) | 1.06 | 1, 1.20 | Log-normal |  |
| RR for ED visit exacerbation (per 10 ug/m<sup>3</sup> PM<sub>2.5</sub>) | 1.07 | 1, 1.20 | Log-normal | Borchers et al (2019)<sup>10</sup> |
| RR for hospitalization exacerbation (per 10 ug/m<sup>3</sup> PM<sub>2.5</sub>) | 1.06 | 1, 1.20 | Log-normal | Borchers et al (2019)<sup>10</sup> |
| *Air cleaner costs* |  |  |  |  |
| Air cleaner discount (bulk) | 30% |  |  | Assumption |
| Air cleaner unit (after discount, every five years) | \$150 | ± 20% | Gamma | Assumption |
| Annual electricity cost for air cleaner (continuous use)* | \$10.08 | ± 20% | Gamma | |
| Filter replacement (every nine months) | \$30.00 | ± 20% | Gamma | |
| Asthma direct costs |  |  |  |  |
| Well-controlled asthma (monthly) | \$24.41 | ± 20% | Gamma | Sadatsafavi et al (2015)<sup>28</sup> |
| Not-well controlled asthma (monthly) | \$170.07 | ± 20% | Gamma | |
| OCS exacerbation (per event) | \$185.04 | ± 20% | Gamma | Sadatsafavi et al (2021)<sup>41</sup> |
| ED visit exacerbation (per event) | \$585.41 | ± 20% | Gamma | |
| Hospitalization exacerbation (per event) | \$11,211.59 | ± 20% | Gamma | |
| Asthma indirect costs |  |  |  |  |
| Well-controlled asthma (monthly)<sup>†</sup> | \$161.16 (≤18 years)<br>\$1,268.49 (>18 years) | ± 20% | Gamma | Sadatsafavi et al (2015) & Kennedy et al (2024)<sup>26,28</sup> |
| Not well-controlled asthma (monthly)<sup>†</sup> | \$886.37 (≤18 years)<br>\$1,364.34 (>18 years) | ± 20% | Gamma | Sadatsafavi et al (2015) & Kennedy et al (2024)<sup>26,28</sup> |
| OCS exacerbation<sup>‡</sup> | \$591.90 | ± 20% | Gamma | Sadatsafavi et al (2021)<sup>41</sup> |
| ED visit exacerbation<sup>‡</sup> | \$1,183.81 | ± 20% | Gamma | |
| Hospitalization exacerbation<sup>‡</sup> | \$1,775.71 | ± 20% | Gamma | |
| Utilities |  |  |  |  |
| General population | 0.95 (5-11 years)<br>0.89 (12-17 years) | ± 20% | Beta | Yan et al (2024)<sup>43</sup> |
|  | 0.86 (>17 years) |  |  |  |
| Disutility of well-controlled asthma (monthly) | 0.0042 (≤18 years)<br>0.013 (>18 years) | ± 20% | Beta | Lee et al (2024) & Lee et al (2018)<sup>48,49</sup> |
| Disutility of not well-controlled asthma (monthly) | 0.0067 (≤18 years)<br>0.017 (>18 years) | ± 20% | Beta |  |
| Disutility of OCS exacerbation<sup>‡</sup> | 0.0057 | ± 20% | Normal | Lloyd et al (2007)<sup>44</sup> |
| Disutility of ED visit exacerbation<sup>‡</sup> | 0.0075 | ± 20% | Normal |  |
| Disutility of hospitalization exacerbation<sup>‡</sup> | 0.0092 | ± 20% | Normal |  |

Parameters are for both child and adult cohorts, unless otherwise stated.
\*Electricity costs for air cleaner operation were estimated assuming continuous use at maximum fan speed for a typical air cleaner unit (10 W) for 24 hours per day over one year, at a unit electricity rate of \$0.1265 per kWh<sup>50</sup>.
†Indirect costs were calculated by multiplying productivity time losses with 2024 average weekly earnings in Canada of \$1,285.91 per week<sup>42</sup>.
‡Disutility values were scaled to reflect the duration of each exacerbation; 2 days for OCS exacerbation, 4 days for ER exacerbation, and 6 days for hospitalization.

## Results

There was substantial interannual and regional variability in total PM_2.5_ concentrations from 2010 to 2022. Average monthly total PM_2.5_ levels ranged from 4.2 to 97.6 µg/m³ across HSDAs, with the highest concentrations in the Interior and Northern regions and the highest recorded concentration in July (**Appendix 5**). Wildfire PM_2.5_ activity contributed to an average of 42% of total PM_2.5_ from 2018-2022 across all HSDAs monthly. Highest wildfire contributions were observed in Northern Interior (52%), Kootenay Boundary (47%), and Thompson Cariboo Shuswap (45%). Lowest wildfire contributions were observed in South Vancouver Island (29%), Central Vancouver Island (36%), North Vancouver Island (38%). There was relatively high agreement between total PM_2.5_ estimates from CanOSSEM and RAQDPS from 2018–2022, with RAQDPS overestimating peak PM_2.5_ during wildfires, and underestimating baseline PM_2.5_ exposure (See **Appendix 1 & B7**). Across HSDAs, the total cumulative increase in wildfire-attributable PM_2.5_ concentration from 2023-2036 was 98 µg/m³ under the moderate wildfire increase scenario and 123 µg/m³ under the high scenario, based on the sum of monthly concentration increases over the 14-year period (**Appendix 6**). By 2036, the cumulative wildfire-attributable PM_2.5_ exposure increased between 7 and 9 µg/m^3^ per month under the moderate and high wildfire scenarios, respectively, compared to the no-increase scenario (**Appendix 6**). Under the counterfactual scenario of anthropogenic emissions only, projected monthly PM_2.5_ concentrations from 2023– 2036 were 0.9–20.1 µg/m^3^ across all HSDAs, with limited year-to-year variation. (**Appendix 5**).

### Projected asthma incidence attributable to wildfire PM_2.5_

Across BC from 2023 to 2036, there were 13 new asthma cases attributable to wildfire PM_2_ per 100,000 children in the age 5 cohort, with a range across HSDAs of 8.7 to 23.6 per 100,000 people annually (**Table 3**). There were 14 new cases per 100,000 adults in the age 25 cohort, with a range of 9.8 to 14.6 per 100,000 people annually (**Table 3**). Asthma incidence attributable to wildfire PM_2.5_ varied by HSDA; we observed higher incidence rates in Interior and Northern regions of British Columbia, particularly Thompson Cariboo Shuswap (12.6 per 100,000 children annually, 13.0 per 100,000 adults annually), East Kootenay (12.9 per 100,000 children annually, 12.5 per 100,000 adults annually), and Norther Interior (13.2 per 100,000 children annually, 12.1 per 100,000 adults annually) reflecting greater projected wildfire smoke exposure.

**Table 3.** Projected asthma incidence from 2023-2036, with and without wildfire PM_2.5_ exposure.

| HSDA | Annual Incidence Rate (per 100,000) from 2023-2036 |  |  |  |  |  |
| --- | --- | --- | --- | --- | --- | --- |
|  | Age 5 (to 18) cohort |  |  | Age 25 (to 38) cohort |  |  |
|  | Including Wildfire PM<sub>2.5</sub> | No wildfire PM<sub>2.5</sub> counterfactual | Attributable to wildfire PM<sub>2.5</sub> | Including Wildfire PM<sub>2.5</sub> | No wildfire PM<sub>2.5</sub> counterfactual | Attributable to wildfire PM<sub>2.5</sub> |
| Central Vancouver Island | 483 | 469 | 14.6 | 454 | 441 | 13.0 |
| East Kootenay | 462 | 449 | 12.8 | 444 | 432 | 12.5 |
| Fraser East | 468 | 454 | 13.3 | 468 | 454 | 13.4 |
| Fraser North | 492 | 478 | 13.8 | 507 | 493 | 14.3 |
| Fraser South | 471 | 458 | 13.3 | 605 | 588 | 17.2 |
| Kootenay Boundary | 381 | 370 | 10.6 | 506 | 491 | 14.3 |
| North Shore/Coast Garibaldi | 453 | 440 | 12.6 | 441 | 429 | 12.4 |
| North Vancouver Island | 428 | 416 | 12.0 | 407 | 395 | 11.7 |
| Northeast | 455 | 444 | 12.6 | 319 | 310 | 9.0 |
| Northern Interior | 400 | 389 | 13.2 | 422 | 410 | 12.1 |
| Northwest | 388 | 377 | 10.8 | 474 | 460 | 13.5 |
| Okanagan | 510 | 495 | 14.3 | 501 | 487 | 14.3 |
| Richmond | 445 | 433 | 12.4 | 539 | 524 | 15.0 |
| South Vancouver Island | 484 | 472 | 13.6 | 470 | 457 | 13.4 |
| Thompson Cariboo Shuswap | 449 | 437 | 12.6 | 455 | 442 | 13.0 |
| Vancouver | 423 | 411 | 11.8 | 474 | 460 | 13.2 |
| **BC overall\*** | **462 ± 37.4** | **449 ± 36.4** | **13.0 ± 1.1** | **494 ± 61.9** | **480 ± 60.2** | **14.0 ± 1.7** |

HSDA: Health Service Delivery Area
\*Weighted BC average based on population size of each HSDA in 2023 (Appendix 4).

### Cost-effectiveness of a HEPA Air Cleaner Rebate Program

In the base-case analysis the intervention was not cost-effective in any Health Service Delivery Area (HSDA) at a WTP threshold of $50,000 per QALY. There was little variation in ICERs across the province, which ranged from $149,757 to $154,749 per QALY in adults and $150,166 to $155,815 per QALY in children (**Table 4**). In BC, in both child (5 to 30) and adult (25 to 50) cohorts combined, air cleaners resulted in 444 fewer OCS exacerbations, 55 fewer ED visits, and 42 fewer hospitalizations over 25 years. The highest reductions in OCS exacerbations due to air cleaners were observed in Fraser South (41.3 fewer) among children, and Vancouver (48.4 fewer) among adults. The lowest reductions in OCS exacerbations occurred in East Kootenay (2.9 fewer) among children and Kootenay Boundary (2.7 fewer) in adults. This variation is largely attributable to differences in population size among HSDAs. Similarly, air cleaners prevented the most ED exacerbations in Vancouver (6.0 fewer) among adults and (1.7 fewer) among children over 25 years. Air cleaners prevented 4.3 hospitalizations in Vancouver and 3.6 hospitalizations in Fraser South among adults from 2010-2035.

In one-way deterministic sensitivity analysis, results were most sensitive to the relative risk for the association between PM_2.5_ and asthma incidence, not well-controlled asthma, and exacerbations of all severities (**Appendix 7**). The ICERs were also highly influenced by the cost of air cleaners, the direct costs of asthma control and exacerbations. The probability of cost-effectiveness in PSA was <5% in all HSDAs at the $50,000 and $100,000 WTP thresholds but increased to approximately 50% at the $150,000 WTP threshold (**Appendix 8**).

In scenario analysis, the ICERs for government rebates for air cleaners were higher when indirect costs and patient-incurred costs for air filter replacement and electricity were included in the societal perspective, with a BC wide ICER of $445,907/QALY and $418,306/QALY for the child and adult cohorts, respectively (**Appendix 9**). Alternative discount rates of 0% and 3% per annum had a smaller impact on the results and did not result in cost-effectiveness in any HSDA (**Appendix 10 & 11**).

When the rebate of air cleaners was reduced to $50, the intervention was cost-effective across all HSDAs, with an ICER of $31,176/QALY for the child cohorts and $33,431/QALY for the adult cohorts (**Appendix 12**). At the $100 rebate level, the intervention was cost-effective in a three HSDAs across both child and adult cohorts, including Kootenay Boundary ($43,414/QALY child; $40,700/QALY adult), Northeast ($47,098/QALY child, $44,404/QALY adult), and Thompson-Cariboo Shuswap ($43,332/QALY child, $46,291/QALY adult) (**Appendix 12**). At the $200 rebate level, the intervention was not cost effective in any HSDA (**Appendix 12**).

**Table 4.**
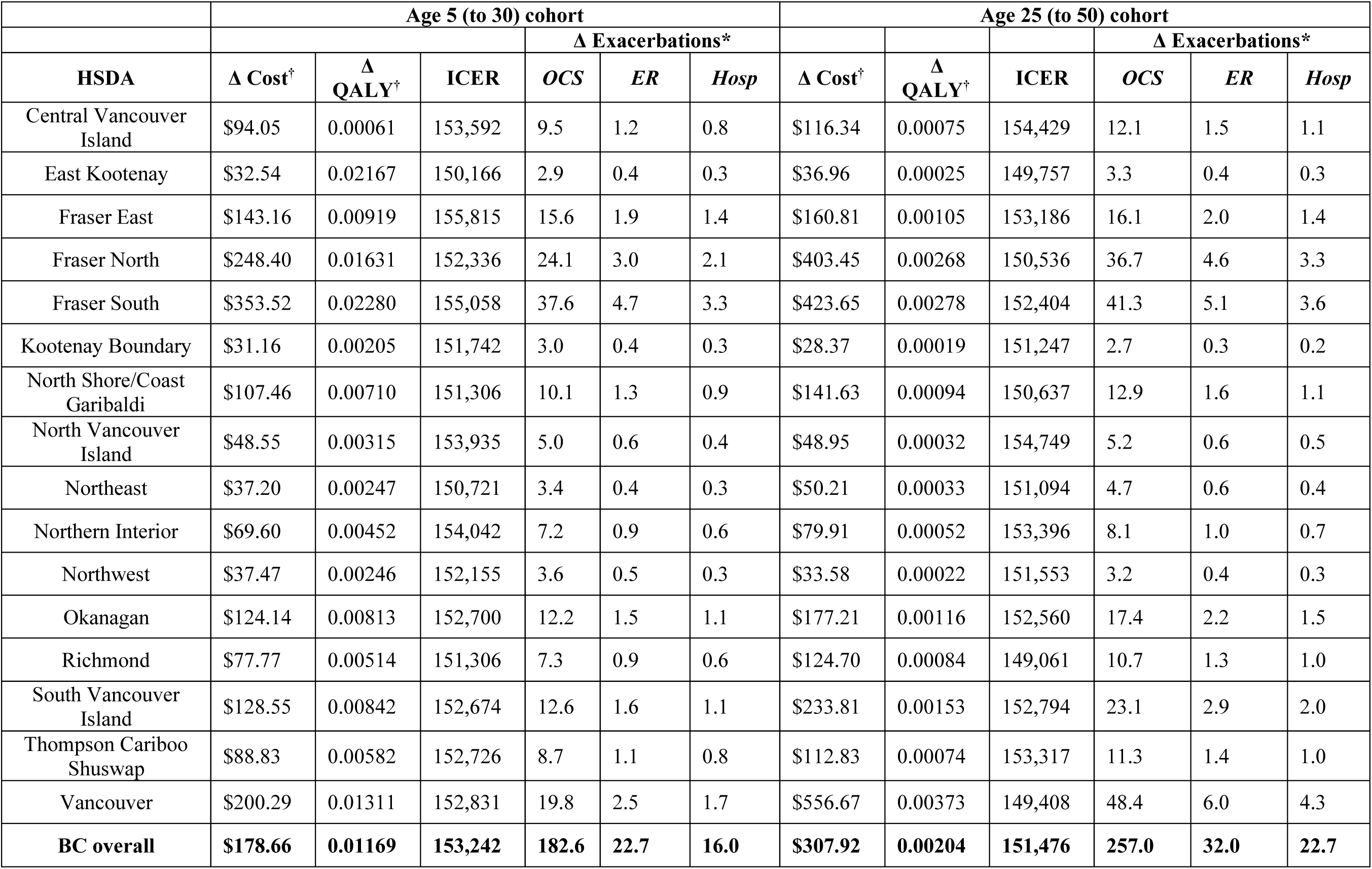
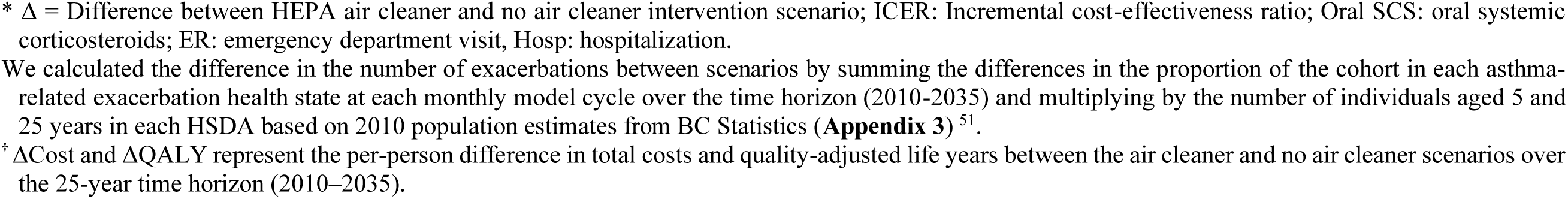
Base-cost effectiveness analysis from healthcare payer perspective from 2010-2035 (25-year time horizon). Costs and Quality Adjusted Life-years (QALYs) are reported per-patient.

|  | Age 5 (to 30) cohort |  |  |  |  |  | Age 25 (to 50) cohort |  |  |  |  |  |
| --- | --- | --- | --- | --- | --- | --- | --- | --- | --- | --- | --- | --- |
|  |  |  |  | Δ Exacerbations* |  |  |  |  |  | Δ Exacerbations* |  |  |
| HSDA | Δ Cost<sup>†</sup> | Δ QALY<sup>†</sup> | ICER | OCS | ER | Hosp | Δ Cost<sup>†</sup> | Δ QALY<sup>†</sup> | ICER | OCS | ER | Hosp |
| Central Vancouver Island | \$94.05 | 0.00061 | 153,592 | 9.5 | 1.2 | 0.8 | \$116.34 | 0.00075 | 154,429 | 12.1 | 1.5 | 1.1 |
| East Kootenay | \$32.54 | 0.02167 | 150,166 | 2.9 | 0.4 | 0.3 | \$36.96 | 0.00025 | 149,757 | 3.3 | 0.4 | 0.3 |
| Fraser East | \$143.16 | 0.00919 | 155,815 | 15.6 | 1.9 | 1.4 | \$160.81 | 0.00105 | 153,186 | 16.1 | 2.0 | 1.4 |
| Fraser North | \$248.40 | 0.01631 | 152,336 | 24.1 | 3.0 | 2.1 | \$403.45 | 0.00268 | 150,536 | 36.7 | 4.6 | 3.3 |
| Fraser South | \$353.52 | 0.02280 | 155,058 | 37.6 | 4.7 | 3.3 | \$423.65 | 0.00278 | 152,404 | 41.3 | 5.1 | 3.6 |
| Kootenay Boundary | \$31.16 | 0.00205 | 151,742 | 3.0 | 0.4 | 0.3 | \$28.37 | 0.00019 | 151,247 | 2.7 | 0.3 | 0.2 |
| North Shore/Coast Garibaldi | \$107.46 | 0.00710 | 151,306 | 10.1 | 1.3 | 0.9 | \$141.63 | 0.00094 | 150,637 | 12.9 | 1.6 | 1.1 |
| North Vancouver Island | \$48.55 | 0.00315 | 153,935 | 5.0 | 0.6 | 0.4 | \$48.95 | 0.00032 | 154,749 | 5.2 | 0.6 | 0.5 |
| Northeast | \$37.20 | 0.00247 | 150,721 | 3.4 | 0.4 | 0.3 | \$50.21 | 0.00033 | 151,094 | 4.7 | 0.6 | 0.4 |
| Northern Interior | \$69.60 | 0.00452 | 154,042 | 7.2 | 0.9 | 0.6 | \$79.91 | 0.00052 | 153,396 | 8.1 | 1.0 | 0.7 |
| Northwest | \$37.47 | 0.00246 | 152,155 | 3.6 | 0.5 | 0.3 | \$33.58 | 0.00022 | 151,553 | 3.2 | 0.4 | 0.3 |
| Okanagan | \$124.14 | 0.00813 | 152,700 | 12.2 | 1.5 | 1.1 | \$177.21 | 0.00116 | 152,560 | 17.4 | 2.2 | 1.5 |
| Richmond | \$77.77 | 0.00514 | 151,306 | 7.3 | 0.9 | 0.6 | \$124.70 | 0.00084 | 149,061 | 10.7 | 1.3 | 1.0 |
| South Vancouver Island | \$128.55 | 0.00842 | 152,674 | 12.6 | 1.6 | 1.1 | \$233.81 | 0.00153 | 152,794 | 23.1 | 2.9 | 2.0 |
| Thompson Cariboo Shuswap | \$88.83 | 0.00582 | 152,726 | 8.7 | 1.1 | 0.8 | \$112.83 | 0.00074 | 153,317 | 11.3 | 1.4 | 1.0 |
| Vancouver | \$200.29 | 0.01311 | 152,831 | 19.8 | 2.5 | 1.7 | \$556.67 | 0.00373 | 149,408 | 48.4 | 6.0 | 4.3 |
| **BC overall** | **\$178.66** | **0.01169** | **153,242** | **182.6** | **22.7** | **16.0** | **\$307.92** | **0.00204** | **151,476** | **257.0** | **32.0** | **22.7** |

\* $\Delta$ = Difference between HEPA air cleaner and no air cleaner intervention scenario; ICER: Incremental cost-effectiveness ratio; Oral SCS: oral systemic corticosteroids; ER: emergency department visit, Hosp: hospitalization.
We calculated the difference in the number of exacerbations between scenarios by summing the differences in the proportion of the cohort in each asthma-related exacerbation health state at each monthly model cycle over the time horizon (2010-2035) and multiplying by the number of individuals aged 5 and 25 years in each HSDA based on 2010 population estimates from BC Statistics (**Appendix 3**)<sup>51</sup>.
<sup>†</sup> $\Delta$ Cost and $\Delta$ QALY represent the per-person difference in total costs and quality-adjusted life years between the air cleaner and no air cleaner scenarios over the 25-year time horizon (2010–2035).

## Discussion

Asthma poses a significant burden on population health and healthcare systems in BC, and PM_2.5_ exposure is a major contributor to this burden. In our study, wildfire PM_2.5_ contributed substantially to annual PM_2.5_ concentrations in BC, resulting in new asthma cases and asthma-related morbidity and mortality. Wildfire PM_2.5_ caused 2-3% of incident asthma cases annually in BC, for a total attributable burden of 13–14 out of 605 total cases per 100,000 person-years. Although this is lower than the estimated 13%-33% of asthma cases worldwide attributable to PM_2.5_ from traffic-related air pollution and indoor exposures^9,52,53^, wildfire PM_2.5_ remains a major risk factor for asthma. The difference may result from our differing methodologies or may reflect the episodic and regionally variable nature of wildfire smoke compared to persistent, year-round exposure from traffic and indoor sources^7,54^. As wildfires become more frequent and intense due to climate change, the associated burden on asthma incidence is likely to increase substantially^12,17^.

Implementation of a universal air cleaner rebate program reduced both asthma incidence and morbidity, but $150 government rebate of air cleaners was not cost-effective in any HSDA at a WTP threshold of $50,000/QALY. From the societal perspective, the ICERs were substantially higher when the patient costs of electricity use, and air cleaner filter replacement were included. However, a reduced rebate of $50 in all HSDAs and $100 of the air cleaner purchase cost was cost-effective in three HSDAs (Kootenay Boundary, Northeast, and Thompson Cariboo Shuswap) from the healthcare perspective (which did not include electricity or filter replacement costs), suggesting lower rebate levels may offer good value for money.

Previous studies have demonstrated the use of HEPA air filters results in reductions in indoor PM_2.5_. However, evidence of improved respiratory symptoms among children and adults with asthma or other pre-existing respiratory conditions is mixed^55–58^. In a previous cost-effectiveness analysis (CEA) using a similar model over a retrospective time horizon, $150 rebate for air cleaners was cost-effective (at a $50,000/QALY WTP threshold) among adults with prevalent asthma, but only in those HSDAs most affected by wildfire smoke. The ICERs in this study ranged from $40,509 to $89,206 per QALY depending on region, which is 39-73% lower than the ICERs determined in our study^59^. However, in our study, air cleaners were distributed to the general population of children and adults in BC, while the previous study only examined adults with prevalent asthma. Whereas the previous model used retrospective PM_2.5_ over a five-year time horizon, our model incorporated future PM_2.5_ projections across a 25-year time horizon and explored asthma health outcomes in both pediatric and adult populations. Our study represents an important advancement in the evaluation of air cleaners for primary prevention of asthma.

As noted, the health gains from preventing asthma in a small proportion of the population were insufficient to offset the costs of $150 rebates for air cleaners, however a partial rebate of $50 and $100 was cost effective in regions with high PM_2.5_ exposure. The model was highly sensitive to the association between PM_2.5_ and asthma health outcomes, including asthma incidence, worsened asthma control, and risk of exacerbations (OCS, ER visit, and hospitalization). Wildfire PM_2.5_ has been linked to asthma morbidity and related healthcare utilization, in some cases with a greater effect size compared to PM_2.5_ from other sources^7,8,10,53,60^. Wildfire PM_2.5_ may have higher toxicity compared to other sources of PM_2.5_, but this is likely to vary based on source and particle aging^61^. As a result, the potential unique contribution of wildfire-specific PM_2.5_ to asthma incidence has not been quantified. While our model CRF estimates were based on all-source PM_2.5_, if PM_2.5_ from wildfire smoke does indeed pose greater respiratory toxicity in equivalent doses due to its unique chemical composition, then the CRFs in our current model may underestimate the potential health benefits of air cleaners^4^.

Long-term PM_2.5_ exposure is linked to asthma incidence across the lifespan, as supported by meta-analyses from large, population-based cohorts^2,5^. Despite modelling childhood and adult cohorts separately using age-specific concentration-response functions (CRFs), incidence rates, asthma progression parameters, and economic burden, we found that ICERs were similar across both age groups. Although CRFs differed slightly by age, we modeled identical exposure to PM_2.5_, resulting in similar reductions in asthma incidence and exacerbations in both cohorts. This may reflect the broad population-level exposure reduction achieved by the intervention. While children are often more susceptible to environmental exposures, our findings suggest that portable HEPA air cleaners confer similar modeled health gains per person across the life course ^6,61^. Despite these benefits, $150 air cleaner rebates were not cost-effective in either cohort under base case assumptions, emphasizing the need to explore more targeted or high-risk subgroups, such as individuals with pre-existing asthma, young children, older adults, or those living in regions with persistently high PM_2.5_ exposure, where the benefits of air cleaner use may outweigh the costs^62^. Alternatively, a reduced air cleaner rebate ($50 and $100) is likely to be cost-effective in most regions with high wildfire smoke exposure.

Our model has several limitations. For parsimony we assumed idealized use of air cleaners (e.g., continuous operation, timely filter replacement), which may overestimate real-world effectiveness^63^. Air cleaners are also known to decrease in effectiveness and efficiency in reducing PM_2.5_ over time between filter replacements, which we did not account for^64^. While we did not specify a particular air cleaner model, the rebate amount modeled in this study would likely apply to smaller, entry-level portable units^16^. These devices are typically designed for use in small rooms (∼18 m²), which may be insufficient to effectively reduce PM_2.5_ exposure in larger or open living spaces, potentially leading to reduced real-world effectiveness. Individuals are also assigned to an air cleaner, rather than modelling air cleaner use at the household level, which may overestimate intervention costs. Our model did not account for indoor sources of PM_2.5_. Since indoor sources of PM_2.5_ would be present in both intervention and compactor scenarios, their exclusion likely had minimal impact on relative effect estimates, but they may influence absolute exposure levels and differences in effect along the concentration-response function. Our projections of wildfire PM_2.5_ involved simplified scaling assumptions, which were highly conservative and assumed a marginal increase over average PM_2.5_ concentrations from 2018-2022. These estimates are likely to change depending on future climate and meteorological conditions^37^. We have not captured nonlinear changes in wildfire activity due to extreme drought, fuel conditions, and other compounding effects. While our projections incorporate future anthropogenic emissions under a business-as-usual scenario, they do not account for other factors which may influence non-wildfire PM_2.5_ such as climate-driven changes in meteorology or future policy interventions^65–67^.

Our study is one of the first to evaluate the cost-effectiveness of air cleaner rebates as a primary prevention strategy for asthma. We incorporated the most recent effect estimates for age-specific relationships between PM_2.5_ concentrations and asthma incidence and morbidity, enhancing clinical and epidemiological validity^2,5,10^ and reflecting the potential for unique impacts across the life course^68^. While our analysis focuses on asthma burden alone, PM_2.5_ exposure is related to multiple disease states, including cardiovascular disease, COPD, lung cancer, diabetes, adverse birth outcomes, and neurocognitive decline^69–75^, and air cleaners may therefore have multiple co-benefits when deployed in a general population. Clinical trials have assessed air cleaners for short-term disease management, demonstrating improved indoor air quality, reduced asthma symptoms, and decreased medication use^57,76,77^. However, studies have largely focused on small populations over limited time periods. In contrast, evaluating air cleaners for primary prevention requires large populations and long-term follow-up to fully assess impact, which is not feasible in trial settings. Cost-effectiveness modeling is particularly well-suited for investigating the long-term health and economic impacts of such strategies. Our modeling framework represents a valuable tool in evaluating climate adaptation strategies aimed air reducing the health impacts from worsening air pollution^78^.

## Conclusion

Wildfire-derived PM_2.5_ is projected to contribute 13–14 new cases of asthma per 100,000 population annually across British Columbia over a 14-year projection period (2023-2036). A universal $150 rebate program for portable HEPA air cleaners was not cost-effective for preventing asthma and related adverse outcomes, however a rebate of $50 was fully cost-effective and $100 was cost-effective in regions with high wildfire PM_2.5_ exposure. Future evaluations should explore co-benefits for other pollution-related diseases, dynamic climate-exposure modeling frameworks, adherence/usage scenarios, and varying rebate levels to guide public health decision-making.

## Data Availability

The model code and supporting files are publicly available on GitHub (https://github.com/resplab/hepa_ce_v3).

## References

1. Canadian Chronic Disease Surveillance System (CCDSS). Canadian Chronic Disease Surveillance System (CCDSS). August 2022. Accessed March 7, 2023. https://health-infobase.canada.ca/ccdss/data-tool/Index

2. Lee S, Tian D, He R, et al. Ambient air pollution exposure and adult asthma incidence: a systematic review and meta-analysis. The Lancet Planetary Health. 2024;8(12):e1065–e1078. doi:10.1016/S2542-5196(24)00279-1

3. Anenberg SC, Mohegh A, Goldberg DL, et al. Long-term trends in urban NO2 concentrations and associated paediatric asthma incidence: estimates from global datasets. The Lancet Planetary Health. 2022;6(1):e49–e58. doi:10.1016/S2542-5196(21)00255-2

4. Aguilera R, Corringham T, Gershunov A, Benmarhnia T. Wildfire smoke impacts respiratory health more than fine particles from other sources: observational evidence from Southern California. Nat Commun. 2021;12(1):1493. doi:10.1038/s41467-021-21708-0

5. Khreis H, Kelly C, Tate J, Parslow R, Lucas K, Nieuwenhuijsen M. Exposure to traffic-related air pollution and risk of development of childhood asthma: A systematic review and meta-analysis. Environment International. 2017;100:1–31. doi:10.1016/j.envint.2016.11.012

6. Thurston GD, Balmes JR, Garcia E, et al. Outdoor Air Pollution and New-Onset Airway Disease. An Official American Thoracic Society Workshop Report. Ann Am Thorac Soc. 2020;17(4):387–398. doi:10.1513/AnnalsATS.202001-046ST

7. Brauer M, Roth GA, Aravkin AY, et al. Global burden and strength of evidence for 88 risk factors in 204 countries and 811 subnational locations, 1990–2021: a systematic analysis for the Global Burden of Disease Study 2021. The Lancet. 2024;403(10440):2162–2203. doi:10.1016/S0140-6736(24)00933-4

8. Meng J, Martin RV, Li C, et al. Source Contributions to Ambient Fine Particulate Matter for Canada. Environ Sci Technol. 2019;53(17):10269–10278. doi:10.1021/acs.est.9b02461

9. Ni R, Su H, Burnett RT, Guo Y, Cheng Y. Long-term exposure to PM2.5 has significant adverse effects on childhood and adult asthma: A global meta-analysis and health impact assessment. One Earth. 2024;7(11):1953–1969. doi:10.1016/j.oneear.2024.09.022

10. Borchers Arriagada N, Horsley JA, Palmer AJ, Morgan GG, Tham R, Johnston FH. Association between fire smoke fine particulate matter and asthma-related outcomes: Systematic review and meta-analysis. Environmental Research. 2019;179:108777. doi:10.1016/j.envres.2019.108777

11. Yao J, Eyamie J, Henderson SB. Evaluation of a spatially resolved forest fire smoke model for population-based epidemiologic exposure assessment. J Expo Sci Environ Epidemiol. 2016;26(3):233–240. doi:10.1038/jes.2014.67

12. Burke M, Childs ML, De La Cuesta B, et al. The contribution of wildfire to PM2.5 trends in the USA. Nature. 2023;622(7984):761–766. doi:10.1038/s41586-023-06522-6

13. Carlsten C, Brauer M, Camp PG, Nesbitt L, Turner J. British Columbia, Canada, as a bellwether for climate-driven respiratory and allergic disorders. Journal of Allergy and Clinical Immunology. 2023;152(5):1087–1089. doi:10.1016/j.jaci.2023.09.018

14. Parisien MA, Barber QE, Bourbonnais ML, et al. Abrupt, climate-induced increase in wildfires in British Columbia since the mid-2000s. Commun Earth Environ. 2023;4(1):309. doi:10.1038/s43247-023-00977-1

15. Nugent R, Bertram MY, Jan S, et al. Investing in non-communicable disease prevention and management to advance the Sustainable Development Goals. The Lancet. 2018;391(10134):2029–2035. doi:10.1016/S0140-6736(18)30667-6

16. Barn P, Larson T, Noullett M, Kennedy S, Copes R, Brauer M. Infiltration of forest fire and residential wood smoke: an evaluation of air cleaner effectiveness. J Expo Sci Environ Epidemiol. 2008;18(5):503–511. doi:10.1038/sj.jes.7500640

17. Matz CJ, Stieb DM, Davis K, et al. Effects of Age, Season, Gender and Urban-Rural Status on Time-Activity: Canadian Human Activity Pattern Survey 2 (CHAPS 2). Int J Environ Res Public Health. 2014;11(2):2108–2124. doi:10.3390/ijerph110202108

18. Klepeis NE, Nelson WC, Ott WR, et al. The National Human Activity Pattern Survey (NHAPS): a resource for assessing exposure to environmental pollutants. J Expo Sci Environ Epidemiol. 2001;11(3):231–252. doi:10.1038/sj.jea.7500165

19. Carlsten C, Salvi S, Wong GWK, Chung KF. Personal strategies to minimise effects of air pollution on respiratory health: advice for providers, patients and the public. European Respiratory Journal. Published online January 1, 2020. doi:10.1183/13993003.02056-2019

20. Han D, Guo Y, Wang J, Zhao B. Global disparities in indoor wildfire-PM2.5 exposure and mitigation costs. Science Advances. 2025;11(20):eads4360. doi:10.1126/sciadv.ads4360

21. Husereau D, Drummond M, Augustovski F, et al. Consolidated Health Economic Evaluation Reporting Standards 2022 (CHEERS 2022) statement: updated reporting guidance for health economic evaluations. BMC Medicine. 2022;20(1):23. doi:10.1186/s12916-021-02204-0

22. Guidelines for the Economic Evaluation of Health Technologies: Canada (4th Edition).

23. Guidelines for the Economic Evaluation of Health Technologies: Canada | CDA-AMC. Accessed November 11, 2024. https://www.cda-amc.ca/guidelines-economic-evaluation-health-technologies-canada-0

24. Boulet LP, Reddel HK, Bateman E, Pedersen S, FitzGerald JM, O’Byrne PM. The Global Initiative for Asthma (GINA): 25 years later. Eur Respir J. 2019;54(2):1900598. doi:10.1183/13993003.00598-2019

25. British Columbia Chronic Disease Registries (BCCDR) Case Definitions. British Columbia (BC) Ministry of Health Accessed February 28, 2023. http://www.bccdc.ca/resource-gallery/Documents/Chronic-Disease-Dashboard/asthma.pdf

26. Kennedy CT, Scotland GS, Cotton S, Turner SW. Direct and indirect costs of paediatric asthma in the UK: a cost analysis. Arch Dis Child. Published online May 27, 2024:archdischild-2023-326306. doi:10.1136/archdischild-2023-326306

27. O’Byrne PM, FitzGerald JM, Bateman ED, et al. Effect of a single day of increased as-needed budesonide–formoterol use on short-term risk of severe exacerbations in patients with mild asthma: a post-hoc analysis of the SYGMA 1 study. The Lancet Respiratory Medicine. 2021;9(2):149–158. doi:10.1016/S2213-2600(20)30416-1

28. Sadatsafavi M, McTaggart-Cowan H, Chen W, Mark FitzGerald J, Economic Burden of Asthma (EBA) Study Group. Quality of Life and Asthma Symptom Control: Room for Improvement in Care and Measurement. Value Health. 2015;18(8):1043–1049. doi:10.1016/j.jval.2015.07.008

29. Ho JK, Shaker M, Greenhawt M, et al. Cost-effectiveness of budesonide-formoterol vs inhaled epinephrine in US adults with mild asthma. Annals of Allergy, Asthma & Immunology. 2024;132(2):229–239.e3. doi:10.1016/j.anai.2023.10.024

30. Adams RJ, Fuhlbrigge A, Finkelstein JA, et al. Impact of Inhaled Antiinflammatory Therapy on Hospitalization and Emergency Department Visits for Children With Asthma. Pediatrics. 2001;107(4):706–711. doi:10.1542/peds.107.4.706

31. Pollack M, Gandhi H, Tkacz J, Lanz M, Lugogo N, Gilbert I. The use of short-acting bronchodilators and cost burden of asthma across Global Initiative for Asthma-based severity levels: Insights from a large US commercial and managed Medicaid population. J Manag Care Spec Pharm. 2022;28(8):881–891. doi:10.18553/jmcp.2022.21498

32. Watson L, Turk F, James P, Holgate ST. Factors associated with mortality after an asthma admission: a national United Kingdom database analysis. Respir Med. 2007;101(8):1659–1664. doi:10.1016/j.rmed.2007.03.006

33. Government of Canada SC. Life expectancy and other elements of the complete life table, single-year estimates, Canada, all provinces except Prince Edward Island. January 24, 2022. Accessed April 25, 2024. https://www150.statcan.gc.ca/t1/tbl1/en/tv.action?pid=1310083701

34. Paul N, Yao J, McLean KE, Stieb DM, Henderson SB. The Canadian Optimized Statistical Smoke Exposure Model (CanOSSEM): A machine learning approach to estimate national daily fine particulate matter (PM2.5) exposure. Science of The Total Environment. 2022;850:157956. doi:10.1016/j.scitotenv.2022.157956

35. Environment Climate Change Canada S. The Regional Air Quality Deterministic Prediction System (RAQDPS) Version 25.0.0 of the Meteorological Service of Canada (MSC): Technical Specifications Document.

36. Chen J, Pendlebury D, Gravel S, et al. Development and Current Status of the GEM-MACH-Global Modelling System at the Environment and Climate Change Canada. In: Mensink C, Gong W, Hakami A, eds. Air Pollution Modeling and Its Application XXVI. Springer International Publishing; 2020:107–112. doi:10.1007/978-3-030-22055-6_18

37. Liu Y, Liu Y, Fu J, et al. Projection of future wildfire emissions in western USA under climate change: contributions from changes in wildfire, fuel loading and fuel moisture. Int J Wildland Fire. 2021;31(1):1–13. doi:10.1071/WF20190

38. Connected thermostat rebates | FortisBC. FortisBC - Energy solutions for every customer. Accessed July 28, 2025. https://www.fortisbc.com/rebates/home/connected-thermostat-rebates

39. Blue Pure 411i Max | Air purifier for up to 526 ft^2^. Blueair. Accessed April 16, 2025. https://www.blueair.com/en-ca/products/blue-pure-411i-max

40. 40. Huff RD, Traynor RL, Carmargo K, et al. Rapid Review: What Effect Does Indoor Air Filtration and Air Cleaning Have on Concentrations of Pollutants and Human Health Endpoints during Combustion-Derived Air Pollution Episodes? The National Collaborating Centre for Environmental Health and The National Collaborating Centre for Methods and Tools https://ncceh.ca/resources/evidence-reviews/indoor-air-filtration-during-wildfires-impacts-air-quality-and-health#h2-0

41. Sadatsafavi M, FitzGerald JM, O’Byrne PM, et al. The cost-effectiveness of as-needed budesonide-formoterol versus low-dose inhaled corticosteroid maintenance therapy in patients with mild asthma in Canada. *Allergy*, Asthma & Clinical Immunology. 2021;17(1):108. doi:10.1186/s13223-021-00610-w

42. Government of Canada SC. Average weekly earnings, average hourly wage rate and average usual weekly hours by union status, annual. November 26, 2013. Accessed April 16, 2025. https://www150.statcan.gc.ca/t1/tbl1/en/tv.action?pid=1410013401

43. Yan J, Xie S, Johnson JA, et al. Canada population norms for the EQ-5D-5L. Eur J Health Econ. 2024;25(1):147–155. doi:10.1007/s10198-023-01570-1

44. Lloyd A, Price D, Brown R. The impact of asthma exacerbations on health-related quality of life in moderate to severe asthma patients in the UK. Prim Care Respir J. 2007;16(1):22–27. doi:10.3132/pcrj.2007.00002

45. Lunderberg DM, Liang Y, Singer BC, Apte JS, Nazaroff WW, Goldstein AH. Assessing residential PM _2.5_ concentrations and infiltration factors with high spatiotemporal resolution using crowdsourced sensors. Proc Natl Acad Sci USA. 2023;120(50). doi:10.1073/pnas.2308832120

46. Filipović-Pierucci A, Zarca K, Durand-Zaleski I. Markov Models for Health Economic Evaluations: The R Package heemod. *arXiv*. Preprint posted online April 25, 2017. doi:10.48550/arXiv.1702.03252

47. Bateman ED, Reddel HK, O’Byrne PM, et al. As-Needed Budesonide–Formoterol versus Maintenance Budesonide in Mild Asthma. New England Journal of Medicine. 2018;378(20):1877–1887. doi:10.1056/NEJMoa1715275

48. Lee LK, Obi E, Paknis B, Kavati A, Chipps B. Asthma control and disease burden in patients with asthma and allergic comorbidities. Journal of Asthma. 2018;55(2):208–219. doi:10.1080/02770903.2017.1316394

49. Lee TY, Petkau J, Johnson KM, et al. Development and Validation of an Asthma Policy Model for Canada: Lifetime Exposures and Asthma outcomes Projection (LEAP). medRxiv. Preprint posted online March 13, 2024:2024.03.11.24304122. doi:10.1101/2024.03.11.24304122

50. Appliance cost calculator. Accessed June 6, 2025. https://www.bchydro.com/powersmart/residential/tools-and-calculators/cost-calculator.html

51. BC Population Estimates & Projections. Accessed April 16, 2025. https://bcstats.shinyapps.io/popApp/

52. Anenberg SC, Mohegh A, Goldberg DL, et al. Long-term trends in urban NO2 concentrations and associated paediatric asthma incidence: estimates from global datasets. The Lancet Planetary Health. 2022;6(1):e49–e58. doi:10.1016/S2542-5196(21)00255-2

53. Anenberg SC, Henze DK, Tinney V, et al. Estimates of the Global Burden of Ambient PM2.5, Ozone, and NO2 on Asthma Incidence and Emergency Room Visits. Environmental Health Perspectives. 126(10):107004. doi:10.1289/EHP3766

54. Bell ML, Dominici F, Ebisu K, Zeger SL, Samet JM. Spatial and Temporal Variation in PM2.5 Chemical Composition in the United States for Health Effects Studies. Environmental Health Perspectives. 2007;115(7):989–995. doi:10.1289/ehp.9621

55. Ulziikhuu B, Gombojav E, Banzrai C, et al. Portable HEPA Filter Air Cleaner Use during Pregnancy and Children’s Cognitive Performance at Four Years of Age: The UGAAR Randomized Controlled Trial. Environmental Health Perspectives. 2022;130(6):067006. doi:10.1289/EHP10302

56. Weichenthal S, Mallach G, Kulka R, et al. A randomized double-blind crossover study of indoor air filtration and acute changes in cardiorespiratory health in a First Nations community. Indoor Air. 2013;23(3):175–184. doi:10.1111/ina.12019

57. Muanprasong S, Aqilah S, Hermayurisca F, Taneepanichskul N. Effectiveness of Asthma Home Management Manual and Low-Cost Air Filter on Quality of Life Among Asthma Adults: A 3-Arm Randomized Controlled Trial. J Multidiscip Healthc. 2024;17:2613–2622. doi:10.2147/JMDH.S397388

58. Park HJ, Lee HY, Suh CH, et al. The Effect of Particulate Matter Reduction by Indoor Air Filter Use on Respiratory Symptoms and Lung Function: A Systematic Review and Meta-analysis. Allergy Asthma Immunol Res. 2021;13(5):719–732. doi:10.4168/aair.2021.13.5.719

59. Adibi A, Barn P, Shellington EM, Harvard S, Johnson KM, Carlsten C. High-Efficiency Particulate Air Filters for Preventing Wildfire-related Asthma Complications: A Cost-Effectiveness Study. Am J Respir Crit Care Med. 2024;209(2):175–184. doi:10.1164/rccm.202307-1205OC

60. Rice MB, Henderson SB, Lambert AA, et al. Respiratory Impacts of Wildland Fire Smoke: Future Challenges and Policy Opportunities. An Official American Thoracic Society Workshop Report. Annals ATS. 2021;18(6):921–930. doi:10.1513/AnnalsATS.202102-148ST

61. Rice MB, Henderson SB, Lambert AA, et al. Respiratory Impacts of Wildland Fire Smoke: Future Challenges and Policy Opportunities. An Official American Thoracic Society Workshop Report. Annals ATS. 2021;18(6):921–930. doi:10.1513/AnnalsATS.202102-148ST

62. Dennin LR, Nock D, Muller NZ, Akindele M, Adams PJ. Socially vulnerable communities face disproportionate exposure and susceptibility to U.S. wildfire and prescribed burn smoke. Commun Earth Environ. 2025;6(1):1–19. doi:10.1038/s43247-025-02100-y

63. Lorizio W, Woo H, McCormack MC, et al. Patterns and Predictors of Air Cleaner Adherence Among Adults with COPD. Chronic Obstr Pulm Dis. 9(3):366–376. doi:10.15326/jcopdf.2022.0309

64. Jiang J, Liu J, Wang C, et al. Exploring the long-term performance of air purifiers in removing particulate matter and formaldehyde across different residential environments. Environmental Research. 2024;263:120194. doi:10.1016/j.envres.2024.120194

65. Bélanger D, Séguin J, Canada, eds. Human Health in a Changing Climate: A Canadian Assessment of Vulnerabilities and Adaptive Capacity. Health Canada; 2008.

66. CLIMATE CHANGE AND SOCIAL VULNERABILITY IN THE UNITED STATES - A focus on Six Impacts. doi:10.1163/9789004322714_cclc_2021-0166-513

67. Jacob DJ, Winner DA. Effect of climate change on air quality. Atmospheric Environment. 2009;43(1):51–63. doi:10.1016/j.atmosenv.2008.09.051

68. Trivedi M, Denton E. Asthma in Children and Adults—What Are the Differences and What Can They Tell us About Asthma? Front Pediatr. 2019;7:256. doi:10.3389/fped.2019.00256

69. Wang W, Mu S, Yan W, Ke N, Cheng H, Ding R. Prenatal PM2.5 exposure increases the risk of adverse pregnancy outcomes: evidence from meta-analysis of cohort studies. Environ Sci Pollut Res. 2023;30(48):106145–106197. doi:10.1007/s11356-023-29700-5

70. Hayes RB, Lim C, Zhang Y, et al. PM2.5 air pollution and cause-specific cardiovascular disease mortality. Int J Epidemiol. 2020;49(1):25–35. doi:10.1093/ije/dyz114

71. Alexeeff SE, Deosaransingh K, Van Den Eeden S, Schwartz J, Liao NS, Sidney S. Association of Long-term Exposure to Particulate Air Pollution With Cardiovascular Events in California. JAMA Network Open. 2023;6(2):e230561. doi:10.1001/jamanetworkopen.2023.0561

72. Yang X, Zhang T, Zhang Y, Chen H, Sang S. Global burden of COPD attributable to ambient PM2.5 in 204 countries and territories, 1990 to 2019: A systematic analysis for the Global Burden of Disease Study 2019. Science of The Total Environment. 2021;796:148819. doi:10.1016/j.scitotenv.2021.148819

73. Zhang T, Mao W, Gao J, et al. The effects of PM2.5 on lung cancer-related mortality in different regions and races: A systematic review and meta-analysis of cohort studies. Air Qual Atmos Health. 2022;15(9):1523–1532. doi:10.1007/s11869-022-01193-0

74. Ren Z, Yuan J, Luo Y, Wang J, Li Y. Association of air pollution and fine particulate matter (PM2.5) exposure with gestational diabetes: a systematic review and meta-analysis. Ann Transl Med. 2023;11(1):23. doi:10.21037/atm-22-6306

75. Thompson R, Smith RB, Karim YB, et al. Air pollution and human cognition: A systematic review and meta-analysis. Science of The Total Environment. 2023;859:160234. doi:10.1016/j.scitotenv.2022.160234

76. Drieling RL, Sampson PD, Krenz JE, et al. Randomized trial of a portable HEPA air cleaner intervention to reduce asthma morbidity among Latino children in an agricultural community. Environ Health. 2022;21(1):1. doi:10.1186/s12940-021-00816-w

77. Hansel NN, Putcha N, Woo H, et al. Randomized Clinical Trial of Air Cleaners to Improve Indoor Air Quality and Chronic Obstructive Pulmonary Disease Health: Results of the CLEAN AIR Study. Am J Respir Crit Care Med. 2022;205(4):421–430. doi:10.1164/rccm.202103-0604OC

78. Martins FP, Paschoalotto MAC, Closs J, Bukowski M, Veras MM. The Double Burden: Climate Change Challenges for Health Systems. Environ Health Insights. 2024;18:11786302241298789. doi:10.1177/11786302241298789

79. Whaley CH, Galarneau E, Makar PA, et al. GEM-MACH-PAH (rev2488): a new high-resolution chemical transport model for North American polycyclic aromatic hydrocarbons and benzene. Geoscientific Model Development. 2018;11(7):2609–2632. doi:10.5194/gmd-11-2609-2018

