## Supplemental Data for "Cost-effectiveness of a government rebate program for air cleaners in preventing asthma and related adverse health outcomes"

Spencer Lee, Amin Adibi, Amanda Giang, Chris Carlsten, Naman Paul, Emily Brigham, Kate M. Johnson

**Appendix 1.** Air pollution model sources.

**Historical $PM_{2.5}$**

*Canadian Optimized Statistical Smoke Exposure Model (CanOSSEM)*

The Canadian Optimized Statistical Smoke Exposure Model (CanOSSEM) is a machine learning model that estimates daily outdoor $PM_{2.5}$ concentrations at a 5×5 km spatial resolution across populated regions of Canada for 2010 to 2022[34]. CanOSSEM integrates ground-based measurements with remote-sensed smoke plume data, wildfire intensity, meteorological factors, and satellite aerosol optical depth observations to reconstruct past levels of air pollution, particularly due to biomass smoke. Because it is historically calibrated and validated against National Air Pollution Surveillance (NAPS) ground monitoring stations, CanOSSEM is currently considered the benchmark for measuring historical total $PM_{2.5}$ in Canada. In regions without NAPS stations, CanOSSEM relies on satellite observations and spatial predictors to estimate $PM_{2.5}$ concentrations. Its robust validations make it the most reliable dataset for historical $PM_{2.5}$ exposure assessment in Canada. However, potential biases remain, including residual misclassification from reliance on satellite proxies, uncertainties in smoke plume modeling, and limited representation of local-scale variability. We applied spatial and temporal transformations to convert the 5×5 km gridded CanOSSEM $PM_{2.5}$ outputs to monthly $PM_{2.5}$ concentrations aggregated by Health Service Delivery Area (HSDA), using R (v4.3.1) and the sf (v1.0.15), bcmaps (v2.2.0), and dplyr (v1.1.4) packages.

*Regional Air Quality Deterministic Prediction System (RAQDPS)*

The Regional Air Quality Deterministic Prediction System (RAQDPS) is a short-term air quality forecasting model developed by Environment and Climate Change Canada (ECCC)[35]. Since 2009, it has been used operationally to produce 3-day forecasts for pollutants such as $PM_{2.5}$, ozone ($O_3$), nitrogen dioxide ($NO_2$), and sulfur dioxide ($SO_2$) based on meteorological inputs, emissions inventories, and chemical transport modeling. These forecasts are used to inform the Air Quality Health Index (AQHI). RAQDPS is a forecast-oriented model and does not incorporate direct air pollutant measurements (e.g., $PM_{2.5}$, $NO_2$). Instead, it applies post-processing bias corrections using observed air quality data from ground-based monitoring networks like the National Air Pollution Surveillance (NAPS) program.

Although RAQDPS is primarily a short-term forecasting system, it can be adapted to inform future $PM_{2.5}$ projections. In this study, we used outputs from two configurations: RAQDPS, which estimates total $PM_{2.5}$ excluding wildfire emissions, and RAQDPS-FW, which includes wildfire-attributable $PM_{2.5}$. By comparing these outputs, we were able to isolate and quantify the wildfire-specific contribution to total $PM_{2.5}$ concentrations across British Columbia under various future wildfire activity scenarios.

*Comparison of CanOSSEM and RAQDPS*

While CanOSSEM and RAQDPS differ in their objectives and modeling approaches, CanOSSEM reconstructs historical $PM_{2.5}$ concentrations using machine learning, whereas RAQDPS forecasts future levels using emissions inventories and chemical transport models, they offer complementary insights for exposure assessment. Despite these differences, the models showed moderate-to-strong agreement in our analysis, with a Pearson correlation coefficient of 0.72, and normalized mean bias (NMB) of 19.6% for total $PM_{2.5}$ across overlapping years (2018-2022). This supports the validity of combining outputs from both models to estimate historical and projected $PM_{2.5}$ exposure across British Columbia.

*Projected anthropogenic $PM_{2.5}$ - GEM-Modelling Air-quality and Chemistry (GEM-MACH)*

GEM-MACH predicts future $PM_{2.5}$ (among other air pollutants including $NO_2$, $O_3$, and $SO_2$) levels from human (anthropogenic) sources using a chemical transport model that accounts for emissions from industry, transportation, and residential activities. GEM-MACH has been operationally used by ECCC since 2009 for air quality forecasting and undergoes routine validation against ground-based air quality measurements, including NAPS data[79]. However, GEM-MACH does not directly assimilate real-time $PM_{2.5}$ observations into its forecasts. Instead, its performance is evaluated post-run by comparing predicted $PM_{2.5}$ concentrations against observed values from NAPS stations. GEM-MACH $PM_{2.5}$ outputs are available for the years 2023, 2028, 2033, and 2036. To generate annual $PM_{2.5}$ estimates for intervening years, we applied linear interpolation across each 5-year interval.

**Appendix 2.** Annual incidence rate of asthma in British Columbia by Health Service Delivery Area and age group (1-19, 20-34, 35-49, and 50-64) from 2010 to 2019. Data sourced from the British Columbia Centre for Disease Control (BCCDC) Chronic Disease Dashboard[25].

| | | Incidence rate per 100,000 people (age group) | | | |
|---|---|---|---|---|---|
| **Health Service Delivery Area** | **Year** | **1-19** | **20-34** | **35-49** | **50-64** |
| Central Vancouver Island | 2010 | 1255.3 (1151.4–1366.0) | 553.0 (478.4–635.8) | 536.7 (470.7–609.4) | 539.9 (482.0–602.8) |
| | 2011 | 1135.3 (1035.9–1241.7) | 368.8 (308.3–437.6) | 352.6 (298.7–413.3) | 433.7 (382.3–490.1) |
| | 2012 | 980.4 (887.4–1080.4) | 447.8 (380.5–523.6) | 511.9 (445.6–585.3) | 445.4 (393.3–502.5) |
| | 2013 | 1146.7 (1045.5–1255.1) | 377.9 (316.2–448.1) | 464.3 (400.4–535.4) | 441.8 (389.9–498.7) |
| | 2014 | 1111.3 (1011.4–1218.4) | 377.7 (316.0–447.9) | 470.0 (405.2–542.2) | 511.5 (455.7–572.2) |
| | 2015 | 1104.3 (1005.4–1210.2) | 394.4 (331.8–465.4) | 509.9 (442.3–584.9) | 508.5 (453.2–568.7) |
| | 2016 | 1102.9 (1005.5–1207.2) | 354.7 (296.4–421.2) | 531.0 (462.9–606.4) | 512.8 (457.7–572.8) |
| | 2017 | 938.2 (849.5–1033.7) | 446.0 (380.9–519.0) | 513.1 (447.0–586.3) | 564.7 (506.8–627.5) |
| | 2018 | 962.6 (873.7–1058.2) | 412.5 (350.5–482.3) | 511.2 (445.9–583.4) | 437.1 (386.2–493.0) |
| | 2019 | 1021.7 (930.6–1119.2) | 483.7 (416.7–558.4) | 547.5 (480.4–621.3) | 576.2 (517.1–640.1) |
| East Kootenay | 2010 | 1025.7 (870.6–1200.5) | 467.1 (355.6–602.5) | 570.1 (454.0–706.7) | 461.0 (365.6–573.8) |
| | 2011 | 906.5 (759.5–1073.6) | 442.9 (333.7–576.5) | 530.5 (417.3–665.0) | 523.8 (422.3–642.4) |
| | 2012 | 869.3 (724.1–1035.0) | 467.9 (354.4–606.2) | 458.2 (352.1–586.3) | 541.7 (438.3–662.2) |
| | 2013 | 723.3 (591.0–876.4) | 432.5 (323.0–567.2) | 519.2 (404.8–656.0) | 365.3 (281.3–466.4) |
| | 2014 | 813.2 (671.9–975.3) | 474.4 (359.3–614.6) | 469.9 (361.1–601.2) | 411.1 (321.7–517.7) |
| | 2015 | 839.0 (696.2–1002.5) | 517.6 (396.9–663.6) | 535.2 (418.7–673.9) | 474.7 (378.1–588.5) |
| | 2016 | 992.3 (836.8–1168.2) | 343.0 (246.2–465.4) | 512.7 (399.7–647.8) | 437.4 (344.6–547.5) |
| | 2017 | 680.2 (553.5–827.3) | 375.1 (274.7–500.4) | 621.6 (497.2–767.7) | 516.3 (414.6–635.3) |
| | 2018 | 721.3 (591.7–870.9) | 492.2 (377.4–631.0) | 497.5 (388.6–627.6) | 429.3 (336.5–539.8) |
| | 2019 | 736.9 (606.8–886.7) | 435.4 (328.9–565.3) | 380.6 (287.5–494.3) | 567.1 (459.4–692.6) |
| Fraser East | 2010 | 1648.8 (1544.7–1758.0) | 483.9 (422.9–551.3) | 637.7 (569.9–711.4) | 743.0 (667.2–825.1) |
| | 2011 | 1521.6 (1421.1–1627.3) | 542.2 (477.3–613.4) | 577.1 (512.1–648.0) | 593.9 (527.3–666.6) |
| | 2012 | 1372.9 (1277.0–1474.1) | 481.1 (420.0–548.6) | 612.8 (545.3–686.4) | 649.7 (580.4–725.1) |
| | 2013 | 1336.1 (1241.4–1436.1) | 402.6 (346.9–464.8) | 678.6 (606.9–756.4) | 642.6 (574.4–716.7) |
| | 2014 | 1467.7 (1368.9–1571.9) | 493.2 (431.4–561.4) | 644.5 (574.4–720.8) | 654.7 (586.6–728.4) |
| | 2015 | 1244.0 (1154.0–1339.2) | 498.7 (437.1–566.6) | 678.2 (606.6–756.0) | 644.4 (577.8–716.7) |
| | 2016 | 1273.5 (1183.6–1368.3) | 505.8 (444.5–573.3) | 519.4 (457.9–587.0) | 647.5 (581.4–719.0) |
| | 2017 | 1193.7 (1107.9–1284.4) | 529.9 (467.8–597.9) | 521.0 (459.9–587.9) | 604.7 (541.2–673.6) |
| | 2018 | 1117.1 (1034.9–1204.1) | 442.2 (386.3–503.9) | 563.7 (500.8–632.4) | 679.9 (612.5–752.6) |
| | 2019 | 1308.8 (1220.7–1401.6) | 478.3 (420.9–541.4) | 585.9 (522.7–654.7) | 635.7 (570.6–706.1) |
| Fraser North | 2010 | 1294.9 (1228.3–1364.1) | 415.7 (378.6–455.6) | 520.6 (482.8–560.6) | 565.8 (522.4–611.8) |
| | 2011 | 1067.6 (1007.1–1130.9) | 347.5 (313.7–383.9) | 469.8 (433.7–508.1) | 467.3 (428.6–508.6) |
| | 2012 | 998.3 (939.6–1059.8) | 373.0 (338.1–410.5) | 422.5 (388.0–459.2) | 435.3 (398.4–474.6) |
| | 2013 | 1013.7 (954.2–1075.8) | 338.1 (304.9–373.9) | 465.5 (428.9–504.5) | 489.8 (451.1–531.0) |
| | 2014 | 909.3 (853.3–968.1) | 349.5 (316.0–385.6) | 433.6 (397.9–471.5) | 453.3 (416.4–492.5) |
| | 2015 | 967.4 (909.9–1027.6) | 358.4 (324.7–394.7) | 454.0 (417.6–492.9) | 478.6 (441.0–518.5) |
| | 2016 | 922.2 (866.4–980.6) | 322.1 (290.4–356.2) | 444.9 (408.8–483.3) | 467.9 (430.9–507.2) |
| | 2017 | 904.4 (849.5–961.9) | 377.2 (343.2–413.7) | 465.7 (429.0–504.8) | 464.7 (428.0–503.8) |
| | 2018 | 864.4 (811.1–920.2) | 384.5 (350.5–420.8) | 427.5 (392.5–464.9) | 438.5 (403.0–476.4) |
| | 2019 | 848.2 (795.9–903.1) | 388.3 (354.7–424.1) | 493.6 (456.2–533.3) | 477.3 (440.4–516.5) |

| | | Incidence rate per 100,000 people (age group) | | | |
|---|---|---|---|---|---|
| **Health Service Delivery Area** | **Year** | **1-19** | **20-34** | **35-49** | **50-64** |
| Fraser South | 2010 | 1545.3 (1481.5–1611.1) | 496.3 (457.7–537.3) | 639.3 (598.4–682.2) | 656.6 (612.5–702.9) |
| | 2011 | 1317.4 (1258.7–1378.1) | 414.7 (379.6–452.1) | 507.5 (471.1–545.8) | 567.9 (527.5–610.5) |
| | 2012 | 1217.0 (1160.8–1275.2) | 428.2 (392.7–466.0) | 555.8 (517.7–595.9) | 582.6 (542.2–625.3) |
| | 2013 | 1291.1 (1233.4–1350.9) | 422.9 (387.8–460.5) | 528.7 (491.5–567.9) | 612.9 (571.9–656.2) |
| | 2014 | 1329.4 (1271.1–1389.7) | 459.3 (422.8–498.1) | 548.1 (510.3–587.9) | 604.7 (564.3–647.1) |
| | 2015 | 1231.2 (1175.6–1288.9) | 421.3 (386.7–458.3) | 554.0 (516.2–593.8) | 584.1 (544.8–625.4) |
| | 2016 | 1087.4 (1035.8–1140.9) | 442.4 (407.3–479.7) | 528.8 (492.2–567.3) | 586.6 (547.5–627.8) |
| | 2017 | 1044.7 (994.7–1096.6) | 461.8 (426.5–499.2) | 547.5 (510.7–586.3) | 595.9 (556.6–637.2) |
| | 2018 | 1026.9 (978.0–1077.6) | 393.9 (362.4–427.4) | 514.3 (479.0–551.6) | 552.8 (515.2–592.4) |
| | 2019 | 1031.4 (982.8–1081.8) | 423.8 (392.3–457.2) | 516.1 (481.1–553.0) | 581.6 (543.1–622.0) |
| Kootenay Boundary | 2010 | 1125.4 (953.1–1319.9) | 467.6 (349.2–613.2) | 509.2 (399.8–639.3) | 503.5 (407.4–615.5) |
| | 2011 | 1091.7 (920.7–1285.3) | 411.4 (300.1–550.4) | 635.1 (510.7–780.7) | 586.8 (483.2–706.1) |
| | 2012 | 1014.0 (847.2–1204.0) | 450.1 (331.8–596.7) | 659.2 (530.7–809.3) | 613.3 (506.8–735.6) |
| | 2013 | 910.5 (751.0–1093.8) | 462.1 (340.7–612.7) | 500.9 (388.2–636.1) | 577.8 (474.0–697.7) |
| | 2014 | 841.1 (687.2–1019.1) | 595.3 (455.4–764.7) | 547.9 (428.7–690.0) | 521.6 (423.0–636.4) |
| | 2015 | 815.3 (664.8–989.7) | 443.8 (324.9–591.9) | 418.8 (315.5–545.1) | 614.5 (506.9–738.1) |
| | 2016 | 634.4 (503.0–789.5) | 332.3 (231.4–462.1) | 351.0 (257.9–466.8) | 507.9 (410.0–622.2) |
| | 2017 | 977.3 (814.1–1163.6) | 387.5 (279.2–523.7) | 350.3 (258.3–464.4) | 314.5 (238.2–407.4) |
| | 2018 | 655.6 (523.6–810.6) | 391.4 (283.3–527.3) | 448.4 (344.6–573.7) | 374.2 (290.0–475.2) |
| | 2019 | 835.6 (686.1–1008.0) | 505.7 (383.0–655.2) | 510.6 (400.3–642.0) | 459.3 (364.8–570.9) |
| North Shore/Coast Garibaldi | 2010 | 1080.6 (991.3–1175.8) | 505.3 (441.1–576.1) | 551.7 (492.6–615.9) | 539.9 (482.0–602.8) |
| | 2011 | 955.8 (872.4–1044.9) | 435.8 (376.3–502.0) | 498.9 (442.5–560.4) | 482.4 (428.2–541.6) |
| | 2012 | 862.8 (783.7–947.7) | 452.7 (392.1–520.0) | 462.6 (408.1–522.4) | 426.2 (375.5–481.8) |
| | 2013 | 966.9 (883.1–1056.5) | 395.9 (339.5–458.9) | 489.6 (433.0–551.5) | 416.1 (366.3–470.7) |
| | 2014 | 939.5 (857.1–1027.6) | 370.8 (316.4–431.9) | 451.8 (397.4–511.6) | 430.2 (379.8–485.5) |
| | 2015 | 955.8 (872.8–1044.6) | 425.0 (366.3–490.5) | 445.4 (391.3–504.8) | 438.1 (387.1–493.9) |
| | 2016 | 862.1 (783.2–946.7) | 427.0 (368.1–492.5) | 430.8 (377.8–489.1) | 454.0 (401.8–511.0) |
| | 2017 | 832.2 (754.9–915.2) | 412.0 (354.5–476.2) | 418.1 (366.1–475.4) | 456.4 (404.1–513.7) |
| | 2018 | 859.9 (781.8–943.7) | 383.7 (328.6–445.3) | 414.4 (362.9–471.1) | 452.1 (400.0–509.2) |
| | 2019 | 939.2 (857.9–1026.1) | 437.7 (379.5–502.4) | 458.2 (404.5–517.1) | 472.3 (419.0–530.5) |
| North Vancouver Island | 2010 | 1143.1 (1001.9–1298.7) | 460.8 (360.5–580.3) | 548.4 (453.1–657.7) | 473.0 (395.5–561.3) |
| | 2011 | 979.0 (847.7–1124.9) | 504.2 (398.6–629.3) | 549.5 (452.5–661.2) | 420.1 (347.7–503.1) |
| | 2012 | 952.0 (821.8–1097.1) | 402.5 (307.9–517.0) | 446.2 (357.4–550.4) | 422.8 (349.9–506.3) |
| | 2013 | 973.1 (840.4–1120.9) | 378.8 (286.9–490.8) | 414.7 (327.8–517.6) | 477.2 (399.6–565.6) |
| | 2014 | 969.5 (836.6–1117.6) | 487.1 (381.8–612.5) | 484.9 (389.4–596.7) | 473.7 (396.4–561.8) |
| | 2015 | 1181.4 (1034.5–1343.2) | 356.4 (267.7–465.0) | 499.9 (403.0–613.1) | 498.1 (419.0–587.8) |
| | 2016 | 986.3 (854.0–1133.3) | 360.6 (272.4–468.3) | 480.1 (386.1–590.1) | 516.0 (435.7–606.8) |
| | 2017 | 871.5 (748.8–1008.5) | 347.7 (262.0–452.6) | 482.0 (389.0–590.5) | 488.0 (410.0–576.6) |
| | 2018 | 985.0 (855.3–1128.7) | 379.4 (290.2–487.4) | 525.9 (429.7–637.2) | 546.9 (464.0–640.5) |
| | 2019 | 1020.0 (888.5–1165.5) | 554.3 (446.3–680.6) | 506.5 (413.0–614.8) | 477.3 (399.3–566.0) |
| Northeast | 2010 | 901.4 (762.0–1058.9) | 432.9 (331.9–555.0) | 575.7 (455.8–717.5) | 432.8 (320.2–572.2) |
| | 2011 | 946.8 (804.0–1107.6) | 428.2 (329.1–547.9) | 490.7 (380.3–623.2) | 529.7 (406.1–679.0) |
| | 2012 | 999.2 (852.5–1163.8) | 362.5 (273.1–471.8) | 416.0 (314.3–540.3) | 514.0 (394.1–658.9) |
| | 2013 | 885.2 (747.4–1041.0) | 395.3 (302.4–507.8) | 332.9 (241.9–446.8) | 362.1 (263.1–486.2) |
| | 2014 | 709.8 (587.0–850.7) | 321.4 (238.6–423.8) | 331.7 (240.0–446.8) | 320.7 (228.0–438.4) |
| | 2015 | 785.0 (654.9–933.4) | 351.2 (263.1–459.4) | 376.6 (277.7–499.3) | 372.6 (271.8–498.6) |
| | 2016 | 915.0 (773.0–1075.4) | 364.1 (271.9–477.4) | 311.4 (221.4–425.6) | 485.0 (368.3–627.0) |
| | 2017 | 697.8 (575.1–838.9) | 330.2 (241.7–440.4) | 327.3 (234.9–444.0) | 437.0 (326.4–573.0) |
| | 2018 | 526.1 (420.8–649.7) | 342.5 (251.6–455.4) | 229.1 (153.4–329.0) | 353.9 (255.1–478.4) |

| | | Incidence rate per 100,000 people (age group) | | | |
|---|---|---|---|---|---|
| **Health Service Delivery Area** | **Year** | **1-19** | **20-34** | **35-49** | **50-64** |
| | 2019 | 610.6 (497.4–742.0) | 311.5 (225.5–419.7) | 296.1 (209.5–406.4) | 338.2 (241.6–460.5) |
| Northern Interior | 2010 | 1087.9 (972.1–1213.6) | 455.1 (375.0–547.1) | 466.4 (391.0–552.0) | 373.0 (305.7–450.8) |
| | 2011 | 1087.0 (970.6–1213.5) | 424.3 (347.0–513.6) | 409.8 (338.3–491.9) | 457.0 (382.9–541.2) |
| | 2012 | 1018.1 (905.2–1141.2) | 397.0 (322.3–483.8) | 469.3 (391.6–558.1) | 354.9 (290.3–429.6) |
| | 2013 | 926.5 (818.1–1045.3) | 399.6 (324.8–486.5) | 473.6 (394.2–564.3) | 371.1 (305.2–446.8) |
| | 2014 | 972.1 (860.2–1094.5) | 443.9 (364.1–535.9) | 400.1 (326.3–485.7) | 489.8 (413.5–576.0) |
| | 2015 | 1243.2 (1115.4–1381.5) | 467.8 (385.5–562.4) | 491.7 (408.3–587.1) | 461.3 (387.0–545.6) |
| | 2016 | 918.1 (808.6–1038.2) | 413.4 (336.3–502.8) | 488.5 (405.0–584.1) | 429.7 (357.7–511.9) |
| | 2017 | 906.7 (798.4–1025.6) | 448.9 (369.0–541.1) | 459.6 (378.8–552.6) | 452.0 (377.9–536.4) |
| | 2018 | 770.9 (672.0–880.3) | 397.7 (323.3–484.2) | 470.2 (388.6–564.0) | 443.0 (369.3–527.0) |
| | 2019 | 731.2 (635.0–837.9) | 316.7 (251.2–394.2) | 356.1 (285.6–438.8) | 326.5 (263.5–400.0) |
| Northwest | 2010 | 960.1 (818.8–1118.8) | 404.4 (300.2–533.2) | 539.0 (428.7–669.1) | 593.0 (477.5–728.1) |
| | 2011 | 1036.6 (888.3–1202.6) | 354.1 (257.3–475.4) | 430.5 (331.5–549.7) | 617.1 (499.9–753.6) |
| | 2012 | 926.3 (785.4–1085.3) | 377.6 (277.4–502.1) | 484.3 (377.5–611.8) | 446.0 (347.7–563.5) |
| | 2013 | 983.1 (837.1–1147.3) | 448.7 (339.9–581.4) | 477.3 (369.9–606.2) | 579.7 (466.8–711.8) |
| | 2014 | 978.9 (831.7–1144.5) | 449.6 (340.5–582.5) | 574.6 (454.9–716.2) | 500.6 (395.7–624.7) |
| | 2015 | 1012.5 (860.8–1183.3) | 388.0 (286.1–514.5) | 562.1 (442.2–704.6) | 598.8 (482.7–734.4) |
| | 2016 | 793.9 (659.3–947.9) | 398.8 (295.0–527.2) | 391.8 (291.7–515.1) | 410.6 (314.8–526.4) |
| | 2017 | 743.3 (613.1–892.9) | 343.3 (247.4–464.1) | 521.2 (403.9–661.9) | 397.3 (302.4–512.5) |
| | 2018 | 774.9 (642.5–926.6) | 398.6 (294.9–527.0) | 414.8 (310.7–542.5) | 468.0 (364.1–592.3) |
| | 2019 | 825.7 (689.3–981.0) | 452.8 (342.9–586.6) | 531.5 (412.7–673.7) | 409.5 (312.5–527.1) |
| Okanagan | 2010 | 1306.5 (1215.0–1403.1) | 537.4 (476.5–604.0) | 610.2 (549.2–676.1) | 596.3 (541.0–655.8) |
| | 2011 | 1125.7 (1040.4–1216.1) | 550.4 (488.3–618.1) | 557.7 (498.8–621.6) | 554.3 (501.5–611.1) |
| | 2012 | 1088.2 (1004.0–1177.7) | 490.8 (432.0–555.5) | 597.4 (535.6–664.4) | 549.5 (497.3–605.7) |
| | 2013 | 1073.8 (989.7–1163.1) | 439.5 (383.7–501.2) | 549.2 (489.4–614.3) | 574.0 (521.0–630.9) |
| | 2014 | 1099.4 (1014.3–1189.7) | 487.7 (429.1–552.1) | 513.6 (455.6–577.0) | 576.8 (524.2–633.3) |
| | 2015 | 1034.0 (952.0–1121.2) | 478.1 (420.6–541.1) | 593.0 (530.7–660.5) | 584.5 (532.0–640.8) |
| | 2016 | 992.0 (912.8–1076.3) | 412.5 (360.1–470.5) | 505.4 (448.6–567.4) | 547.4 (497.0–601.5) |
| | 2017 | 1091.5 (1009.3–1178.7) | 479.9 (423.7–541.4) | 527.1 (469.8–589.3) | 570.6 (519.4–625.5) |
| | 2018 | 886.9 (813.7–965.0) | 506.7 (449.8–568.9) | 499.3 (444.4–559.1) | 608.0 (555.1–664.6) |
| | 2019 | 982.4 (905.9–1063.5) | 506.4 (450.2–567.6) | 508.4 (453.7–567.8) | 575.8 (524.3–631.0) |
| Richmond | 2010 | 1087.5 (983.0–1200.0) | 356.1 (297.5–422.9) | 480.2 (417.5–549.7) | 452.7 (391.3–521.1) |
| | 2011 | 960.8 (862.1–1067.8) | 339.4 (282.5–404.4) | 296.9 (247.7–353.0) | 434.1 (374.4–500.6) |
| | 2012 | 870.2 (775.8–972.8) | 291.1 (238.8–351.4) | 371.2 (315.4–434.1) | 388.7 (332.7–451.6) |
| | 2013 | 942.8 (843.9–1050.0) | 346.7 (289.3–412.3) | 359.2 (303.1–422.7) | 409.5 (352.2–473.5) |
| | 2014 | 908.2 (810.4–1014.6) | 358.1 (299.4–425.0) | 396.7 (336.7–464.3) | 394.9 (338.7–457.8) |
| | 2015 | 861.6 (765.6–966.3) | 400.7 (338.4–471.3) | 436.8 (372.9–508.5) | 457.1 (396.4–524.5) |
| | 2016 | 815.0 (720.9–918.1) | 284.2 (232.2–344.4) | 481.4 (413.4–557.3) | 452.6 (392.1–519.9) |
| | 2017 | 842.2 (746.6–946.6) | 319.3 (264.5–382.1) | 325.4 (270.0–388.9) | 440.2 (380.3–506.9) |
| | 2018 | 912.0 (813.0–1019.8) | 372.7 (314.1–439.0) | 419.0 (355.8–490.2) | 479.8 (417.4–548.9) |
| | 2019 | 933.3 (834.1–1041.0) | 356.9 (300.4–420.9) | 480.0 (412.5–555.5) | 477.3 (415.2–546.0) |
| South Vancouver Island | 2010 | 1224.7 (1133.6–1321.1) | 553.6 (495.3–616.9) | 614.3 (555.6–677.6) | 507.4 (457.8–560.9) |
| | 2011 | 1205.4 (1114.9–1301.2) | 441.6 (389.5–498.6) | 516.2 (462.0–575.0) | 472.8 (425.1–524.3) |
| | 2012 | 1091.5 (1005.5–1182.9) | 412.9 (362.6–468.2) | 464.5 (412.8–520.8) | 436.5 (390.8–486.1) |
| | 2013 | 1252.7 (1160.5–1350.2) | 398.3 (348.8–452.8) | 496.9 (442.9–555.6) | 438.6 (392.8–488.3) |
| | 2014 | 1151.4 (1063.1–1245.0) | 366.6 (319.2–419.0) | 488.8 (435.0–547.5) | 476.2 (428.4–527.8) |
| | 2015 | 1207.2 (1117.4–1302.3) | 428.0 (377.0–484.0) | 502.0 (447.7–561.2) | 505.6 (456.5–558.6) |
| | 2016 | 1029.6 (947.9–1116.5) | 399.3 (350.8–452.5) | 449.1 (398.4–504.3) | 478.4 (430.7–529.9) |
| | 2017 | 954.1 (876.2–1037.2) | 405.6 (357.4–458.6) | 432.2 (383.1–485.8) | 459.7 (412.9–510.4) |

| | | Incidence rate per 100,000 people (age group) | | | |
|---|---|---|---|---|---|
| **Health Service Delivery Area** | **Year** | **1-19** | **20-34** | **35-49** | **50-64** |
| | 2018 | 865.4 (791.8–943.9) | 394.1 (347.2–445.6) | 448.1 (398.7–501.8) | 457.5 (410.7–508.3) |
| | 2019 | 975.1 (897.7–1057.4) | 473.2 (422.5–528.4) | 545.0 (491.3–603.0) | 538.7 (487.7–593.6) |
| Thompson Cariboo Shuswap | 2010 | 1159.4 (1055.3–1271.0) | 472.8 (402.8–551.5) | 578.8 (506.5–658.7) | 610.2 (542.0–684.6) |
| | 2011 | 991.5 (894.3–1096.4) | 479.6 (408.6–559.4) | 457.0 (391.8–530.0) | 518.6 (456.3–587.0) |
| | 2012 | 1204.0 (1095.8–1320.0) | 318.1 (260.4–384.7) | 520.2 (449.2–599.2) | 479.4 (419.4–545.5) |
| | 2013 | 1124.0 (1018.4–1237.5) | 385.9 (321.9–458.8) | 534.0 (461.0–615.4) | 538.5 (474.8–608.3) |
| | 2014 | 1176.7 (1068.0–1293.5) | 445.0 (376.5–522.5) | 527.2 (453.8–609.1) | 585.7 (519.4–658.3) |
| | 2015 | 1107.7 (1002.8–1220.5) | 449.7 (381.1–527.2) | 508.7 (436.5–589.4) | 577.0 (511.4–648.7) |
| | 2016 | 1069.7 (967.5–1179.9) | 413.8 (348.5–487.7) | 477.2 (407.8–555.1) | 471.4 (412.5–536.4) |
| | 2017 | 1062.1 (961.0–1170.9) | 449.5 (381.9–525.5) | 580.9 (504.6–665.4) | 645.8 (576.5–721.2) |
| | 2018 | 819.0 (731.3–914.3) | 442.9 (377.1–516.8) | 619.2 (541.1–705.4) | 564.8 (499.9–635.8) |
| | 2019 | 957.6 (863.1–1059.6) | 417.3 (353.8–488.9) | 579.8 (504.7–662.9) | 640.2 (570.7–715.9) |
| Vancouver | 2010 | 1164.6 (1096.0–1236.5) | 488.5 (452.8–526.3) | 536.6 (500.1–574.9) | 535.1 (493.5–579.2) |
| | 2011 | 1042.7 (977.9–1110.7) | 429.8 (396.6–465.1) | 431.2 (398.5–465.8) | 434.0 (397.1–473.5) |
| | 2012 | 960.1 (897.9–1025.5) | 393.7 (362.1–427.2) | 476.8 (442.2–513.3) | 437.0 (400.1–476.3) |
| | 2013 | 952.3 (890.2–1017.5) | 392.6 (361.1–426.1) | 466.6 (431.9–503.4) | 469.3 (431.2–509.8) |
| | 2014 | 915.7 (854.7–979.9) | 442.7 (409.2–478.2) | 433.0 (399.2–468.9) | 435.0 (398.5–474.0) |
| | 2015 | 967.3 (904.1–1033.7) | 452.3 (418.4–488.2) | 464.4 (429.1–501.9) | 490.1 (451.2–531.4) |
| | 2016 | 863.0 (802.9–926.3) | 414.5 (382.3–448.7) | 474.8 (438.8–512.9) | 489.3 (450.4–530.7) |
| | 2017 | 846.0 (786.5–908.9) | 426.9 (394.6–461.1) | 458.7 (423.4–496.3) | 478.7 (440.2–519.6) |
| | 2018 | 832.7 (774.1–894.6) | 439.6 (407.5–473.6) | 455.2 (420.1–492.5) | 476.1 (437.8–516.8) |
| | 2019 | 937.7 (875.7–1002.9) | 499.7 (466.1–535.1) | 514.5 (477.4–553.8) | 443.6 (406.8–482.7) |

**Appendix 3.** Annual prevalence rate of asthma in British Columbia by Health Service Delivery Area and age group (1-19, 20-34, 35-49, and 50-64) from 2010 to 2019. Data sourced from the British Columbia Centre for Disease Control (BCCDC) Chronic Disease Dashboard[25].

| | | Prevalence rate (%) | | | |
|---|---|---|---|---|---|
| **Health Service Delivery Area** | **Year** | **1-19** | **20-34** | **35-49** | **50-64** |
| Central Vancouver Island | 2010 | 15.1 (14.7–15.4) | 15.3 (14.9–15.7) | 10.5 (10.2–10.8) | 9.6 (9.4–9.9) |
| | 2011 | 14.9 (14.5–15.2) | 15.9 (15.6–16.3) | 10.7 (10.4–11.0) | 9.9 (9.6–10.1) |
| | 2012 | 14.6 (14.3–15.0) | 16.6 (16.2–17.0) | 11.1 (10.8–11.4) | 10.2 (10.0–10.5) |
| | 2013 | 14.6 (14.2–14.9) | 17.1 (16.7–17.5) | 11.5 (11.2–11.9) | 10.5 (10.3–10.8) |
| | 2014 | 14.3 (14.0–14.7) | 17.5 (17.1–17.9) | 11.9 (11.6–12.2) | 10.8 (10.5–11.0) |
| | 2015 | 13.9 (13.6–14.3) | 17.8 (17.4–18.2) | 12.4 (12.1–12.7) | 11.1 (10.9–11.4) |
| | 2016 | 13.6 (13.2–13.9) | 18.0 (17.6–18.4) | 12.9 (12.6–13.2) | 11.4 (11.2–11.7) |
| | 2017 | 13.1 (12.8–13.4) | 18.3 (17.9–18.7) | 13.2 (12.9–13.5) | 11.8 (11.6–12.1) |
| | 2018 | 12.6 (12.3–12.9) | 18.4 (18.0–18.8) | 13.6 (13.2–13.9) | 12.1 (11.9–12.4) |
| | 2019 | 12.3 (12.0–12.6) | 18.7 (18.3–19.1) | 14.0 (13.6–14.3) | 12.6 (12.3–12.9) |
| East Kootenay | 2010 | 10.3 (9.9–10.8) | 9.2 (8.7–9.7) | 7.8 (7.4–8.3) | 7.4 (7.0–7.8) |
| | 2011 | 10.4 (9.9–10.9) | 9.9 (9.4–10.4) | 8.4 (7.9–8.8) | 7.6 (7.2–8.0) |
| | 2012 | 10.6 (10.1–11.1) | 10.6 (10.0–11.1) | 8.8 (8.3–9.3) | 8.0 (7.6–8.4) |
| | 2013 | 10.2 (9.7–10.7) | 11.1 (10.5–11.6) | 9.0 (8.5–9.5) | 8.3 (7.9–8.7) |
| | 2014 | 10.0 (9.5–10.5) | 11.6 (11.0–12.1) | 9.1 (8.7–9.6) | 8.5 (8.1–8.9) |
| | 2015 | 9.8 (9.3–10.3) | 12.1 (11.6–12.7) | 9.4 (8.9–9.9) | 8.7 (8.3–9.2) |
| | 2016 | 9.9 (9.4–10.4) | 12.3 (11.8–13.0) | 9.7 (9.2–10.2) | 9.2 (8.8–9.6) |
| | 2017 | 9.5 (9.0–10.0) | 12.4 (11.8–13.0) | 10.1 (9.6–10.6) | 9.5 (9.1–9.9) |
| | 2018 | 9.3 (8.8–9.7) | 12.4 (11.9–13.0) | 10.3 (9.8–10.8) | 9.8 (9.4–10.3) |
| | 2019 | 9.0 (8.5–9.5) | 12.7 (12.2–13.3) | 10.2 (9.7–10.7) | 10.1 (9.6–10.5) |
| Fraser East | 2010 | 17.5 (17.2–17.8) | 13.5 (13.2–13.8) | 12.0 (11.7–12.2) | 11.6 (11.3–11.9) |
| | 2011 | 17.7 (17.4–18.0) | 14.4 (14.1–14.7) | 12.4 (12.1–12.7) | 11.9 (11.6–12.2) |
| | 2012 | 17.7 (17.3–18.0) | 15.2 (14.9–15.5) | 12.8 (12.5–13.1) | 12.4 (12.1–12.7) |
| | 2013 | 17.6 (17.3–17.9) | 15.8 (15.5–16.1) | 13.1 (12.8–13.5) | 12.7 (12.4–13.0) |
| | 2014 | 17.4 (17.1–17.7) | 16.7 (16.4–17.0) | 13.7 (13.4–14.0) | 13.2 (12.9–13.5) |
| | 2015 | 17.0 (16.7–17.3) | 17.4 (17.0–17.7) | 14.2 (13.8–14.5) | 13.6 (13.3–13.9) |
| | 2016 | 16.6 (16.3–17.0) | 18.1 (17.7–18.4) | 14.4 (14.1–14.7) | 13.9 (13.6–14.2) |
| | 2017 | 16.1 (15.8–16.4) | 18.5 (18.2–18.9) | 14.6 (14.3–14.9) | 14.3 (14.0–14.6) |
| | 2018 | 15.6 (15.4–15.9) | 18.8 (18.5–19.1) | 14.7 (14.4–15.0) | 14.8 (14.5–15.1) |
| | 2019 | 15.3 (15.1–15.6) | 18.9 (18.5–19.2) | 14.8 (14.5–15.1) | 15.3 (15.0–15.6) |
| Fraser North | 2010 | 13.3 (13.1–13.5) | 10.5 (10.3–10.7) | 8.3 (8.1–8.4) | 9.6 (9.4–9.7) |
| | 2011 | 13.3 (13.1–13.5) | 10.9 (10.8–11.1) | 8.5 (8.3–8.6) | 9.7 (9.6–9.9) |
| | 2012 | 13.2 (13.0–13.4) | 11.4 (11.2–11.6) | 8.8 (8.6–8.9) | 9.9 (9.7–10.1) |
| | 2013 | 13.1 (12.9–13.3) | 11.9 (11.7–12.0) | 9.1 (9.0–9.3) | 10.2 (10.1–10.4) |
| | 2014 | 12.8 (12.6–13.0) | 12.3 (12.1–12.5) | 9.4 (9.2–9.5) | 10.4 (10.2–10.6) |
| | 2015 | 12.6 (12.4–12.8) | 12.7 (12.5–12.8) | 9.6 (9.4–9.7) | 10.7 (10.5–10.9) |
| | 2016 | 12.2 (12.0–12.4) | 12.9 (12.7–13.1) | 9.7 (9.6–9.9) | 10.9 (10.8–11.1) |
| | 2017 | 11.9 (11.7–12.1) | 13.2 (13.0–13.3) | 9.9 (9.7–10.0) | 11.2 (11.0–11.4) |
| | 2018 | 11.5 (11.4–11.7) | 13.2 (13.0–13.4) | 10.0 (9.9–10.2) | 11.4 (11.2–11.6) |
| | 2019 | 11.2 (11.0–11.3) | 13.2 (13.0–13.3) | 10.2 (10.1–10.4) | 11.6 (11.4–11.8) |

| | | Prevalence rate (%) | | | |
|---|---|---|---|---|---|
| **Health Service Delivery Area** | **Year** | **1-19** | **20-34** | **35-49** | **50-64** |
| Fraser South | 2010 | 16.9 (16.7–17.1) | 12.7 (12.5–12.9) | 10.3 (10.2–10.5) | 11.0 (10.8–11.2) |
| | 2011 | 16.7 (16.5–16.9) | 13.4 (13.2–13.6) | 10.6 (10.4–10.7) | 11.3 (11.1–11.5) |
| | 2012 | 16.5 (16.3–16.7) | 14.0 (13.8–14.2) | 10.8 (10.6–11.0) | 11.7 (11.5–11.9) |
| | 2013 | 16.3 (16.1–16.5) | 14.6 (14.4–14.8) | 11.0 (10.8–11.1) | 12.1 (11.9–12.2) |
| | 2014 | 16.2 (16.0–16.4) | 15.1 (14.9–15.3) | 11.2 (11.1–11.4) | 12.4 (12.2–12.6) |
| | 2015 | 15.8 (15.6–16.0) | 15.7 (15.5–15.9) | 11.4 (11.2–11.5) | 12.7 (12.5–12.9) |
| | 2016 | 15.3 (15.1–15.5) | 15.9 (15.7–16.1) | 11.5 (11.3–11.7) | 12.9 (12.7–13.1) |
| | 2017 | 14.8 (14.6–15.0) | 15.9 (15.8–16.1) | 11.6 (11.4–11.8) | 13.2 (13.1–13.4) |
| | 2018 | 14.1 (14.0–14.3) | 15.5 (15.3–15.7) | 11.7 (11.5–11.8) | 13.4 (13.3–13.6) |
| | 2019 | 13.8 (13.6–13.9) | 14.9 (14.7–15.0) | 11.7 (11.6–11.9) | 13.7 (13.5–13.8) |
| Kootenay Boundary | 2010 | 12.5 (11.9–13.1) | 11.3 (10.7–11.9) | 8.2 (7.8–8.7) | 7.8 (7.4–8.2) |
| | 2011 | 12.5 (12.0–13.1) | 11.8 (11.2–12.4) | 8.9 (8.4–9.3) | 8.0 (7.6–8.4) |
| | 2012 | 12.5 (11.9–13.1) | 12.5 (11.9–13.1) | 9.1 (8.6–9.6) | 8.5 (8.1–8.9) |
| | 2013 | 12.4 (11.9–13.0) | 13.1 (12.4–13.7) | 9.6 (9.1–10.2) | 9.0 (8.6–9.4) |
| | 2014 | 12.1 (11.5–12.7) | 14.0 (13.3–14.7) | 10.1 (9.6–10.7) | 9.1 (8.7–9.5) |
| | 2015 | 11.5 (11.0–12.1) | 14.1 (13.4–14.7) | 10.6 (10.1–11.1) | 9.5 (9.1–10.0) |
| | 2016 | 10.9 (10.4–11.5) | 14.1 (13.5–14.8) | 10.6 (10.1–11.1) | 9.8 (9.4–10.3) |
| | 2017 | 10.7 (10.2–11.3) | 14.1 (13.5–14.8) | 10.9 (10.4–11.5) | 10.2 (9.8–10.7) |
| | 2018 | 10.3 (9.8–10.8) | 14.5 (13.8–15.2) | 11.2 (10.7–11.7) | 10.4 (9.9–10.8) |
| | 2019 | 10.1 (9.6–10.7) | 14.8 (14.1–15.4) | 11.4 (10.9–11.9) | 10.8 (10.3–11.2) |
| North Shore/Coast Garibaldi | 2010 | 12.2 (11.9–12.5) | 10.5 (10.2–10.8) | 8.5 (8.3–8.8) | 8.9 (8.7–9.2) |
| | 2011 | 11.9 (11.6–12.2) | 11.1 (10.8–11.4) | 8.9 (8.7–9.2) | 9.2 (8.9–9.4) |
| | 2012 | 11.7 (11.4–12.0) | 11.6 (11.3–11.9) | 9.0 (8.8–9.3) | 9.5 (9.3–9.8) |
| | 2013 | 11.6 (11.4–11.9) | 12.1 (11.8–12.4) | 9.4 (9.2–9.7) | 9.7 (9.5–10.0) |
| | 2014 | 11.5 (11.2–11.7) | 12.2 (11.9–12.5) | 9.6 (9.4–9.9) | 10.0 (9.7–10.2) |
| | 2015 | 11.2 (11.0–11.5) | 12.5 (12.2–12.8) | 9.7 (9.5–10.0) | 10.3 (10.0–10.5) |
| | 2016 | 11.0 (10.7–11.3) | 12.7 (12.4–13.1) | 9.8 (9.5–10.0) | 10.6 (10.3–10.8) |
| | 2017 | 10.8 (10.5–11.1) | 12.9 (12.6–13.2) | 9.8 (9.6–10.1) | 10.9 (10.6–11.1) |
| | 2018 | 10.6 (10.3–10.9) | 12.8 (12.5–13.2) | 10.0 (9.7–10.2) | 11.1 (10.9–11.4) |
| | 2019 | 10.5 (10.2–10.8) | 12.9 (12.6–13.2) | 10.0 (9.8–10.3) | 11.4 (11.2–11.7) |
| North Vancouver Island | 2010 | 15.6 (15.1–16.1) | 15.7 (15.1–16.3) | 10.4 (9.9–10.8) | 9.4 (9.1–9.7) |
| | 2011 | 15.3 (14.8–15.8) | 16.4 (15.8–17.0) | 10.8 (10.3–11.2) | 9.6 (9.2–9.9) |
| | 2012 | 15.0 (14.5–15.5) | 16.9 (16.3–17.5) | 11.2 (10.7–11.6) | 10.0 (9.6–10.3) |
| | 2013 | 14.6 (14.1–15.1) | 17.2 (16.6–17.8) | 11.5 (11.0–12.0) | 10.3 (10.0–10.7) |
| | 2014 | 14.2 (13.7–14.7) | 17.9 (17.2–18.5) | 11.9 (11.5–12.4) | 10.8 (10.5–11.2) |
| | 2015 | 14.0 (13.5–14.5) | 18.2 (17.6–18.8) | 12.4 (11.9–12.9) | 11.3 (10.9–11.7) |
| | 2016 | 13.6 (13.2–14.1) | 18.4 (17.8–19.0) | 12.8 (12.3–13.3) | 11.7 (11.3–12.1) |
| | 2017 | 13.1 (12.6–13.6) | 18.9 (18.3–19.5) | 13.0 (12.5–13.5) | 11.9 (11.5–12.3) |
| | 2018 | 12.8 (12.3–13.2) | 19.1 (18.5–19.7) | 13.4 (13.0–13.9) | 12.3 (11.9–12.7) |
| | 2019 | 12.6 (12.1–13.0) | 19.3 (18.7–19.9) | 13.7 (13.2–14.2) | 12.6 (12.2–13.0) |
| Northeast | 2010 | 11.3 (10.8–11.8) | 10.9 (10.4–11.4) | 9.0 (8.6–9.5) | 8.6 (8.1–9.1) |
| | 2011 | 11.1 (10.7–11.6) | 11.8 (11.2–12.3) | 9.2 (8.7–9.7) | 8.9 (8.4–9.4) |
| | 2012 | 11.1 (10.6–11.6) | 12.3 (11.8–12.8) | 9.5 (9.0–10.0) | 9.2 (8.7–9.7) |
| | 2013 | 10.9 (10.4–11.4) | 12.8 (12.3–13.3) | 10.0 (9.5–10.5) | 9.3 (8.7–9.8) |
| | 2014 | 10.4 (9.9–10.9) | 13.1 (12.5–13.6) | 10.0 (9.5–10.6) | 9.5 (9.0–10.0) |
| | 2015 | 10.1 (9.6–10.6) | 13.4 (12.8–13.9) | 10.1 (9.6–10.7) | 9.8 (9.3–10.4) |
| | 2016 | 10.0 (9.6–10.5) | 13.8 (13.3–14.4) | 10.3 (9.7–10.8) | 10.2 (9.6–10.7) |
| | 2017 | 9.6 (9.1–10.0) | 14.3 (13.7–14.9) | 10.4 (9.9–11.0) | 10.6 (10.1–11.2) |
| | 2018 | 8.9 (8.5–9.4) | 14.6 (14.0–15.2) | 10.5 (9.9–11.0) | 11.0 (10.4–11.6) |

| | | Prevalence rate (%) | | | |
|---|---|---|---|---|---|
| **Health Service Delivery Area** | **Year** | **1-19** | **20-34** | **35-49** | **50-64** |
| | 2019 | 8.6 (8.2–9.0) | 14.6 (14.0–15.2) | 10.8 (10.3–11.4) | 11.2 (10.6–11.8) |
| Northern Interior | 2010 | 15.7 (15.3–16.2) | 13.7 (13.3–14.1) | 9.9 (9.5–10.2) | 9.6 (9.3–10.0) |
| | 2011 | 15.5 (15.1–15.9) | 14.6 (14.2–15.1) | 10.3 (10.0–10.7) | 9.8 (9.5–10.2) |
| | 2012 | 15.1 (14.7–15.5) | 15.6 (15.2–16.1) | 10.7 (10.3–11.1) | 10.1 (9.8–10.5) |
| | 2013 | 14.7 (14.3–15.1) | 16.1 (15.7–16.6) | 11.1 (10.7–11.5) | 10.3 (9.9–10.6) |
| | 2014 | 14.2 (13.8–14.6) | 16.8 (16.3–17.3) | 11.5 (11.1–11.9) | 10.6 (10.2–10.9) |
| | 2015 | 13.9 (13.5–14.4) | 17.5 (17.1–18.0) | 12.0 (11.5–12.4) | 10.9 (10.5–11.2) |
| | 2016 | 13.5 (13.1–13.9) | 18.1 (17.6–18.5) | 12.5 (12.1–12.9) | 11.3 (10.9–11.6) |
| | 2017 | 13.0 (12.6–13.4) | 18.5 (18.0–19.0) | 12.9 (12.5–13.3) | 11.6 (11.2–11.9) |
| | 2018 | 12.4 (12.0–12.8) | 18.6 (18.1–19.1) | 13.4 (12.9–13.8) | 12.0 (11.7–12.4) |
| | 2019 | 12.0 (11.6–12.3) | 18.6 (18.1–19.1) | 13.7 (13.3–14.2) | 12.3 (11.9–12.6) |
| Northwest | 2010 | 13.0 (12.5–13.5) | 11.5 (10.9–12.1) | 9.1 (8.7–9.6) | 9.3 (8.8–9.8) |
| | 2011 | 13.1 (12.6–13.6) | 12.2 (11.7–12.8) | 9.3 (8.9–9.8) | 9.6 (9.2–10.1) |
| | 2012 | 13.0 (12.5–13.5) | 13.1 (12.5–13.7) | 9.6 (9.1–10.1) | 9.9 (9.4–10.3) |
| | 2013 | 13.0 (12.5–13.5) | 13.6 (13.1–14.3) | 9.9 (9.4–10.4) | 10.3 (9.8–10.8) |
| | 2014 | 12.6 (12.1–13.1) | 14.3 (13.7–15.0) | 10.2 (9.7–10.7) | 10.6 (10.1–11.1) |
| | 2015 | 12.5 (11.9–13.0) | 15.3 (14.7–15.9) | 10.5 (9.9–11.0) | 10.9 (10.4–11.4) |
| | 2016 | 11.8 (11.3–12.4) | 15.9 (15.3–16.6) | 10.5 (10.0–11.1) | 11.4 (10.9–11.9) |
| | 2017 | 11.2 (10.8–11.8) | 16.4 (15.8–17.1) | 11.0 (10.5–11.6) | 11.6 (11.1–12.2) |
| | 2018 | 10.8 (10.3–11.3) | 16.7 (16.1–17.4) | 11.4 (10.9–12.0) | 12.0 (11.4–12.5) |
| | 2019 | 10.5 (10.0–11.0) | 17.0 (16.3–17.6) | 11.9 (11.4–12.5) | 12.1 (11.6–12.7) |
| Okanagan | 2010 | 13.9 (13.6–14.2) | 12.8 (12.6–13.1) | 10.1 (9.8–10.3) | 9.3 (9.1–9.6) |
| | 2011 | 13.8 (13.5–14.0) | 13.6 (13.3–13.9) | 10.5 (10.2–10.7) | 9.7 (9.5–9.9) |
| | 2012 | 13.5 (13.3–13.8) | 14.3 (14.0–14.7) | 10.9 (10.7–11.2) | 10.0 (9.8–10.2) |
| | 2013 | 13.4 (13.1–13.6) | 14.8 (14.5–15.1) | 11.3 (11.0–11.5) | 10.3 (10.1–10.5) |
| | 2014 | 13.1 (12.8–13.4) | 15.3 (15.0–15.6) | 11.5 (11.2–11.7) | 10.6 (10.4–10.8) |
| | 2015 | 12.8 (12.5–13.1) | 15.9 (15.6–16.2) | 11.8 (11.6–12.1) | 11.0 (10.7–11.2) |
| | 2016 | 12.4 (12.1–12.7) | 16.1 (15.8–16.5) | 12.2 (11.9–12.5) | 11.3 (11.0–11.5) |
| | 2017 | 12.1 (11.9–12.4) | 16.4 (16.1–16.8) | 12.3 (12.1–12.6) | 11.6 (11.3–11.8) |
| | 2018 | 11.7 (11.5–12.0) | 16.5 (16.2–16.8) | 12.4 (12.1–12.7) | 12.0 (11.8–12.2) |
| | 2019 | 11.4 (11.2–11.7) | 16.4 (16.1–16.7) | 12.7 (12.4–12.9) | 12.4 (12.2–12.6) |
| Richmond | 2010 | 11.9 (11.6–12.3) | 8.7 (8.4–9.0) | 6.6 (6.4–6.8) | 7.5 (7.2–7.7) |
| | 2011 | 12.0 (11.6–12.3) | 8.9 (8.6–9.2) | 6.6 (6.4–6.8) | 7.6 (7.4–7.9) |
| | 2012 | 11.9 (11.6–12.2) | 9.3 (9.0–9.6) | 6.8 (6.6–7.1) | 7.9 (7.6–8.1) |
| | 2013 | 11.8 (11.5–12.2) | 9.8 (9.5–10.1) | 7.1 (6.9–7.4) | 8.1 (7.8–8.3) |
| | 2014 | 11.7 (11.3–12.0) | 10.1 (9.8–10.4) | 7.2 (7.0–7.5) | 8.3 (8.1–8.6) |
| | 2015 | 11.4 (11.1–11.8) | 10.4 (10.1–10.7) | 7.5 (7.2–7.7) | 8.6 (8.3–8.8) |
| | 2016 | 11.3 (10.9–11.6) | 10.5 (10.2–10.8) | 7.8 (7.5–8.1) | 8.8 (8.6–9.1) |
| | 2017 | 10.9 (10.6–11.3) | 10.5 (10.2–10.8) | 8.0 (7.7–8.2) | 9.1 (8.8–9.4) |
| | 2018 | 10.7 (10.3–11.0) | 10.6 (10.3–11.0) | 8.0 (7.8–8.3) | 9.2 (8.9–9.5) |
| | 2019 | 10.4 (10.1–10.7) | 10.7 (10.4–11.0) | 8.3 (8.1–8.6) | 9.4 (9.1–9.7) |
| South Vancouver Island | 2010 | 13.9 (13.7–14.2) | 13.3 (13.0–13.5) | 9.8 (9.6–10.1) | 9.4 (9.2–9.6) |
| | 2011 | 13.7 (13.5–14.0) | 13.9 (13.6–14.2) | 10.2 (9.9–10.4) | 9.6 (9.4–9.9) |
| | 2012 | 13.5 (13.2–13.8) | 14.4 (14.1–14.7) | 10.5 (10.3–10.8) | 10.0 (9.8–10.2) |
| | 2013 | 13.4 (13.1–13.7) | 14.8 (14.5–15.1) | 10.9 (10.7–11.2) | 10.3 (10.1–10.6) |
| | 2014 | 13.2 (12.9–13.5) | 15.0 (14.7–15.3) | 11.3 (11.0–11.5) | 10.6 (10.4–10.8) |
| | 2015 | 13.0 (12.7–13.3) | 15.4 (15.1–15.7) | 11.6 (11.3–11.8) | 11.0 (10.8–11.2) |
| | 2016 | 12.7 (12.5–13.0) | 15.7 (15.4–16.0) | 11.7 (11.4–11.9) | 11.4 (11.1–11.6) |
| | 2017 | 12.4 (12.1–12.7) | 15.7 (15.4–16.0) | 11.9 (11.7–12.2) | 11.6 (11.4–11.9) |

| | | Prevalence rate (%) | | | |
|---|---|---|---|---|---|
| **Health Service Delivery Area** | **Year** | **1-19** | **20-34** | **35-49** | **50-64** |
| | 2018 | 12.0 (11.7–12.3) | 15.9 (15.6–16.2) | 12.1 (11.9–12.4) | 11.9 (11.7–12.2) |
| | 2019 | 11.7 (11.5–12.0) | 15.8 (15.5–16.1) | 12.4 (12.1–12.6) | 12.3 (12.1–12.5) |
| Thompson Cariboo Shuswap | 2010 | 13.9 (13.6–14.2) | 13.9 (13.5–14.2) | 10.1 (9.8–10.4) | 10.3 (10.1–10.6) |
| | 2011 | 13.7 (13.4–14.1) | 14.6 (14.2–14.9) | 10.6 (10.3–10.9) | 10.5 (10.2–10.8) |
| | 2012 | 13.7 (13.4–14.1) | 15.1 (14.7–15.4) | 11.1 (10.8–11.5) | 10.8 (10.5–11.1) |
| | 2013 | 13.7 (13.4–14.1) | 15.6 (15.2–16.0) | 11.6 (11.3–11.9) | 11.2 (10.9–11.5) |
| | 2014 | 13.7 (13.4–14.1) | 16.1 (15.7–16.5) | 12.2 (11.8–12.5) | 11.5 (11.3–11.8) |
| | 2015 | 13.5 (13.1–13.8) | 16.4 (16.0–16.8) | 12.6 (12.3–13.0) | 11.8 (11.6–12.1) |
| | 2016 | 13.0 (12.7–13.4) | 16.8 (16.4–17.2) | 13.0 (12.6–13.3) | 12.1 (11.8–12.4) |
| | 2017 | 12.8 (12.5–13.1) | 17.1 (16.8–17.5) | 13.4 (13.0–13.7) | 12.5 (12.2–12.8) |
| | 2018 | 12.3 (11.9–12.6) | 17.1 (16.7–17.5) | 13.8 (13.5–14.2) | 12.9 (12.6–13.2) |
| | 2019 | 12.0 (11.7–12.3) | 17.3 (16.9–17.7) | 14.2 (13.9–14.6) | 13.3 (13.0–13.6) |
| Vancouver | 2010 | 14.1 (13.9–14.3) | 9.0 (8.8–9.1) | 8.1 (8.0–8.3) | 9.5 (9.3–9.7) |
| | 2011 | 13.9 (13.6–14.1) | 9.5 (9.3–9.6) | 8.3 (8.1–8.4) | 9.7 (9.5–9.8) |
| | 2012 | 13.7 (13.5–13.9) | 9.8 (9.6–9.9) | 8.5 (8.3–8.6) | 10.0 (9.8–10.2) |
| | 2013 | 13.4 (13.2–13.7) | 10.1 (9.9–10.2) | 8.8 (8.7–9.0) | 10.3 (10.2–10.5) |
| | 2014 | 13.0 (12.8–13.2) | 10.6 (10.4–10.7) | 9.0 (8.8–9.1) | 10.6 (10.4–10.7) |
| | 2015 | 12.6 (12.4–12.9) | 11.0 (10.9–11.2) | 9.0 (8.9–9.2) | 10.8 (10.7–11.0) |
| | 2016 | 12.2 (12.0–12.4) | 11.2 (11.0–11.3) | 9.3 (9.1–9.4) | 11.1 (10.9–11.3) |
| | 2017 | 11.7 (11.5–12.0) | 11.2 (11.1–11.4) | 9.3 (9.2–9.5) | 11.3 (11.1–11.5) |
| | 2018 | 11.3 (11.1–11.5) | 11.2 (11.0–11.3) | 9.5 (9.3–9.6) | 11.5 (11.3–11.7) |
| | 2019 | 10.9 (10.7–11.1) | 11.1 (10.9–11.2) | 9.6 (9.5–9.8) | 11.6 (11.5–11.8) |

**Appendix 4.** Health Service Delivery Area (HSDA) population size for the target population, corresponding to the starting year of the cost-effectiveness analysis (2010) and incidence projection analysis (2023)[51].

| | | Population | |
|---|---|---|---|
| **HSDA** | **Year** | **Age 5** | **Age 25** |
| **East Kootenay** | 2010 | 767 | 871 |
| | 2023 | 868 | 959 |
| **Kootenay Boundary** | 2010 | 735 | 669 |
| | 2023 | 688 | 845 |
| **Okanagan** | 2010 | 2929 | 4181 |
| | 2023 | 3691 | 5275 |
| **Thompson Cariboo Shuswap** | 2010 | 2096 | 2663 |
| | 2023 | 2334 | 3060 |
| **Fraser East** | 2010 | 3382 | 3795 |
| | 2023 | 3976 | 4515 |
| **Fraser North** | 2010 | 5860 | 9511 |
| | 2023 | 7127 | 11988 |
| **Fraser South** | 2010 | 8349 | 9995 |
| | 2023 | 9834 | 15177 |
| **Richmond** | 2010 | 1834 | 2938 |
| | 2023 | 2012 | 3894 |
| **Vancouver** | 2010 | 4726 | 13117 |
| | 2023 | 4933 | 15321 |
| **North Shore/Coast Garibaldi** | 2010 | 2534 | 3339 |
| | 2023 | 2825 | 3657 |
| **South Vancouver Island** | 2010 | 3033 | 5517 |
| | 2023 | 3638 | 6518 |
| **Central Vancouver Island** | 2010 | 2220 | 2747 |
| | 2023 | 2661 | 3173 |
| **North Vancouver Island** | 2010 | 1146 | 1156 |
| | 2023 | 1217 | 1199 |
| **Northwest** | 2010 | 884 | 792 |
| | 2023 | 845 | 946 |
| **Northern Interior** | 2010 | 1643 | 1886 |
| | 2023 | 1631 | 2026 |
| **Northeast** | 2010 | 877 | 1184 |
| | 2023 | 978 | 943 |

A) 0% increase in wildfire PM2.5 from 2023-2036.

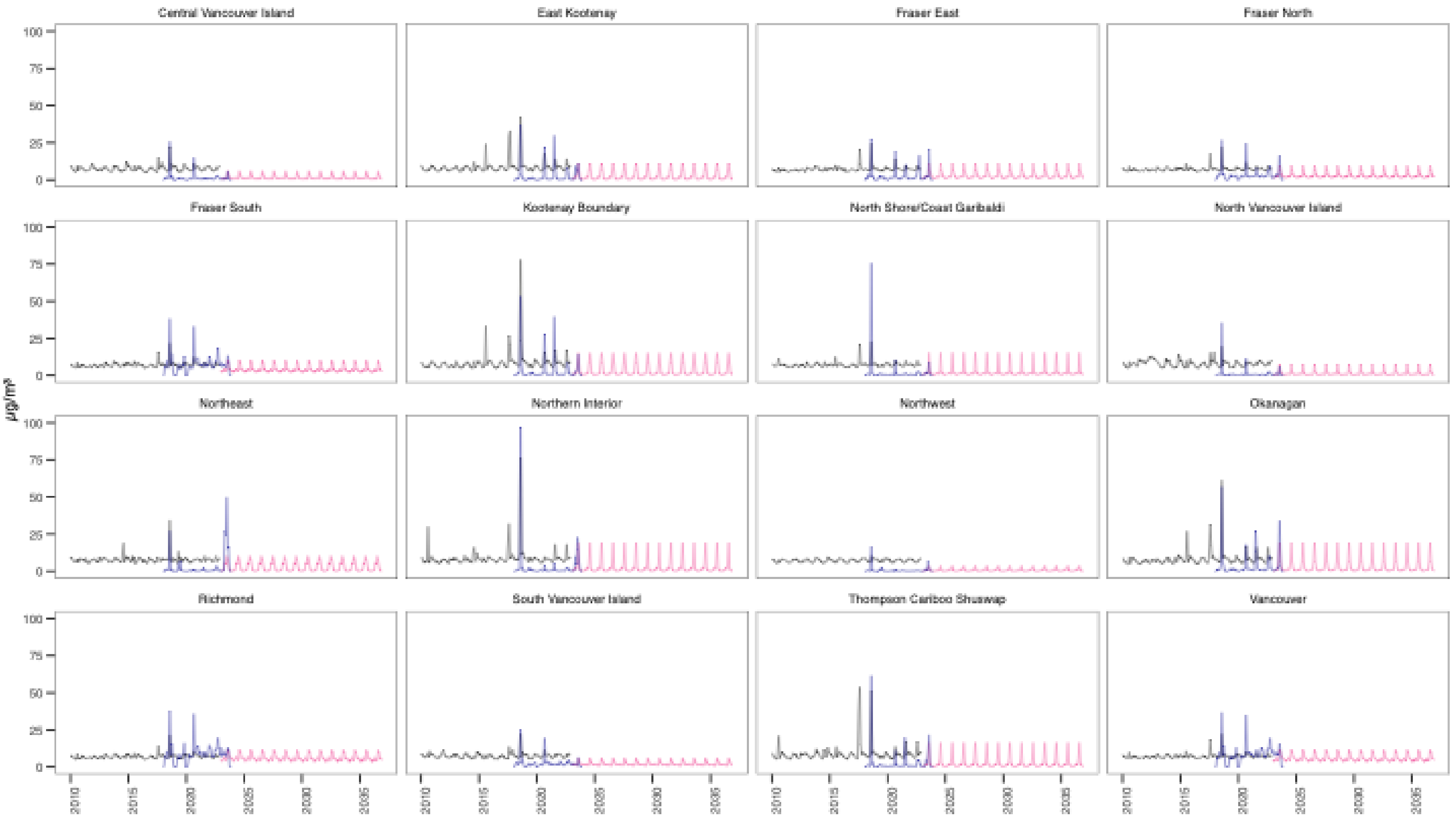


— CanOSSEM (2010-2022); — RAQDPS-FW (2018-2022); — projections (2023-2036)

B) 5.5% increase in wildfire $PM_{2.5}$ from 2023-2036.

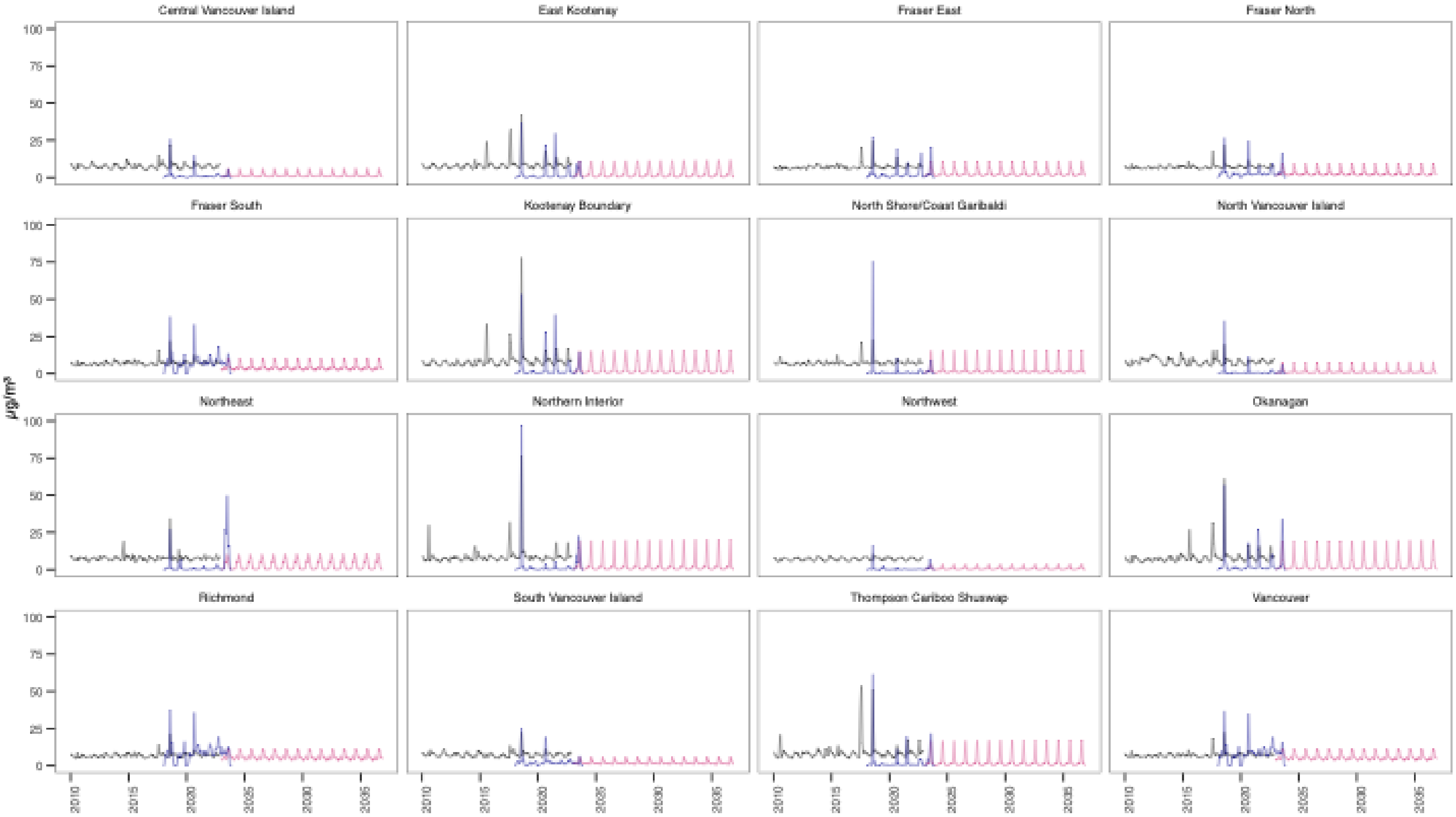


— CanOSSEM (2010-2022); — RAQDPS-FW (2018-2022); — projections (2023-2036)

C) 11.0% increase in wildfire $PM_{2.5}$ from 2023-2036.

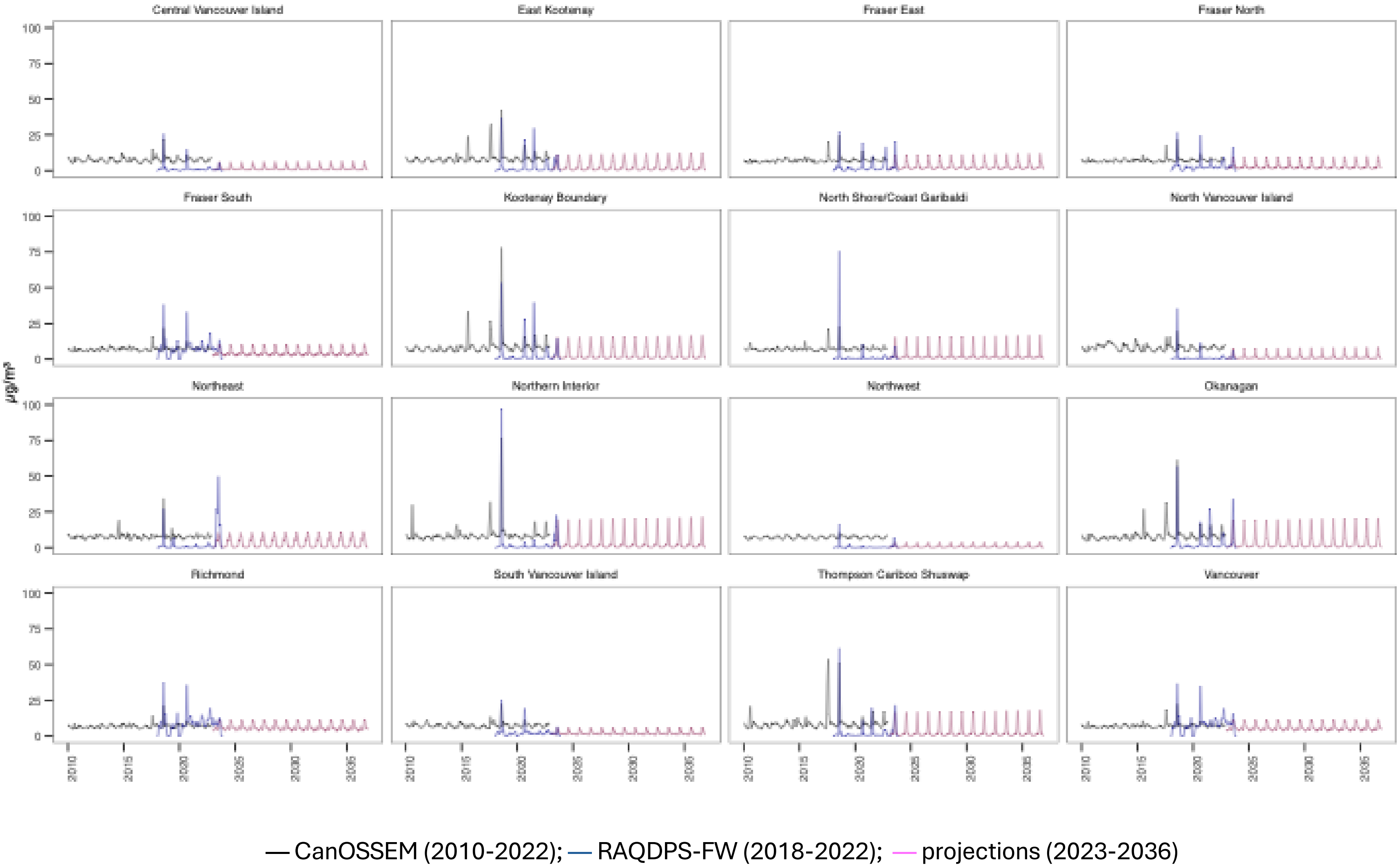

A) Counterfactual no wildfire $PM_{2.5}$ from 2023-2036. Black = CanOSSEM (2010-2022) and GEM-MACH (2023-2036).

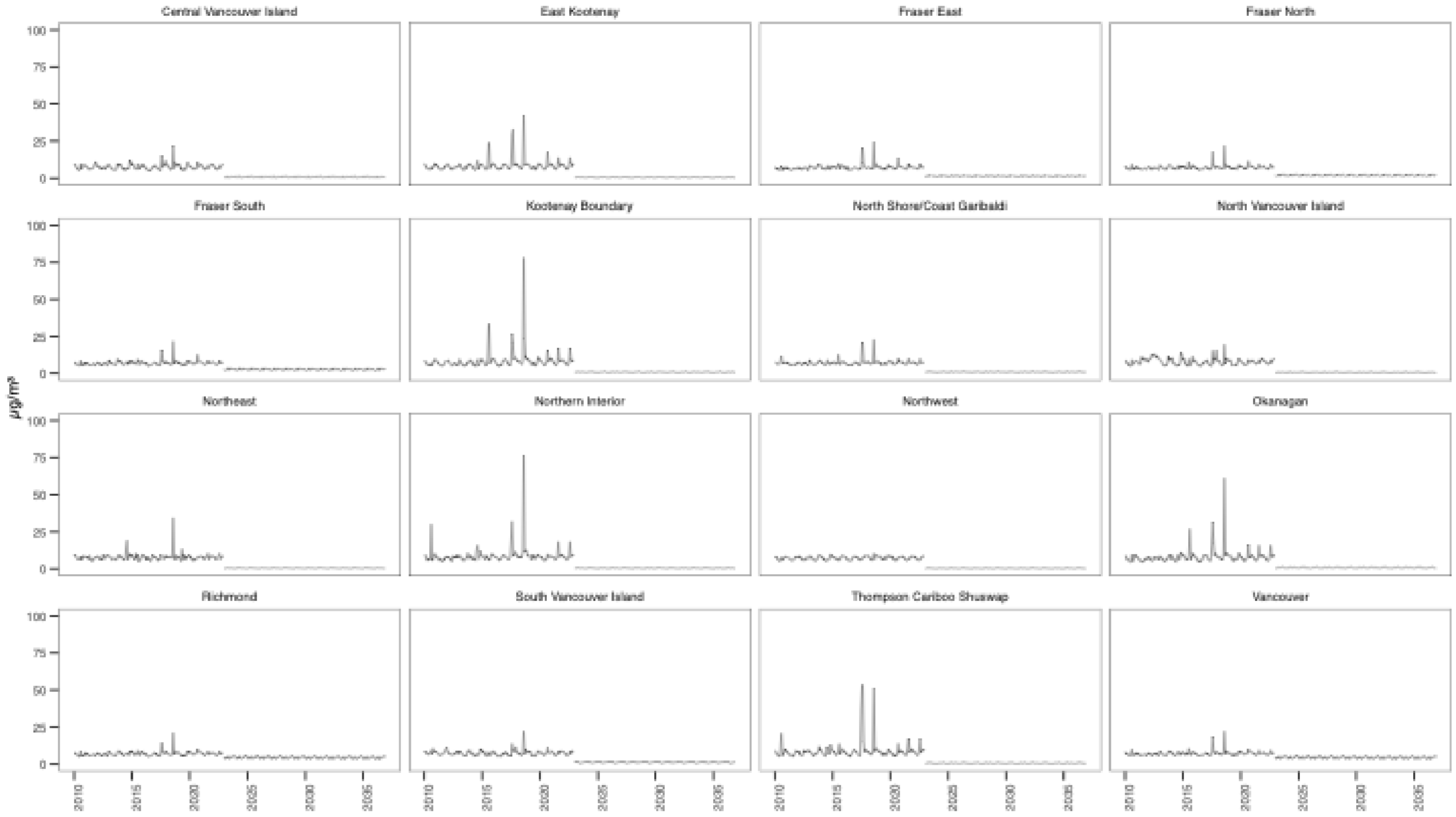


**Appendix 5.** $PM_{2.5}$ exposure by heath service delivery area (HSDA) in the historical (2010-2022, CanOSSEM) and projected (2023-2036) period. A) Assuming 0% increasing in wildfire $PM_{2.5}$ from 2023-2036; B) Assuming 5.5% increasing in wildfire $PM_{2.5}$ from 2023-2036; C) Assuming 11.0% increasing in wildfire $PM_{2.5}$ from 2023-2036; D) Counterfactual no wildfire $PM_{2.5}$ from 2023-2036.

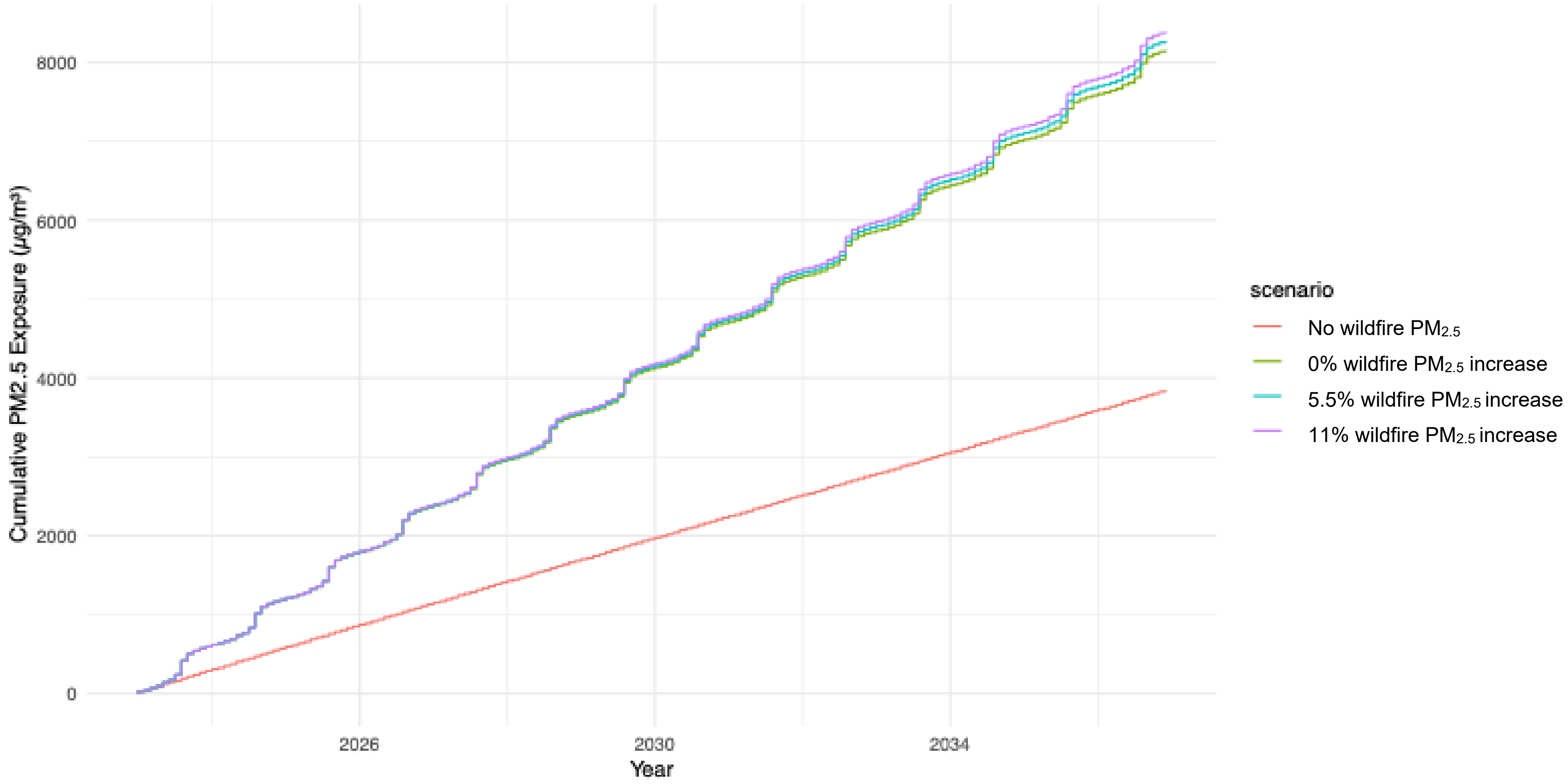


**Appendix 6.** Cumulative sum of monthly $PM_{2.5}$ exposure by projection scenario, averaged across all HSDAs in BC.

**Appendix 7.** Deterministic sensitivity analysis results by Health Service Delivery Area (HSDA). ICERs are shown for each HSDA and cohort (C = child; A = adult) and impact of varying key model parameters from their base-case values.

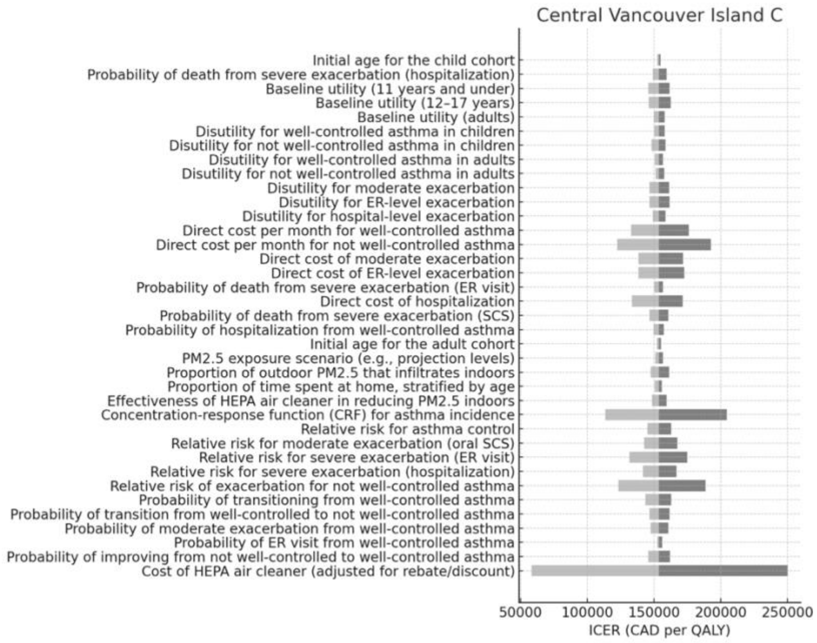


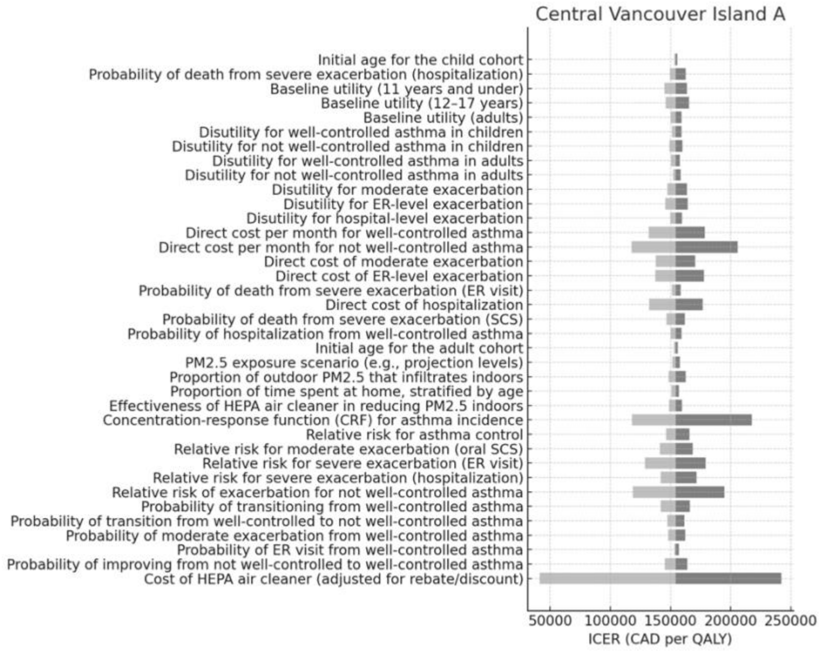

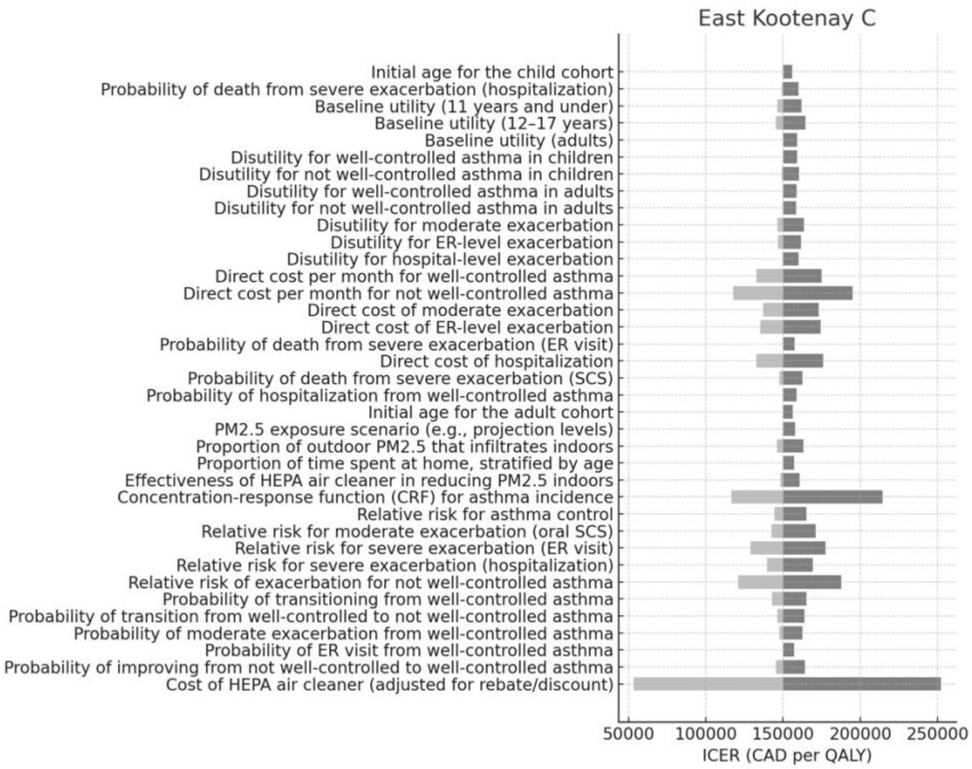
East Kootenay C
Initial age for the child cohort
Probability of death from severe exacerbation (hospitalization)
Baseline utility (11 years and under)
Baseline utility (12–17 years)
Baseline utility (adults)
Disutility for well-controlled asthma in children
Disutility for not well-controlled asthma in children
Disutility for well-controlled asthma in adults
Disutility for not well-controlled asthma in adults
Disutility for moderate exacerbation
Disutility for ER-level exacerbation
Disutility for hospital-level exacerbation
Direct cost per month for well-controlled asthma
Direct cost per month for not well-controlled asthma
Direct cost of moderate exacerbation
Direct cost of ER-level exacerbation
Probability of death from severe exacerbation (ER visit)
Direct cost of hospitalization
Probability of death from severe exacerbation (SCS)
Probability of hospitalization from well-controlled asthma
Initial age for the adult cohort
PM2.5 exposure scenario (e.g., projection levels)
Proportion of outdoor PM2.5 that infiltrates indoors
Proportion of time spent at home, stratified by age
Effectiveness of HEPA air cleaner in reducing PM2.5 indoors
Concentration-response function (CRF) for asthma incidence
Relative risk for asthma control
Relative risk for moderate exacerbation (oral SCS)
Relative risk for severe exacerbation (ER visit)
Relative risk for severe exacerbation (hospitalization)
Relative risk of exacerbation for not well-controlled asthma
Probability of transitioning from well-controlled asthma
Probability of transition from well-controlled to not well-controlled asthma
Probability of moderate exacerbation from well-controlled asthma
Probability of ER visit from well-controlled asthma
Probability of improving from not well-controlled to well-controlled asthma
Cost of HEPA air cleaner (adjusted for rebate/discount)
50000
100000
150000
200000
250000
ICER (CAD per QALY)

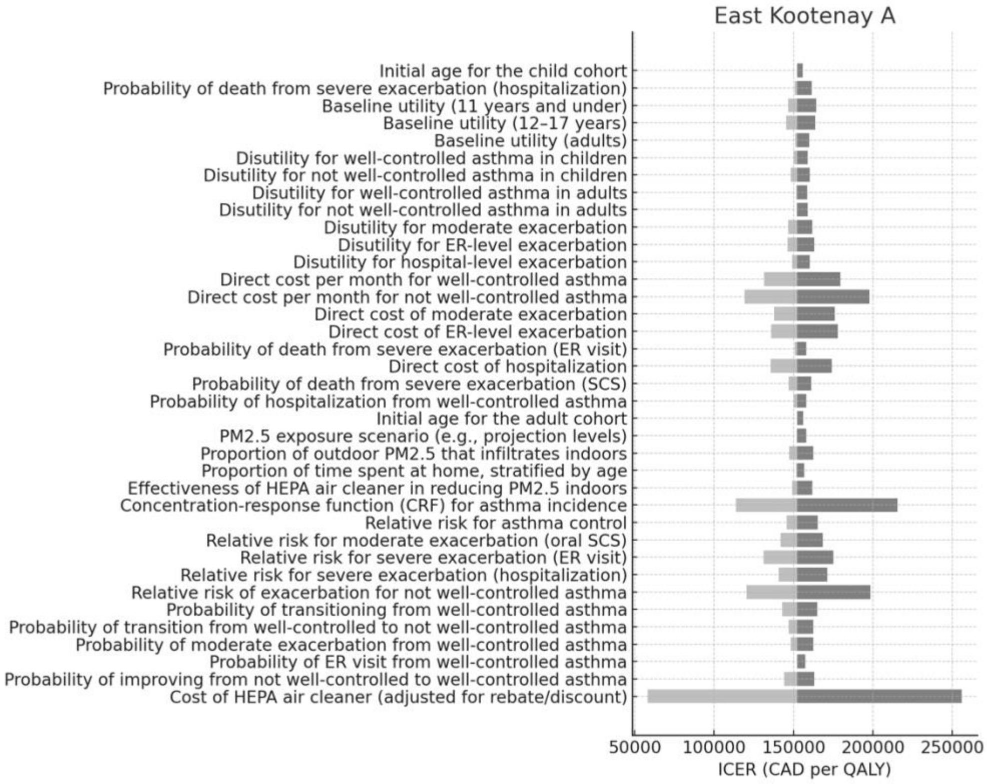
East Kootenay A
Initial age for the child cohort
Probability of death from severe exacerbation (hospitalization)
Baseline utility (11 years and under)
Baseline utility (12–17 years)
Baseline utility (adults)
Disutility for well-controlled asthma in children
Disutility for not well-controlled asthma in children
Disutility for well-controlled asthma in adults
Disutility for not well-controlled asthma in adults
Disutility for moderate exacerbation
Disutility for ER-level exacerbation
Disutility for hospital-level exacerbation
Direct cost per month for well-controlled asthma
Direct cost per month for not well-controlled asthma
Direct cost of moderate exacerbation
Direct cost of ER-level exacerbation
Probability of death from severe exacerbation (ER visit)
Direct cost of hospitalization
Probability of death from severe exacerbation (SCS)
Probability of hospitalization from well-controlled asthma
Initial age for the adult cohort
PM2.5 exposure scenario (e.g., projection levels)
Proportion of outdoor PM2.5 that infiltrates indoors
Proportion of time spent at home, stratified by age
Effectiveness of HEPA air cleaner in reducing PM2.5 indoors
Concentration-response function (CRF) for asthma incidence
Relative risk for asthma control
Relative risk for moderate exacerbation (oral SCS)
Relative risk for severe exacerbation (ER visit)
Relative risk for severe exacerbation (hospitalization)
Relative risk of exacerbation for not well-controlled asthma
Probability of transitioning from well-controlled asthma
Probability of transition from well-controlled to not well-controlled asthma
Probability of moderate exacerbation from well-controlled asthma
Probability of ER visit from well-controlled asthma
Probability of improving from not well-controlled to well-controlled asthma
Cost of HEPA air cleaner (adjusted for rebate/discount)
50000
100000
150000
200000
250000
ICER (CAD per QALY)

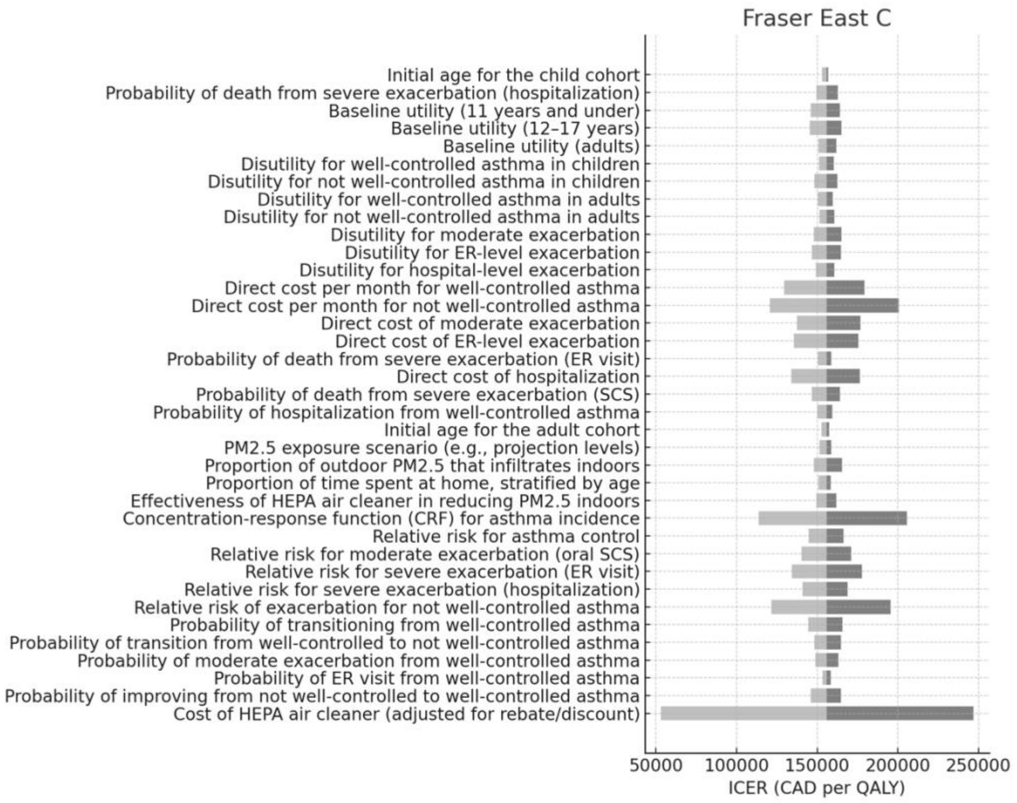
Fraser East C
Initial age for the child cohort
Probability of death from severe exacerbation (hospitalization)
Baseline utility (11 years and under)
Baseline utility (12–17 years)
Baseline utility (adults)
Disutility for well-controlled asthma in children
Disutility for not well-controlled asthma in children
Disutility for well-controlled asthma in adults
Disutility for not well-controlled asthma in adults
Disutility for moderate exacerbation
Disutility for ER-level exacerbation
Disutility for hospital-level exacerbation
Direct cost per month for well-controlled asthma
Direct cost per month for not well-controlled asthma
Direct cost of moderate exacerbation
Direct cost of ER-level exacerbation
Probability of death from severe exacerbation (ER visit)
Direct cost of hospitalization
Probability of death from severe exacerbation (SCS)
Probability of hospitalization from well-controlled asthma
Initial age for the adult cohort
PM2.5 exposure scenario (e.g., projection levels)
Proportion of outdoor PM2.5 that infiltrates indoors
Proportion of time spent at home, stratified by age
Effectiveness of HEPA air cleaner in reducing PM2.5 indoors
Concentration-response function (CRF) for asthma incidence
Relative risk for asthma control
Relative risk for moderate exacerbation (oral SCS)
Relative risk for severe exacerbation (ER visit)
Relative risk for severe exacerbation (hospitalization)
Relative risk of exacerbation for not well-controlled asthma
Probability of transitioning from well-controlled asthma
Probability of transition from well-controlled to not well-controlled asthma
Probability of moderate exacerbation from well-controlled asthma
Probability of ER visit from well-controlled asthma
Probability of improving from not well-controlled to well-controlled asthma
Cost of HEPA air cleaner (adjusted for rebate/discount)
50000
100000
150000
200000
250000
ICER (CAD per QALY)

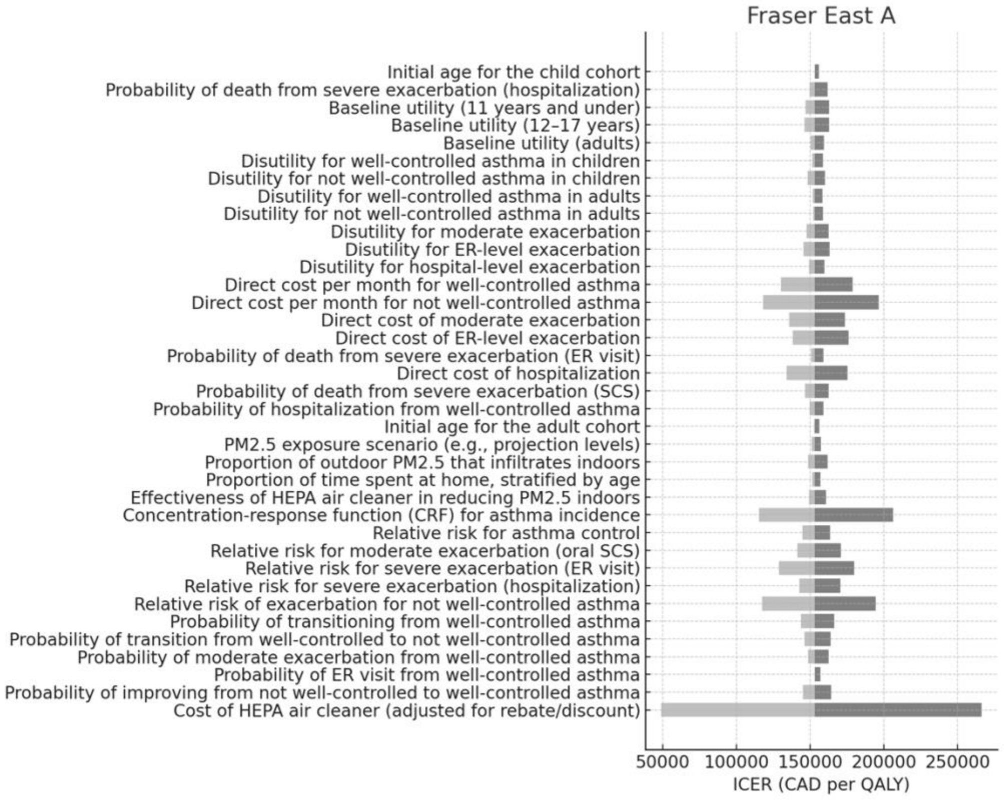
Fraser East A
Initial age for the child cohort
Probability of death from severe exacerbation (hospitalization)
Baseline utility (11 years and under)
Baseline utility (12–17 years)
Baseline utility (adults)
Disutility for well-controlled asthma in children
Disutility for not well-controlled asthma in children
Disutility for well-controlled asthma in adults
Disutility for not well-controlled asthma in adults
Disutility for moderate exacerbation
Disutility for ER-level exacerbation
Disutility for hospital-level exacerbation
Direct cost per month for well-controlled asthma
Direct cost per month for not well-controlled asthma
Direct cost of moderate exacerbation
Direct cost of ER-level exacerbation
Probability of death from severe exacerbation (ER visit)
Direct cost of hospitalization
Probability of death from severe exacerbation (SCS)
Probability of hospitalization from well-controlled asthma
Initial age for the adult cohort
PM2.5 exposure scenario (e.g., projection levels)
Proportion of outdoor PM2.5 that infiltrates indoors
Proportion of time spent at home, stratified by age
Effectiveness of HEPA air cleaner in reducing PM2.5 indoors
Concentration-response function (CRF) for asthma incidence
Relative risk for asthma control
Relative risk for moderate exacerbation (oral SCS)
Relative risk for severe exacerbation (ER visit)
Relative risk for severe exacerbation (hospitalization)
Relative risk of exacerbation for not well-controlled asthma
Probability of transitioning from well-controlled asthma
Probability of transition from well-controlled to not well-controlled asthma
Probability of moderate exacerbation from well-controlled asthma
Probability of ER visit from well-controlled asthma
Probability of improving from not well-controlled to well-controlled asthma
Cost of HEPA air cleaner (adjusted for rebate/discount)
50000
100000
150000
200000
250000
ICER (CAD per QALY)

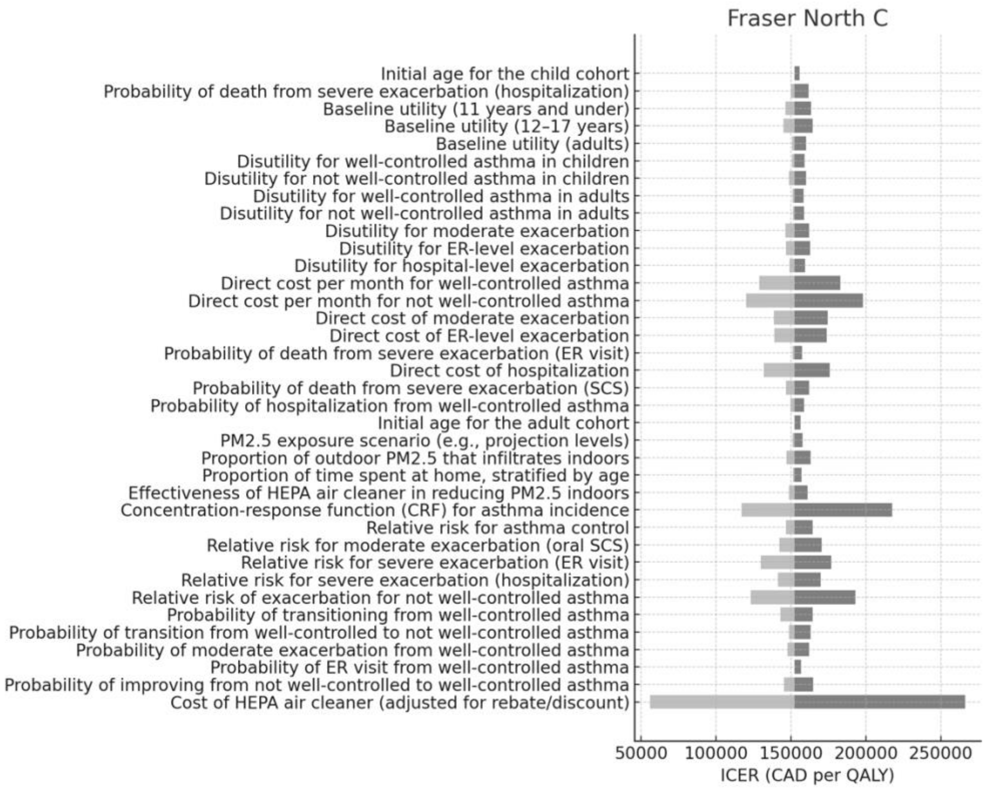
Fraser North C
Initial age for the child cohort
Probability of death from severe exacerbation (hospitalization)
Baseline utility (11 years and under)
Baseline utility (12–17 years)
Baseline utility (adults)
Disutility for well-controlled asthma in children
Disutility for not well-controlled asthma in children
Disutility for well-controlled asthma in adults
Disutility for not well-controlled asthma in adults
Disutility for moderate exacerbation
Disutility for ER-level exacerbation
Disutility for hospital-level exacerbation
Direct cost per month for well-controlled asthma
Direct cost per month for not well-controlled asthma
Direct cost of moderate exacerbation
Direct cost of ER-level exacerbation
Probability of death from severe exacerbation (ER visit)
Direct cost of hospitalization
Probability of death from severe exacerbation (SCS)
Probability of hospitalization from well-controlled asthma
Initial age for the adult cohort
PM2.5 exposure scenario (e.g., projection levels)
Proportion of outdoor PM2.5 that infiltrates indoors
Proportion of time spent at home, stratified by age
Effectiveness of HEPA air cleaner in reducing PM2.5 indoors
Concentration-response function (CRF) for asthma incidence
Relative risk for asthma control
Relative risk for moderate exacerbation (oral SCS)
Relative risk for severe exacerbation (ER visit)
Relative risk for severe exacerbation (hospitalization)
Relative risk of exacerbation for not well-controlled asthma
Probability of transitioning from well-controlled asthma
Probability of transition from well-controlled to not well-controlled asthma
Probability of moderate exacerbation from well-controlled asthma
Probability of ER visit from well-controlled asthma
Probability of improving from not well-controlled to well-controlled asthma
Cost of HEPA air cleaner (adjusted for rebate/discount)
50000
100000
150000
200000
250000
ICER (CAD per QALY)

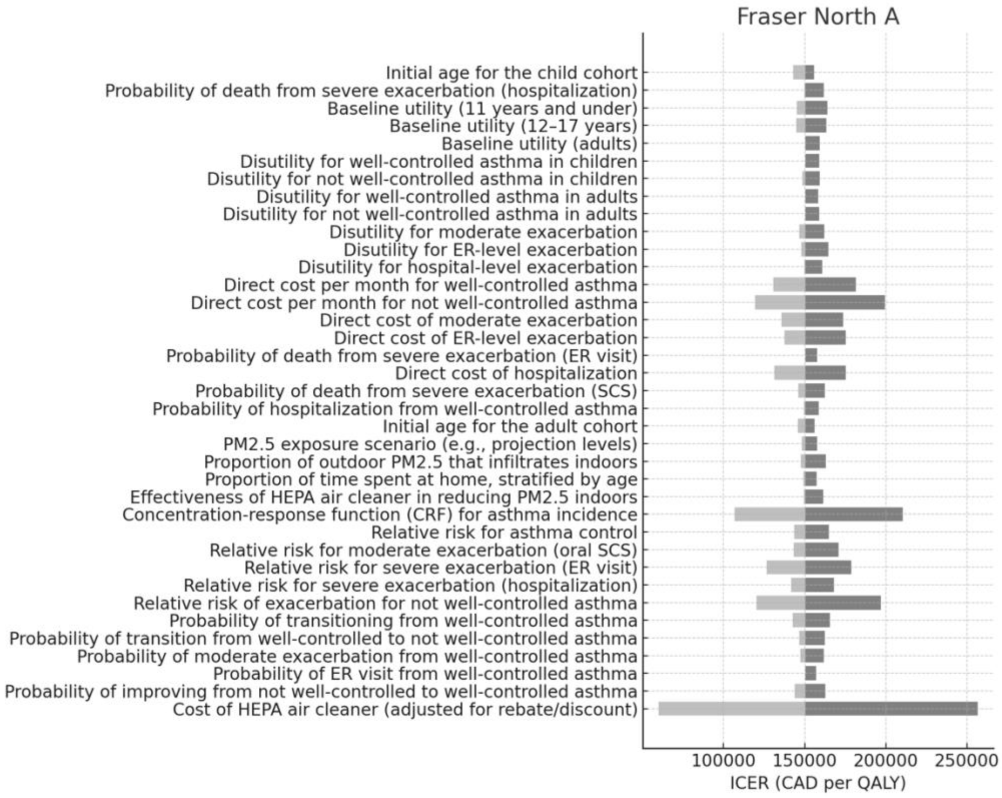
Fraser North A
Initial age for the child cohort
Probability of death from severe exacerbation (hospitalization)
Baseline utility (11 years and under)
Baseline utility (12–17 years)
Baseline utility (adults)
Disutility for well-controlled asthma in children
Disutility for not well-controlled asthma in children
Disutility for well-controlled asthma in adults
Disutility for not well-controlled asthma in adults
Disutility for moderate exacerbation
Disutility for ER-level exacerbation
Disutility for hospital-level exacerbation
Direct cost per month for well-controlled asthma
Direct cost per month for not well-controlled asthma
Direct cost of moderate exacerbation
Direct cost of ER-level exacerbation
Probability of death from severe exacerbation (ER visit)
Direct cost of hospitalization
Probability of death from severe exacerbation (SCS)
Probability of hospitalization from well-controlled asthma
Initial age for the adult cohort
PM2.5 exposure scenario (e.g., projection levels)
Proportion of outdoor PM2.5 that infiltrates indoors
Proportion of time spent at home, stratified by age
Effectiveness of HEPA air cleaner in reducing PM2.5 indoors
Concentration-response function (CRF) for asthma incidence
Relative risk for asthma control
Relative risk for moderate exacerbation (oral SCS)
Relative risk for severe exacerbation (ER visit)
Relative risk for severe exacerbation (hospitalization)
Relative risk of exacerbation for not well-controlled asthma
Probability of transitioning from well-controlled asthma
Probability of transition from well-controlled to not well-controlled asthma
Probability of moderate exacerbation from well-controlled asthma
Probability of ER visit from well-controlled asthma
Probability of improving from not well-controlled to well-controlled asthma
Cost of HEPA air cleaner (adjusted for rebate/discount)
100000
150000
200000
250000
ICER (CAD per QALY)

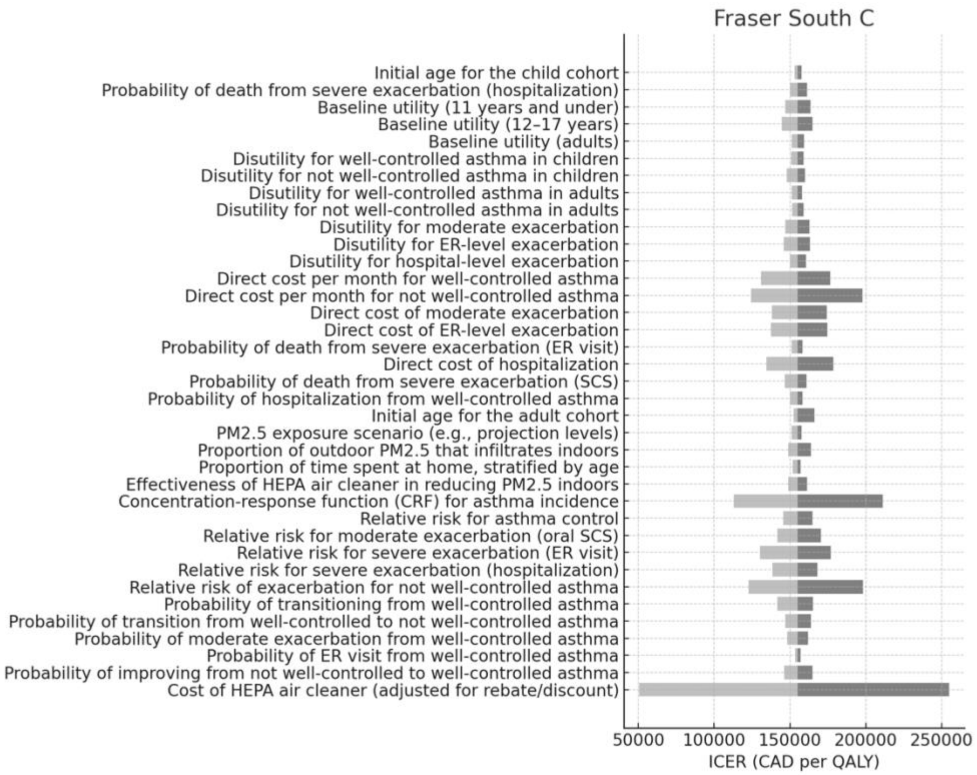
Fraser South C
Initial age for the child cohort
Probability of death from severe exacerbation (hospitalization)
Baseline utility (11 years and under)
Baseline utility (12–17 years)
Baseline utility (adults)
Disutility for well-controlled asthma in children
Disutility for not well-controlled asthma in children
Disutility for well-controlled asthma in adults
Disutility for not well-controlled asthma in adults
Disutility for moderate exacerbation
Disutility for ER-level exacerbation
Disutility for hospital-level exacerbation
Direct cost per month for well-controlled asthma
Direct cost per month for not well-controlled asthma
Direct cost of moderate exacerbation
Direct cost of ER-level exacerbation
Probability of death from severe exacerbation (ER visit)
Direct cost of hospitalization
Probability of death from severe exacerbation (SCS)
Probability of hospitalization from well-controlled asthma
Initial age for the adult cohort
PM2.5 exposure scenario (e.g., projection levels)
Proportion of outdoor PM2.5 that infiltrates indoors
Proportion of time spent at home, stratified by age
Effectiveness of HEPA air cleaner in reducing PM2.5 indoors
Concentration-response function (CRF) for asthma incidence
Relative risk for asthma control
Relative risk for moderate exacerbation (oral SCS)
Relative risk for severe exacerbation (ER visit)
Relative risk for severe exacerbation (hospitalization)
Relative risk of exacerbation for not well-controlled asthma
Probability of transitioning from well-controlled asthma
Probability of transition from well-controlled to not well-controlled asthma
Probability of moderate exacerbation from well-controlled asthma
Probability of ER visit from well-controlled asthma
Probability of improving from not well-controlled to well-controlled asthma
Cost of HEPA air cleaner (adjusted for rebate/discount)
50000
100000
150000
200000
250000
ICER (CAD per QALY)

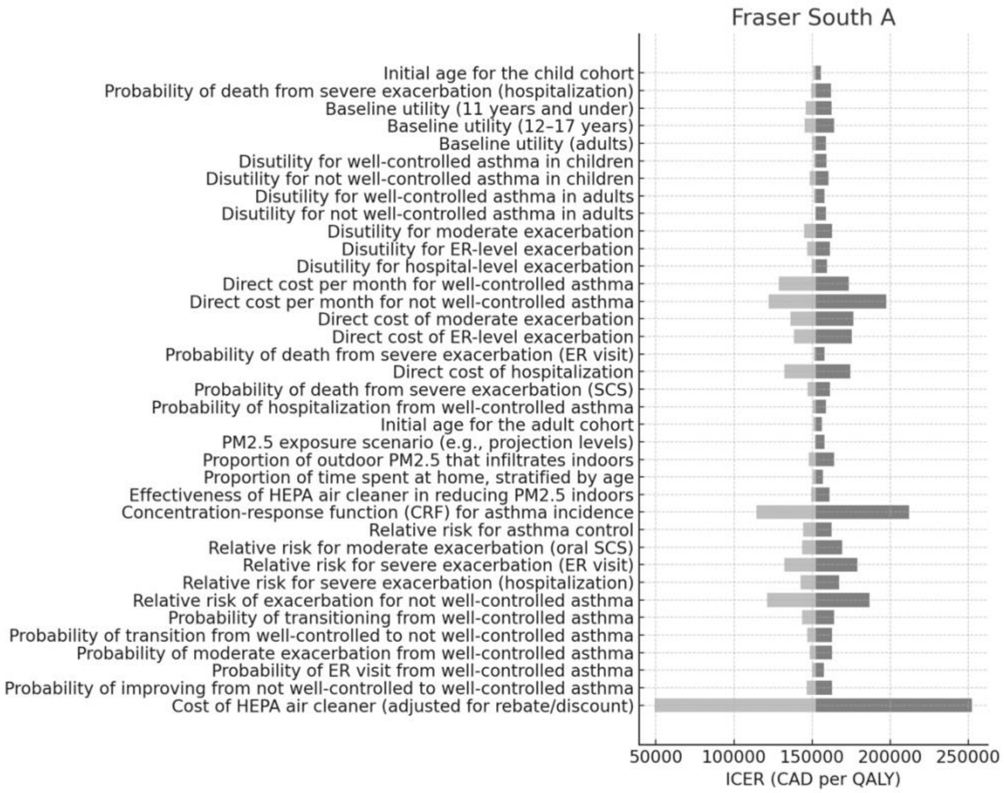
Fraser South A
Initial age for the child cohort
Probability of death from severe exacerbation (hospitalization)
Baseline utility (11 years and under)
Baseline utility (12–17 years)
Baseline utility (adults)
Disutility for well-controlled asthma in children
Disutility for not well-controlled asthma in children
Disutility for well-controlled asthma in adults
Disutility for not well-controlled asthma in adults
Disutility for moderate exacerbation
Disutility for ER-level exacerbation
Disutility for hospital-level exacerbation
Direct cost per month for well-controlled asthma
Direct cost per month for not well-controlled asthma
Direct cost of moderate exacerbation
Direct cost of ER-level exacerbation
Probability of death from severe exacerbation (ER visit)
Direct cost of hospitalization
Probability of death from severe exacerbation (SCS)
Probability of hospitalization from well-controlled asthma
Initial age for the adult cohort
PM2.5 exposure scenario (e.g., projection levels)
Proportion of outdoor PM2.5 that infiltrates indoors
Proportion of time spent at home, stratified by age
Effectiveness of HEPA air cleaner in reducing PM2.5 indoors
Concentration-response function (CRF) for asthma incidence
Relative risk for asthma control
Relative risk for moderate exacerbation (oral SCS)
Relative risk for severe exacerbation (ER visit)
Relative risk for severe exacerbation (hospitalization)
Relative risk of exacerbation for not well-controlled asthma
Probability of transitioning from well-controlled asthma
Probability of transition from well-controlled to not well-controlled asthma
Probability of moderate exacerbation from well-controlled asthma
Probability of ER visit from well-controlled asthma
Probability of improving from not well-controlled to well-controlled asthma
Cost of HEPA air cleaner (adjusted for rebate/discount)
50000
100000
150000
200000
250000
ICER (CAD per QALY)

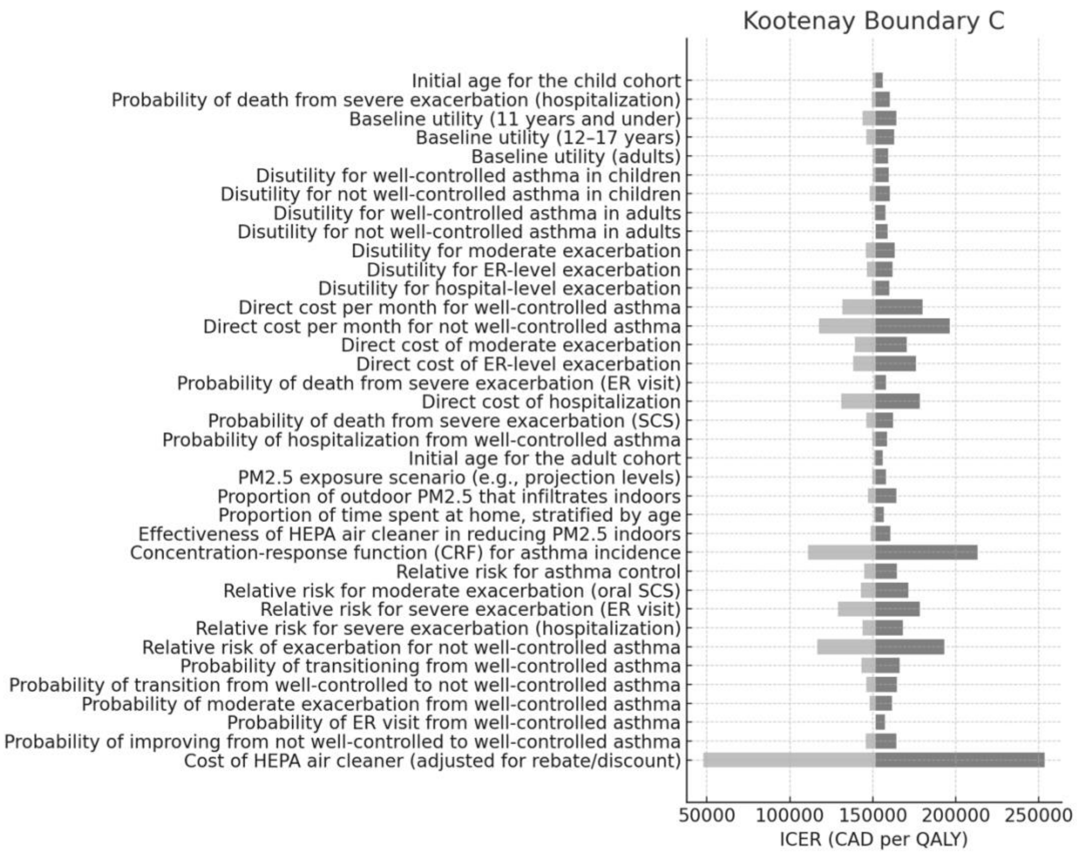
Kootenay Boundary C
Initial age for the child cohort
Probability of death from severe exacerbation (hospitalization)
Baseline utility (11 years and under)
Baseline utility (12–17 years)
Baseline utility (adults)
Disutility for well-controlled asthma in children
Disutility for not well-controlled asthma in children
Disutility for well-controlled asthma in adults
Disutility for not well-controlled asthma in adults
Disutility for moderate exacerbation
Disutility for ER-level exacerbation
Disutility for hospital-level exacerbation
Direct cost per month for well-controlled asthma
Direct cost per month for not well-controlled asthma
Direct cost of moderate exacerbation
Direct cost of ER-level exacerbation
Probability of death from severe exacerbation (ER visit)
Direct cost of hospitalization
Probability of death from severe exacerbation (SCS)
Probability of hospitalization from well-controlled asthma
Initial age for the adult cohort
PM2.5 exposure scenario (e.g., projection levels)
Proportion of outdoor PM2.5 that infiltrates indoors
Proportion of time spent at home, stratified by age
Effectiveness of HEPA air cleaner in reducing PM2.5 indoors
Concentration-response function (CRF) for asthma incidence
Relative risk for asthma control
Relative risk for moderate exacerbation (oral SCS)
Relative risk for severe exacerbation (ER visit)
Relative risk for severe exacerbation (hospitalization)
Relative risk of exacerbation for not well-controlled asthma
Probability of transitioning from well-controlled asthma
Probability of transition from well-controlled to not well-controlled asthma
Probability of moderate exacerbation from well-controlled asthma
Probability of ER visit from well-controlled asthma
Probability of improving from not well-controlled to well-controlled asthma
Cost of HEPA air cleaner (adjusted for rebate/discount)
50000
100000
150000
200000
250000
ICER (CAD per QALY)

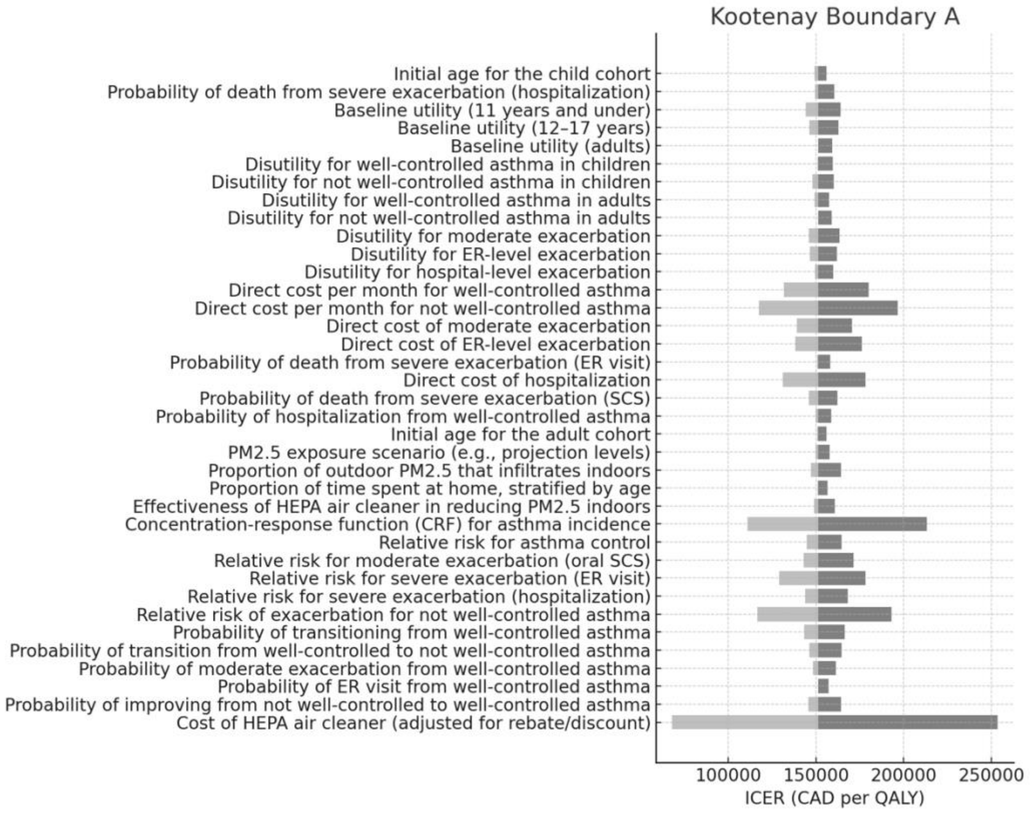
Kootenay Boundary A
Initial age for the child cohort
Probability of death from severe exacerbation (hospitalization)
Baseline utility (11 years and under)
Baseline utility (12–17 years)
Baseline utility (adults)
Disutility for well-controlled asthma in children
Disutility for not well-controlled asthma in children
Disutility for well-controlled asthma in adults
Disutility for not well-controlled asthma in adults
Disutility for moderate exacerbation
Disutility for ER-level exacerbation
Disutility for hospital-level exacerbation
Direct cost per month for well-controlled asthma
Direct cost per month for not well-controlled asthma
Direct cost of moderate exacerbation
Direct cost of ER-level exacerbation
Probability of death from severe exacerbation (ER visit)
Direct cost of hospitalization
Probability of death from severe exacerbation (SCS)
Probability of hospitalization from well-controlled asthma
Initial age for the adult cohort
PM2.5 exposure scenario (e.g., projection levels)
Proportion of outdoor PM2.5 that infiltrates indoors
Proportion of time spent at home, stratified by age
Effectiveness of HEPA air cleaner in reducing PM2.5 indoors
Concentration-response function (CRF) for asthma incidence
Relative risk for asthma control
Relative risk for moderate exacerbation (oral SCS)
Relative risk for severe exacerbation (ER visit)
Relative risk for severe exacerbation (hospitalization)
Relative risk of exacerbation for not well-controlled asthma
Probability of transitioning from well-controlled asthma
Probability of transition from well-controlled to not well-controlled asthma
Probability of moderate exacerbation from well-controlled asthma
Probability of ER visit from well-controlled asthma
Probability of improving from not well-controlled to well-controlled asthma
Cost of HEPA air cleaner (adjusted for rebate/discount)
100000
150000
200000
250000
ICER (CAD per QALY)

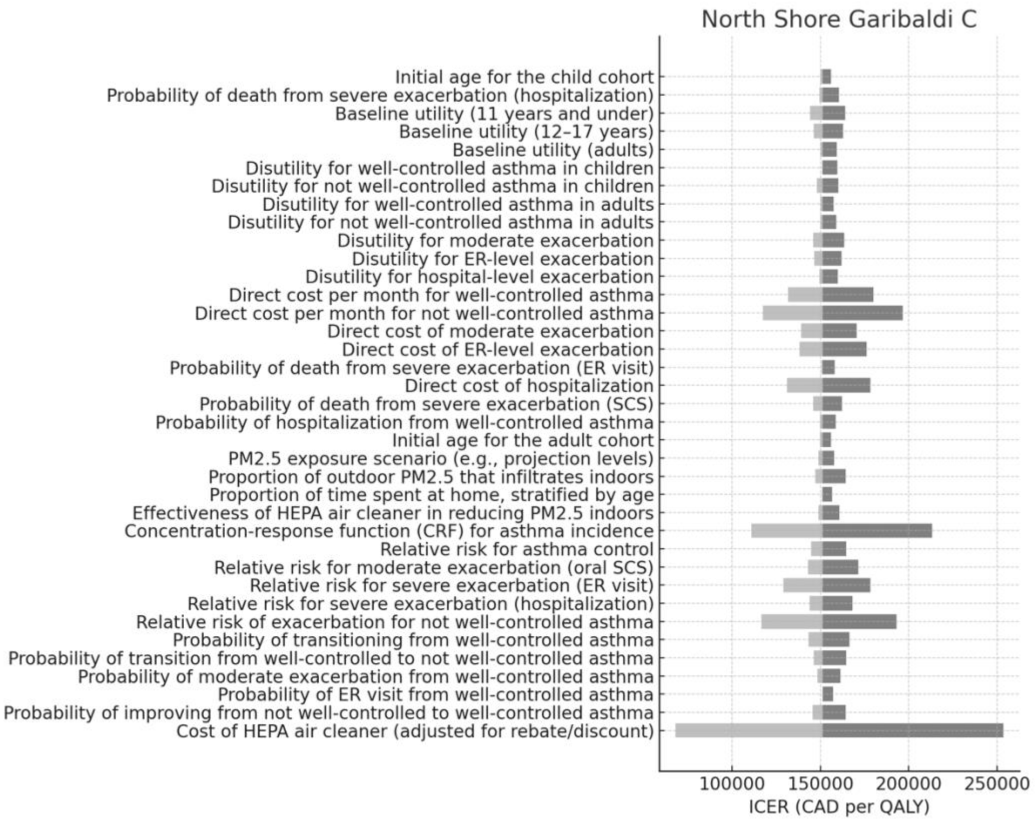
North Shore Garibaldi C
Initial age for the child cohort
Probability of death from severe exacerbation (hospitalization)
Baseline utility (11 years and under)
Baseline utility (12–17 years)
Baseline utility (adults)
Disutility for well-controlled asthma in children
Disutility for not well-controlled asthma in children
Disutility for well-controlled asthma in adults
Disutility for not well-controlled asthma in adults
Disutility for moderate exacerbation
Disutility for ER-level exacerbation
Disutility for hospital-level exacerbation
Direct cost per month for well-controlled asthma
Direct cost per month for not well-controlled asthma
Direct cost of moderate exacerbation
Direct cost of ER-level exacerbation
Probability of death from severe exacerbation (ER visit)
Direct cost of hospitalization
Probability of death from severe exacerbation (SCS)
Probability of hospitalization from well-controlled asthma
Initial age for the adult cohort
PM2.5 exposure scenario (e.g., projection levels)
Proportion of outdoor PM2.5 that infiltrates indoors
Proportion of time spent at home, stratified by age
Effectiveness of HEPA air cleaner in reducing PM2.5 indoors
Concentration-response function (CRF) for asthma incidence
Relative risk for asthma control
Relative risk for moderate exacerbation (oral SCS)
Relative risk for severe exacerbation (ER visit)
Relative risk for severe exacerbation (hospitalization)
Relative risk of exacerbation for not well-controlled asthma
Probability of transitioning from well-controlled asthma
Probability of transition from well-controlled to not well-controlled asthma
Probability of moderate exacerbation from well-controlled asthma
Probability of ER visit from well-controlled asthma
Probability of improving from not well-controlled to well-controlled asthma
Cost of HEPA air cleaner (adjusted for rebate/discount)
100000
150000
200000
250000
ICER (CAD per QALY)

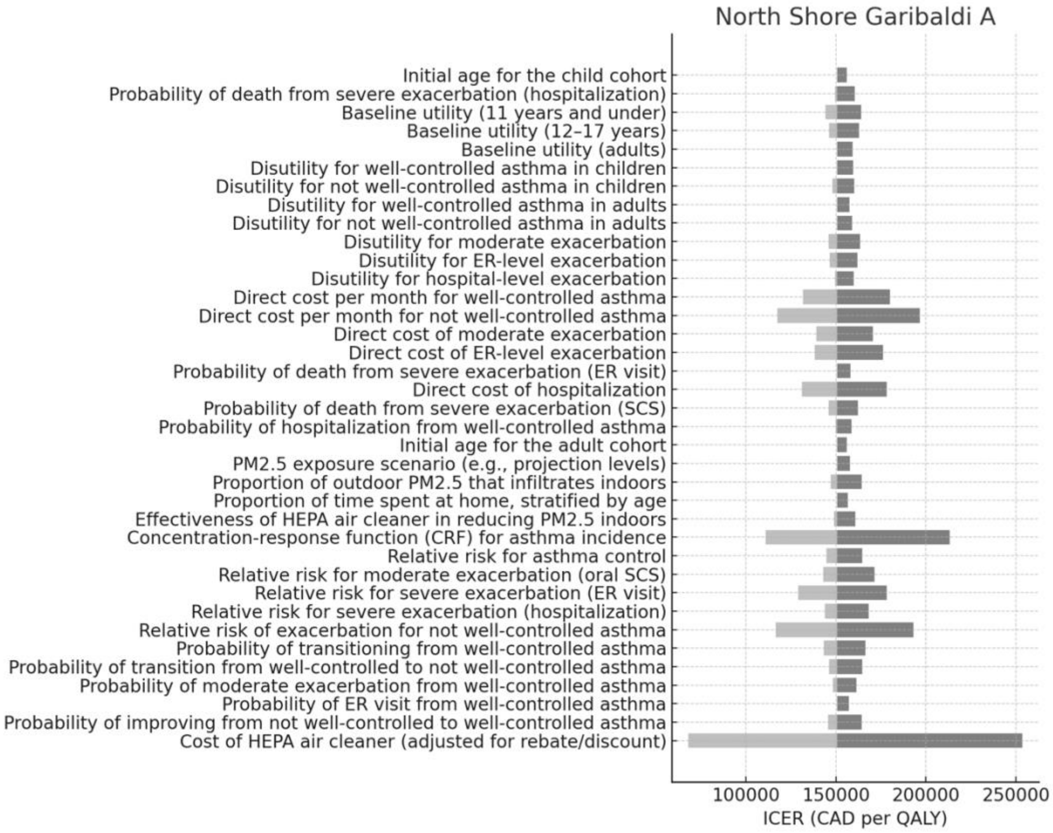
North Shore Garibaldi A
Initial age for the child cohort
Probability of death from severe exacerbation (hospitalization)
Baseline utility (11 years and under)
Baseline utility (12–17 years)
Baseline utility (adults)
Disutility for well-controlled asthma in children
Disutility for not well-controlled asthma in children
Disutility for well-controlled asthma in adults
Disutility for not well-controlled asthma in adults
Disutility for moderate exacerbation
Disutility for ER-level exacerbation
Disutility for hospital-level exacerbation
Direct cost per month for well-controlled asthma
Direct cost per month for not well-controlled asthma
Direct cost of moderate exacerbation
Direct cost of ER-level exacerbation
Probability of death from severe exacerbation (ER visit)
Direct cost of hospitalization
Probability of death from severe exacerbation (SCS)
Probability of hospitalization from well-controlled asthma
Initial age for the adult cohort
PM2.5 exposure scenario (e.g., projection levels)
Proportion of outdoor PM2.5 that infiltrates indoors
Proportion of time spent at home, stratified by age
Effectiveness of HEPA air cleaner in reducing PM2.5 indoors
Concentration-response function (CRF) for asthma incidence
Relative risk for asthma control
Relative risk for moderate exacerbation (oral SCS)
Relative risk for severe exacerbation (ER visit)
Relative risk for severe exacerbation (hospitalization)
Relative risk of exacerbation for not well-controlled asthma
Probability of transitioning from well-controlled asthma
Probability of transition from well-controlled to not well-controlled asthma
Probability of moderate exacerbation from well-controlled asthma
Probability of ER visit from well-controlled asthma
Probability of improving from not well-controlled to well-controlled asthma
Cost of HEPA air cleaner (adjusted for rebate/discount)
100000
150000
200000
250000
ICER (CAD per QALY)

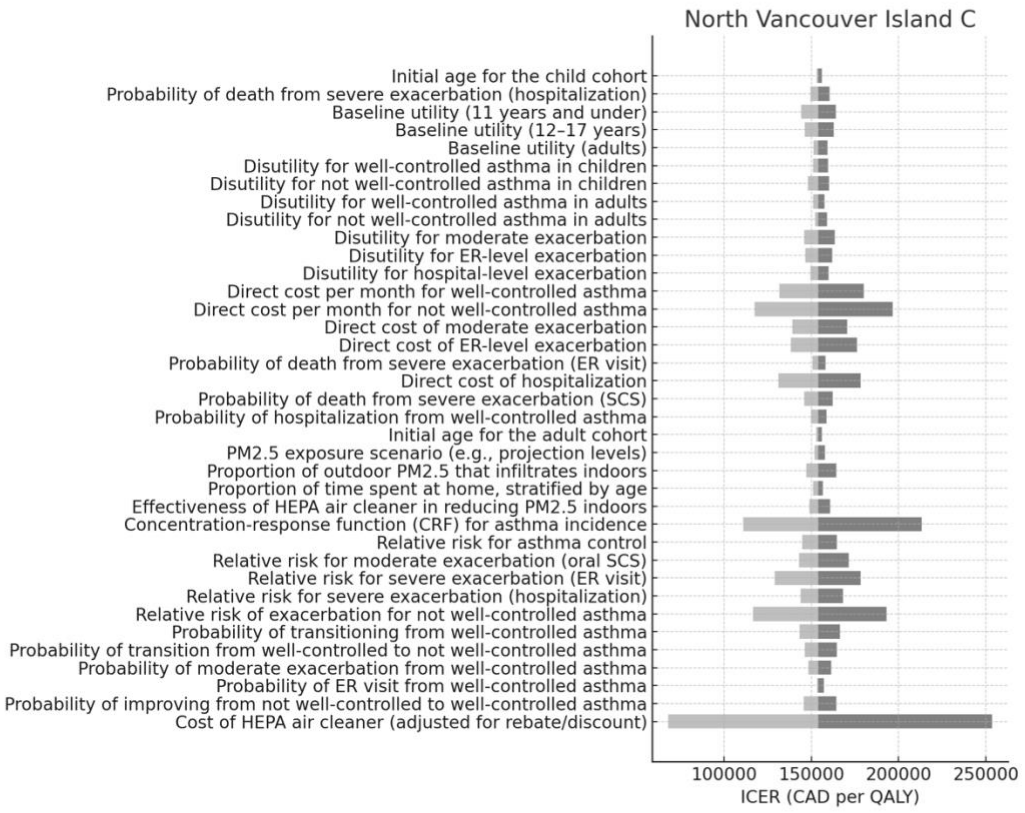
North Vancouver Island C
Initial age for the child cohort
Probability of death from severe exacerbation (hospitalization)
Baseline utility (11 years and under)
Baseline utility (12–17 years)
Baseline utility (adults)
Disutility for well-controlled asthma in children
Disutility for not well-controlled asthma in children
Disutility for well-controlled asthma in adults
Disutility for not well-controlled asthma in adults
Disutility for moderate exacerbation
Disutility for ER-level exacerbation
Disutility for hospital-level exacerbation
Direct cost per month for well-controlled asthma
Direct cost per month for not well-controlled asthma
Direct cost of moderate exacerbation
Direct cost of ER-level exacerbation
Probability of death from severe exacerbation (ER visit)
Direct cost of hospitalization
Probability of death from severe exacerbation (SCS)
Probability of hospitalization from well-controlled asthma
Initial age for the adult cohort
PM2.5 exposure scenario (e.g., projection levels)
Proportion of outdoor PM2.5 that infiltrates indoors
Proportion of time spent at home, stratified by age
Effectiveness of HEPA air cleaner in reducing PM2.5 indoors
Concentration-response function (CRF) for asthma incidence
Relative risk for asthma control
Relative risk for moderate exacerbation (oral SCS)
Relative risk for severe exacerbation (ER visit)
Relative risk for severe exacerbation (hospitalization)
Relative risk of exacerbation for not well-controlled asthma
Probability of transitioning from well-controlled asthma
Probability of transition from well-controlled to not well-controlled asthma
Probability of moderate exacerbation from well-controlled asthma
Probability of ER visit from well-controlled asthma
Probability of improving from not well-controlled to well-controlled asthma
Cost of HEPA air cleaner (adjusted for rebate/discount)
100000
150000
200000
250000
ICER (CAD per QALY)

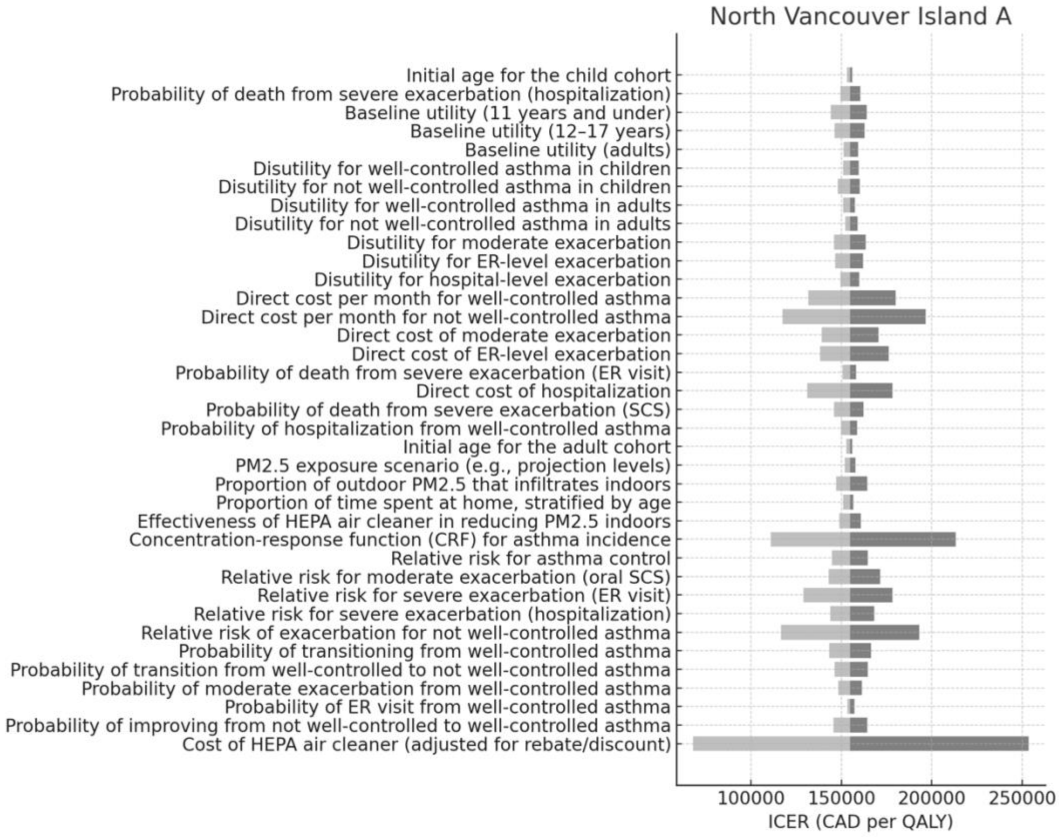
North Vancouver Island A
Initial age for the child cohort
Probability of death from severe exacerbation (hospitalization)
Baseline utility (11 years and under)
Baseline utility (12–17 years)
Baseline utility (adults)
Disutility for well-controlled asthma in children
Disutility for not well-controlled asthma in children
Disutility for well-controlled asthma in adults
Disutility for not well-controlled asthma in adults
Disutility for moderate exacerbation
Disutility for ER-level exacerbation
Disutility for hospital-level exacerbation
Direct cost per month for well-controlled asthma
Direct cost per month for not well-controlled asthma
Direct cost of moderate exacerbation
Direct cost of ER-level exacerbation
Probability of death from severe exacerbation (ER visit)
Direct cost of hospitalization
Probability of death from severe exacerbation (SCS)
Probability of hospitalization from well-controlled asthma
Initial age for the adult cohort
PM2.5 exposure scenario (e.g., projection levels)
Proportion of outdoor PM2.5 that infiltrates indoors
Proportion of time spent at home, stratified by age
Effectiveness of HEPA air cleaner in reducing PM2.5 indoors
Concentration-response function (CRF) for asthma incidence
Relative risk for asthma control
Relative risk for moderate exacerbation (oral SCS)
Relative risk for severe exacerbation (ER visit)
Relative risk for severe exacerbation (hospitalization)
Relative risk of exacerbation for not well-controlled asthma
Probability of transitioning from well-controlled asthma
Probability of transition from well-controlled to not well-controlled asthma
Probability of moderate exacerbation from well-controlled asthma
Probability of ER visit from well-controlled asthma
Probability of improving from not well-controlled to well-controlled asthma
Cost of HEPA air cleaner (adjusted for rebate/discount)
100000
150000
200000
250000
ICER (CAD per QALY)

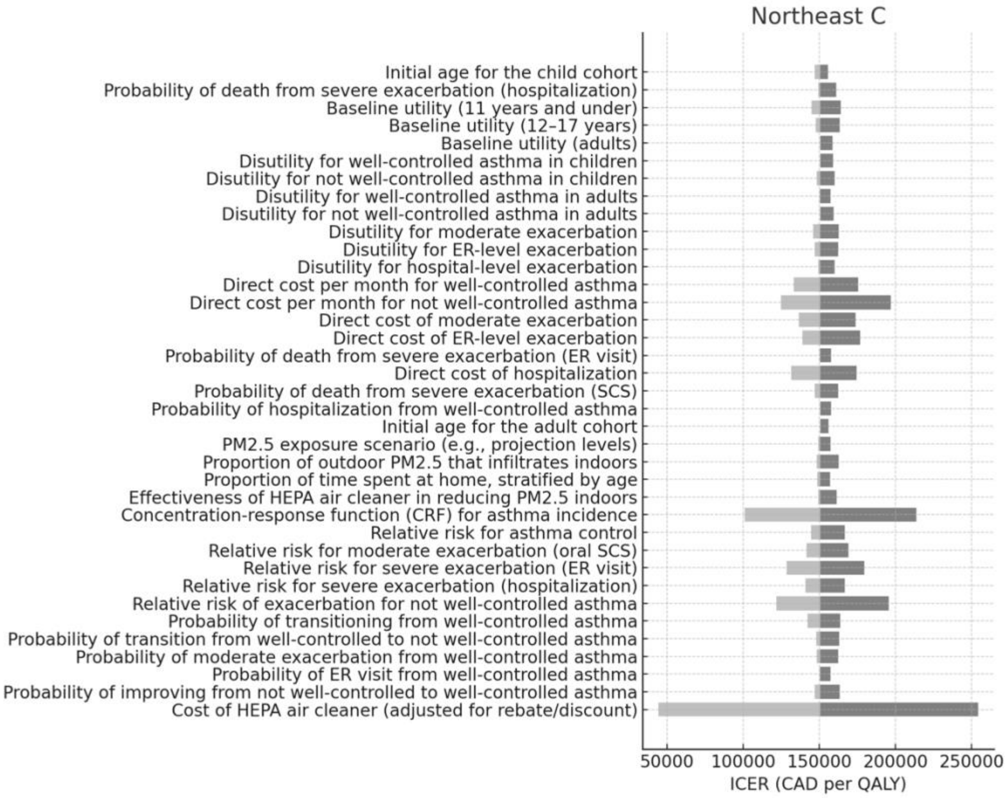
Northeast C
Initial age for the child cohort
Probability of death from severe exacerbation (hospitalization)
Baseline utility (11 years and under)
Baseline utility (12–17 years)
Baseline utility (adults)
Disutility for well-controlled asthma in children
Disutility for not well-controlled asthma in children
Disutility for well-controlled asthma in adults
Disutility for not well-controlled asthma in adults
Disutility for moderate exacerbation
Disutility for ER-level exacerbation
Disutility for hospital-level exacerbation
Direct cost per month for well-controlled asthma
Direct cost per month for not well-controlled asthma
Direct cost of moderate exacerbation
Direct cost of ER-level exacerbation
Probability of death from severe exacerbation (ER visit)
Direct cost of hospitalization
Probability of death from severe exacerbation (SCS)
Probability of hospitalization from well-controlled asthma
Initial age for the adult cohort
PM2.5 exposure scenario (e.g., projection levels)
Proportion of outdoor PM2.5 that infiltrates indoors
Proportion of time spent at home, stratified by age
Effectiveness of HEPA air cleaner in reducing PM2.5 indoors
Concentration-response function (CRF) for asthma incidence
Relative risk for asthma control
Relative risk for moderate exacerbation (oral SCS)
Relative risk for severe exacerbation (ER visit)
Relative risk for severe exacerbation (hospitalization)
Relative risk of exacerbation for not well-controlled asthma
Probability of transitioning from well-controlled asthma
Probability of transition from well-controlled to not well-controlled asthma
Probability of moderate exacerbation from well-controlled asthma
Probability of ER visit from well-controlled asthma
Probability of improving from not well-controlled to well-controlled asthma
Cost of HEPA air cleaner (adjusted for rebate/discount)
50000
100000
150000
200000
250000
ICER (CAD per QALY)

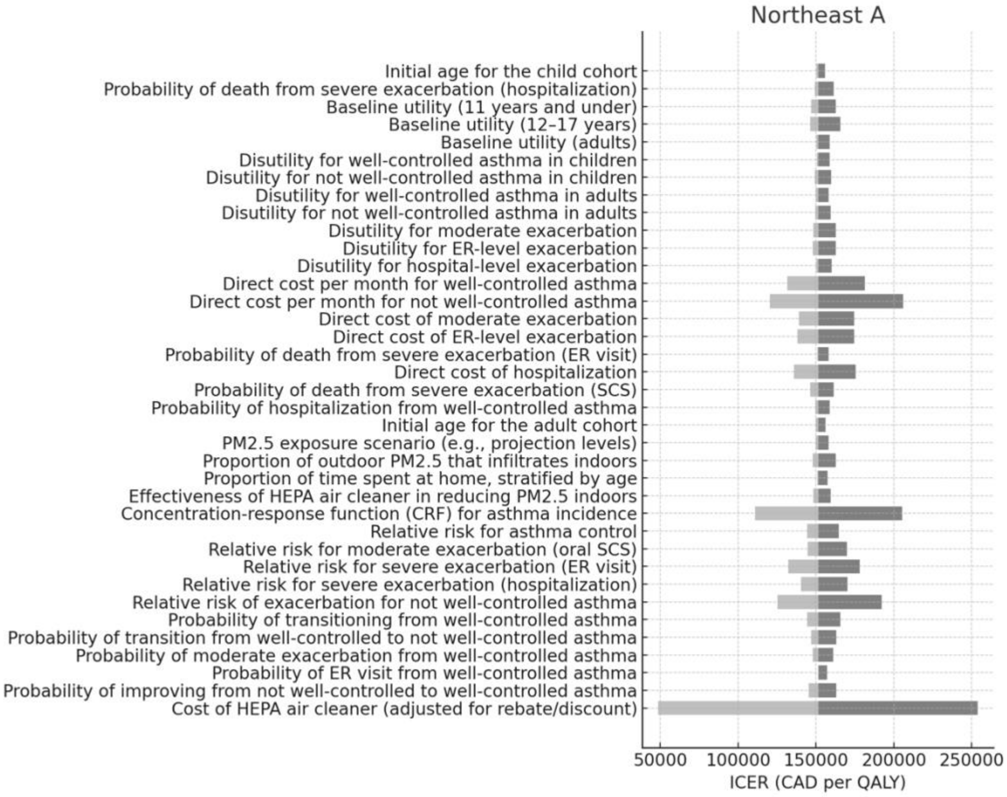
Northeast A
Initial age for the child cohort
Probability of death from severe exacerbation (hospitalization)
Baseline utility (11 years and under)
Baseline utility (12–17 years)
Baseline utility (adults)
Disutility for well-controlled asthma in children
Disutility for not well-controlled asthma in children
Disutility for well-controlled asthma in adults
Disutility for not well-controlled asthma in adults
Disutility for moderate exacerbation
Disutility for ER-level exacerbation
Disutility for hospital-level exacerbation
Direct cost per month for well-controlled asthma
Direct cost per month for not well-controlled asthma
Direct cost of moderate exacerbation
Direct cost of ER-level exacerbation
Probability of death from severe exacerbation (ER visit)
Direct cost of hospitalization
Probability of death from severe exacerbation (SCS)
Probability of hospitalization from well-controlled asthma
Initial age for the adult cohort
PM2.5 exposure scenario (e.g., projection levels)
Proportion of outdoor PM2.5 that infiltrates indoors
Proportion of time spent at home, stratified by age
Effectiveness of HEPA air cleaner in reducing PM2.5 indoors
Concentration-response function (CRF) for asthma incidence
Relative risk for asthma control
Relative risk for moderate exacerbation (oral SCS)
Relative risk for severe exacerbation (ER visit)
Relative risk for severe exacerbation (hospitalization)
Relative risk of exacerbation for not well-controlled asthma
Probability of transitioning from well-controlled asthma
Probability of transition from well-controlled to not well-controlled asthma
Probability of moderate exacerbation from well-controlled asthma
Probability of ER visit from well-controlled asthma
Probability of improving from not well-controlled to well-controlled asthma
Cost of HEPA air cleaner (adjusted for rebate/discount)
50000
100000
150000
200000
250000
ICER (CAD per QALY)

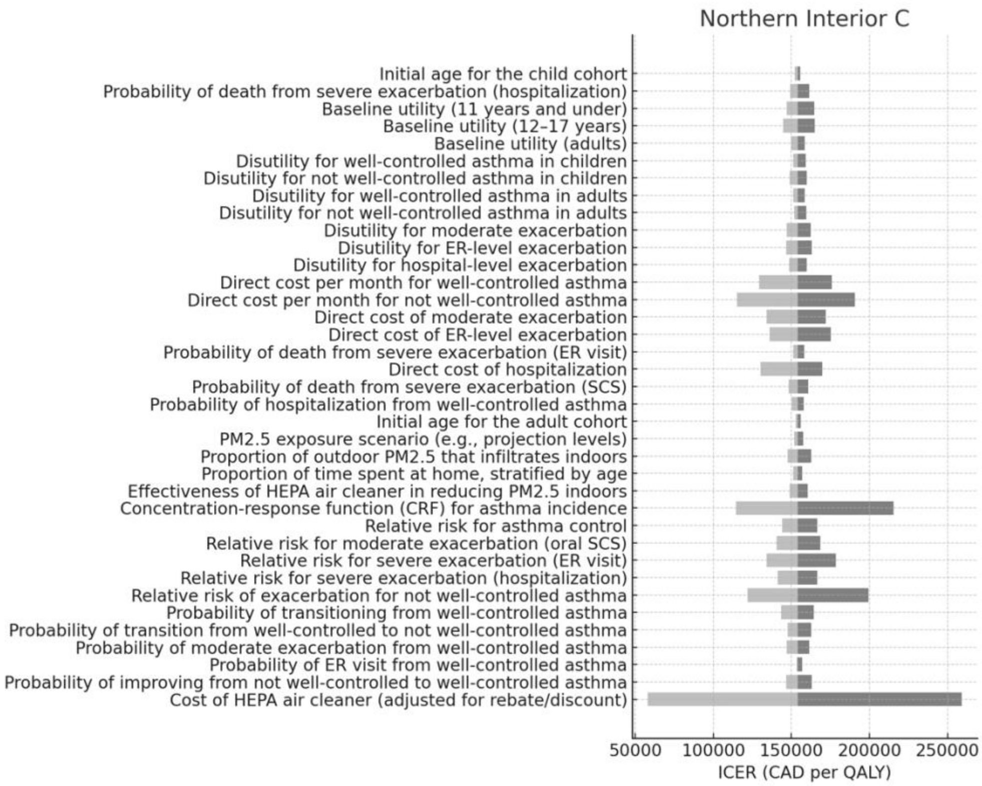
Northern Interior C
Initial age for the child cohort
Probability of death from severe exacerbation (hospitalization)
Baseline utility (11 years and under)
Baseline utility (12–17 years)
Baseline utility (adults)
Disutility for well-controlled asthma in children
Disutility for not well-controlled asthma in children
Disutility for well-controlled asthma in adults
Disutility for not well-controlled asthma in adults
Disutility for moderate exacerbation
Disutility for ER-level exacerbation
Disutility for hospital-level exacerbation
Direct cost per month for well-controlled asthma
Direct cost per month for not well-controlled asthma
Direct cost of moderate exacerbation
Direct cost of ER-level exacerbation
Probability of death from severe exacerbation (ER visit)
Direct cost of hospitalization
Probability of death from severe exacerbation (SCS)
Probability of hospitalization from well-controlled asthma
Initial age for the adult cohort
PM2.5 exposure scenario (e.g., projection levels)
Proportion of outdoor PM2.5 that infiltrates indoors
Proportion of time spent at home, stratified by age
Effectiveness of HEPA air cleaner in reducing PM2.5 indoors
Concentration-response function (CRF) for asthma incidence
Relative risk for asthma control
Relative risk for moderate exacerbation (oral SCS)
Relative risk for severe exacerbation (ER visit)
Relative risk for severe exacerbation (hospitalization)
Relative risk of exacerbation for not well-controlled asthma
Probability of transitioning from well-controlled asthma
Probability of transition from well-controlled to not well-controlled asthma
Probability of moderate exacerbation from well-controlled asthma
Probability of ER visit from well-controlled asthma
Probability of improving from not well-controlled to well-controlled asthma
Cost of HEPA air cleaner (adjusted for rebate/discount)
50000
100000
150000
200000
250000
ICER (CAD per QALY)

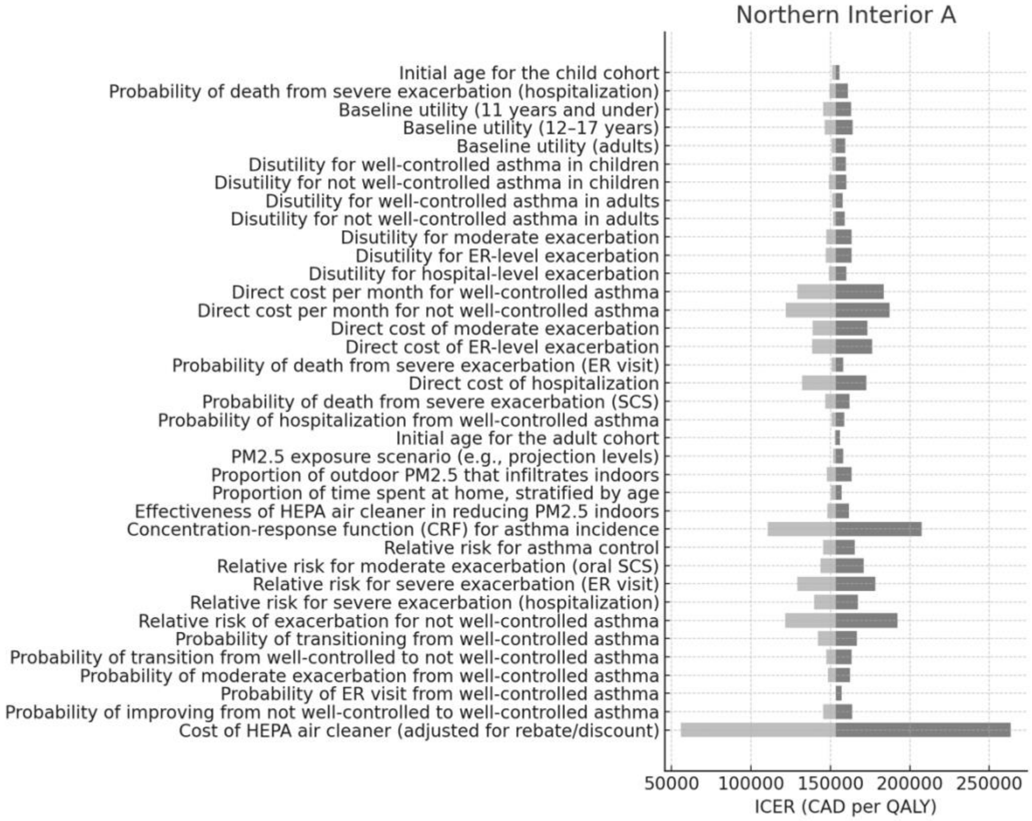
Northern Interior A
Initial age for the child cohort
Probability of death from severe exacerbation (hospitalization)
Baseline utility (11 years and under)
Baseline utility (12–17 years)
Baseline utility (adults)
Disutility for well-controlled asthma in children
Disutility for not well-controlled asthma in children
Disutility for well-controlled asthma in adults
Disutility for not well-controlled asthma in adults
Disutility for moderate exacerbation
Disutility for ER-level exacerbation
Disutility for hospital-level exacerbation
Direct cost per month for well-controlled asthma
Direct cost per month for not well-controlled asthma
Direct cost of moderate exacerbation
Direct cost of ER-level exacerbation
Probability of death from severe exacerbation (ER visit)
Direct cost of hospitalization
Probability of death from severe exacerbation (SCS)
Probability of hospitalization from well-controlled asthma
Initial age for the adult cohort
PM2.5 exposure scenario (e.g., projection levels)
Proportion of outdoor PM2.5 that infiltrates indoors
Proportion of time spent at home, stratified by age
Effectiveness of HEPA air cleaner in reducing PM2.5 indoors
Concentration-response function (CRF) for asthma incidence
Relative risk for asthma control
Relative risk for moderate exacerbation (oral SCS)
Relative risk for severe exacerbation (ER visit)
Relative risk for severe exacerbation (hospitalization)
Relative risk of exacerbation for not well-controlled asthma
Probability of transitioning from well-controlled asthma
Probability of transition from well-controlled to not well-controlled asthma
Probability of moderate exacerbation from well-controlled asthma
Probability of ER visit from well-controlled asthma
Probability of improving from not well-controlled to well-controlled asthma
Cost of HEPA air cleaner (adjusted for rebate/discount)
50000
100000
150000
200000
250000
ICER (CAD per QALY)

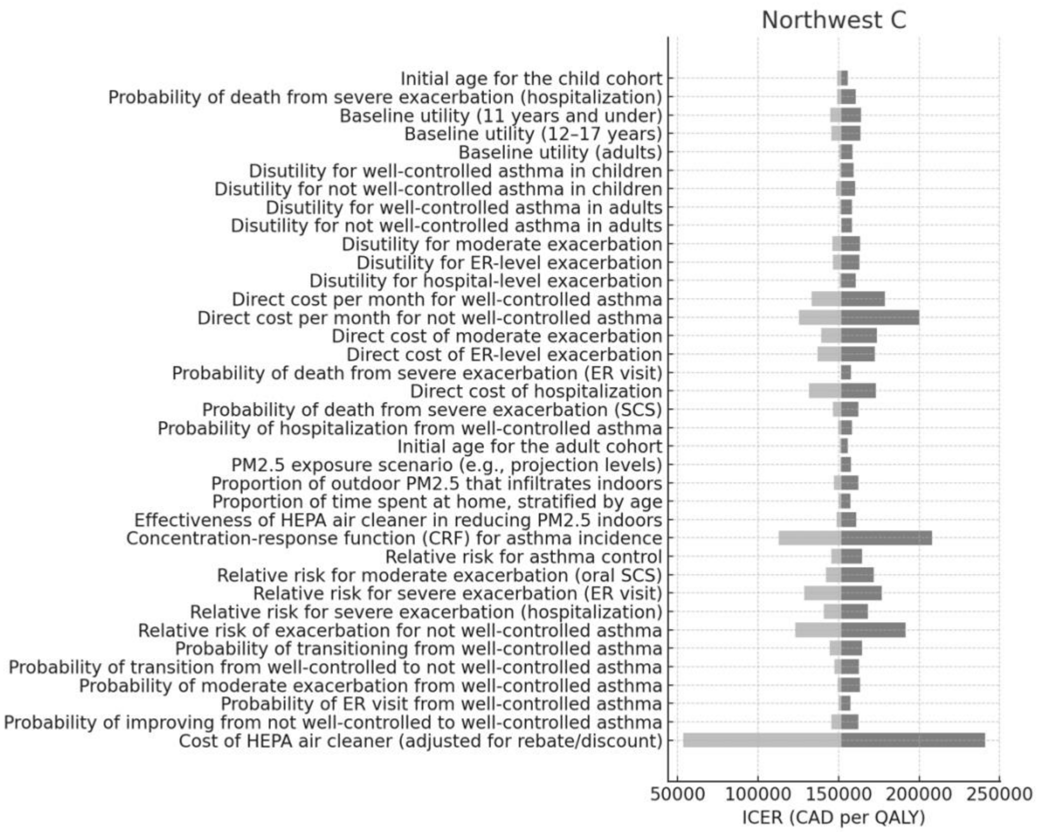
Northwest C
Initial age for the child cohort
Probability of death from severe exacerbation (hospitalization)
Baseline utility (11 years and under)
Baseline utility (12–17 years)
Baseline utility (adults)
Disutility for well-controlled asthma in children
Disutility for not well-controlled asthma in children
Disutility for well-controlled asthma in adults
Disutility for not well-controlled asthma in adults
Disutility for moderate exacerbation
Disutility for ER-level exacerbation
Disutility for hospital-level exacerbation
Direct cost per month for well-controlled asthma
Direct cost per month for not well-controlled asthma
Direct cost of moderate exacerbation
Direct cost of ER-level exacerbation
Probability of death from severe exacerbation (ER visit)
Direct cost of hospitalization
Probability of death from severe exacerbation (SCS)
Probability of hospitalization from well-controlled asthma
Initial age for the adult cohort
PM2.5 exposure scenario (e.g., projection levels)
Proportion of outdoor PM2.5 that infiltrates indoors
Proportion of time spent at home, stratified by age
Effectiveness of HEPA air cleaner in reducing PM2.5 indoors
Concentration-response function (CRF) for asthma incidence
Relative risk for asthma control
Relative risk for moderate exacerbation (oral SCS)
Relative risk for severe exacerbation (ER visit)
Relative risk for severe exacerbation (hospitalization)
Relative risk of exacerbation for not well-controlled asthma
Probability of transitioning from well-controlled asthma
Probability of transition from well-controlled to not well-controlled asthma
Probability of moderate exacerbation from well-controlled asthma
Probability of ER visit from well-controlled asthma
Probability of improving from not well-controlled to well-controlled asthma
Cost of HEPA air cleaner (adjusted for rebate/discount)
50000
100000
150000
200000
250000
ICER (CAD per QALY)

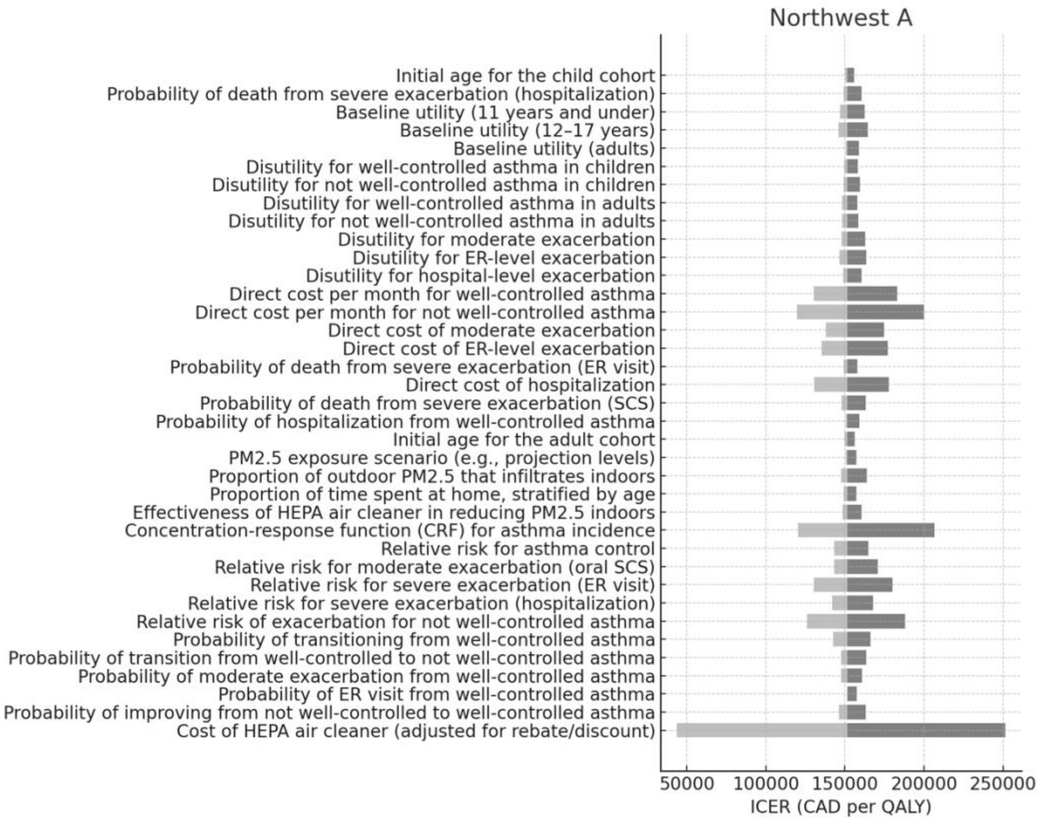
Northwest A
Initial age for the child cohort
Probability of death from severe exacerbation (hospitalization)
Baseline utility (11 years and under)
Baseline utility (12–17 years)
Baseline utility (adults)
Disutility for well-controlled asthma in children
Disutility for not well-controlled asthma in children
Disutility for well-controlled asthma in adults
Disutility for not well-controlled asthma in adults
Disutility for moderate exacerbation
Disutility for ER-level exacerbation
Disutility for hospital-level exacerbation
Direct cost per month for well-controlled asthma
Direct cost per month for not well-controlled asthma
Direct cost of moderate exacerbation
Direct cost of ER-level exacerbation
Probability of death from severe exacerbation (ER visit)
Direct cost of hospitalization
Probability of death from severe exacerbation (SCS)
Probability of hospitalization from well-controlled asthma
Initial age for the adult cohort
PM2.5 exposure scenario (e.g., projection levels)
Proportion of outdoor PM2.5 that infiltrates indoors
Proportion of time spent at home, stratified by age
Effectiveness of HEPA air cleaner in reducing PM2.5 indoors
Concentration-response function (CRF) for asthma incidence
Relative risk for asthma control
Relative risk for moderate exacerbation (oral SCS)
Relative risk for severe exacerbation (ER visit)
Relative risk for severe exacerbation (hospitalization)
Relative risk of exacerbation for not well-controlled asthma
Probability of transitioning from well-controlled asthma
Probability of transition from well-controlled to not well-controlled asthma
Probability of moderate exacerbation from well-controlled asthma
Probability of ER visit from well-controlled asthma
Probability of improving from not well-controlled to well-controlled asthma
Cost of HEPA air cleaner (adjusted for rebate/discount)
50000
100000
150000
200000
250000
ICER (CAD per QALY)

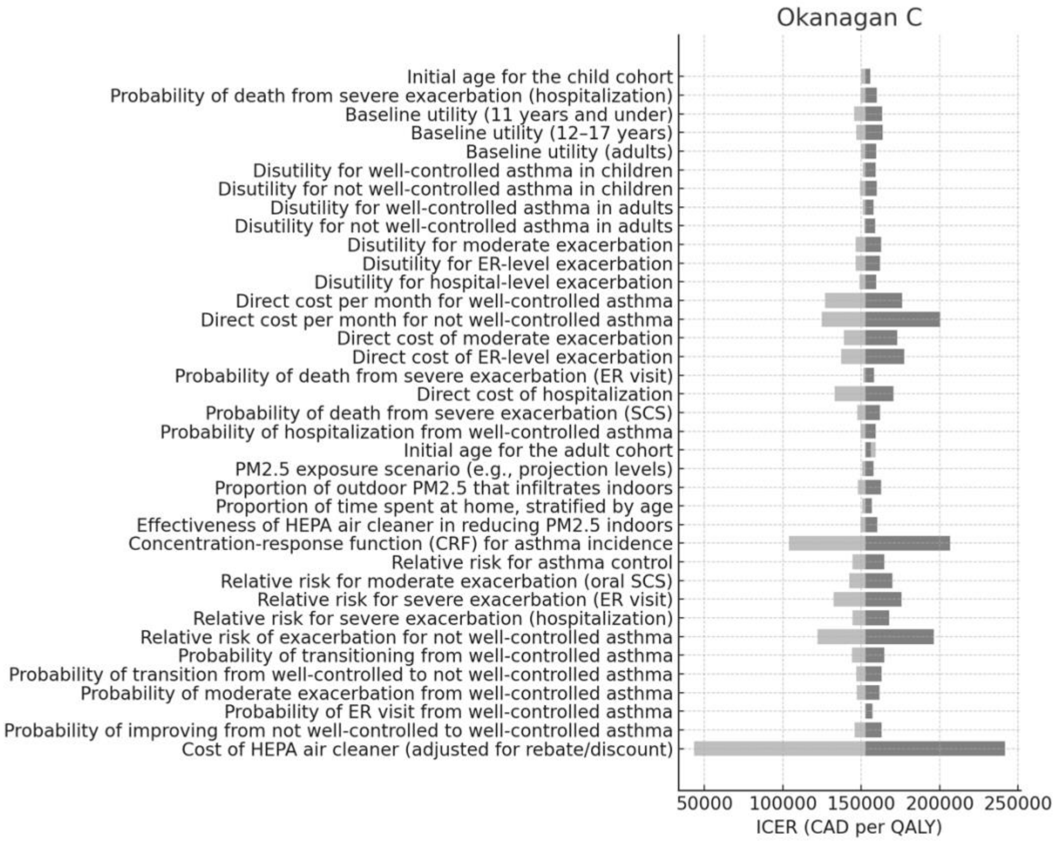

Okanagan C
Initial age for the child cohort
Probability of death from severe exacerbation (hospitalization)
Baseline utility (11 years and under)
Baseline utility (12–17 years)
Baseline utility (adults)
Disutility for well-controlled asthma in children
Disutility for not well-controlled asthma in children
Disutility for well-controlled asthma in adults
Disutility for not well-controlled asthma in adults
Disutility for moderate exacerbation
Disutility for ER-level exacerbation
Disutility for hospital-level exacerbation
Direct cost per month for well-controlled asthma
Direct cost per month for not well-controlled asthma
Direct cost of moderate exacerbation
Direct cost of ER-level exacerbation
Probability of death from severe exacerbation (ER visit)
Direct cost of hospitalization
Probability of death from severe exacerbation (SCS)
Probability of hospitalization from well-controlled asthma
Initial age for the adult cohort
PM2.5 exposure scenario (e.g., projection levels)
Proportion of outdoor PM2.5 that infiltrates indoors
Proportion of time spent at home, stratified by age
Effectiveness of HEPA air cleaner in reducing PM2.5 indoors
Concentration-response function (CRF) for asthma incidence
Relative risk for asthma control
Relative risk for moderate exacerbation (oral SCS)
Relative risk for severe exacerbation (ER visit)
Relative risk for severe exacerbation (hospitalization)
Relative risk of exacerbation for not well-controlled asthma
Probability of transitioning from well-controlled asthma
Probability of transition from well-controlled to not well-controlled asthma
Probability of moderate exacerbation from well-controlled asthma
Probability of ER visit from well-controlled asthma
Probability of improving from not well-controlled to well-controlled asthma
Cost of HEPA air cleaner (adjusted for rebate/discount)
50000 100000 150000 200000 250000
ICER (CAD per QALY)


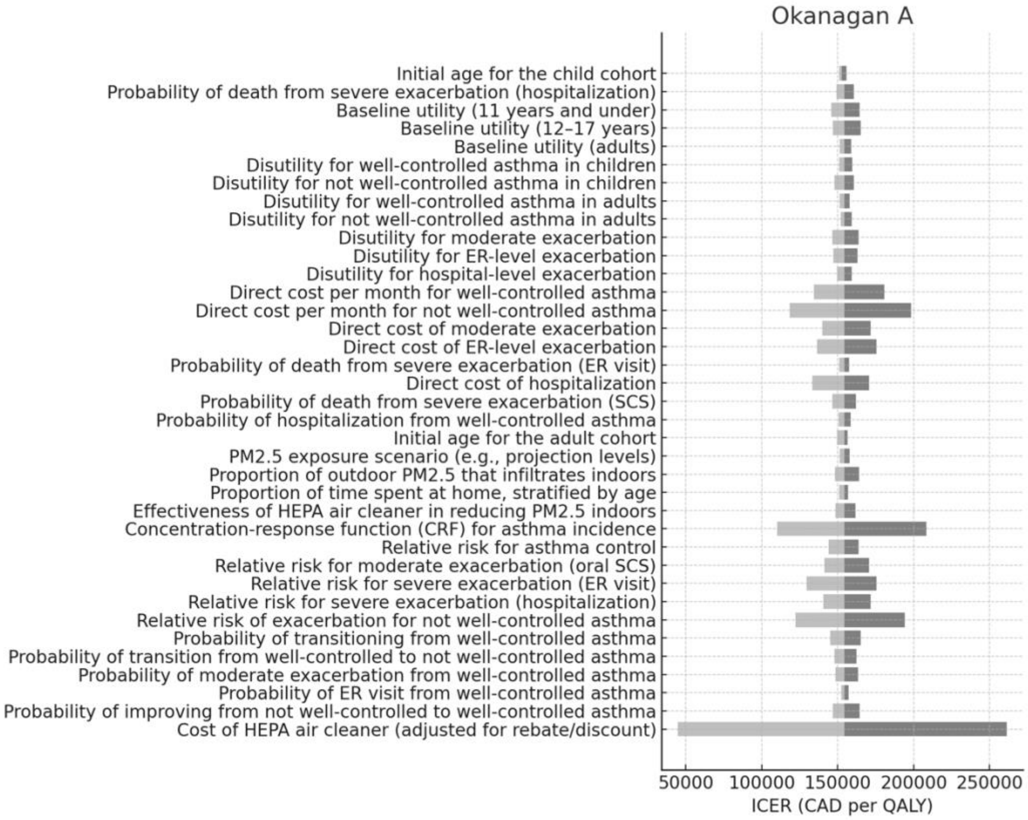

Okanagan A
Initial age for the child cohort
Probability of death from severe exacerbation (hospitalization)
Baseline utility (11 years and under)
Baseline utility (12–17 years)
Baseline utility (adults)
Disutility for well-controlled asthma in children
Disutility for not well-controlled asthma in children
Disutility for well-controlled asthma in adults
Disutility for not well-controlled asthma in adults
Disutility for moderate exacerbation
Disutility for ER-level exacerbation
Disutility for hospital-level exacerbation
Direct cost per month for well-controlled asthma
Direct cost per month for not well-controlled asthma
Direct cost of moderate exacerbation
Direct cost of ER-level exacerbation
Probability of death from severe exacerbation (ER visit)
Direct cost of hospitalization
Probability of death from severe exacerbation (SCS)
Probability of hospitalization from well-controlled asthma
Initial age for the adult cohort
PM2.5 exposure scenario (e.g., projection levels)
Proportion of outdoor PM2.5 that infiltrates indoors
Proportion of time spent at home, stratified by age
Effectiveness of HEPA air cleaner in reducing PM2.5 indoors
Concentration-response function (CRF) for asthma incidence
Relative risk for asthma control
Relative risk for moderate exacerbation (oral SCS)
Relative risk for severe exacerbation (ER visit)
Relative risk for severe exacerbation (hospitalization)
Relative risk of exacerbation for not well-controlled asthma
Probability of transitioning from well-controlled asthma
Probability of transition from well-controlled to not well-controlled asthma
Probability of moderate exacerbation from well-controlled asthma
Probability of ER visit from well-controlled asthma
Probability of improving from not well-controlled to well-controlled asthma
Cost of HEPA air cleaner (adjusted for rebate/discount)
50000 100000 150000 200000 250000
ICER (CAD per QALY)

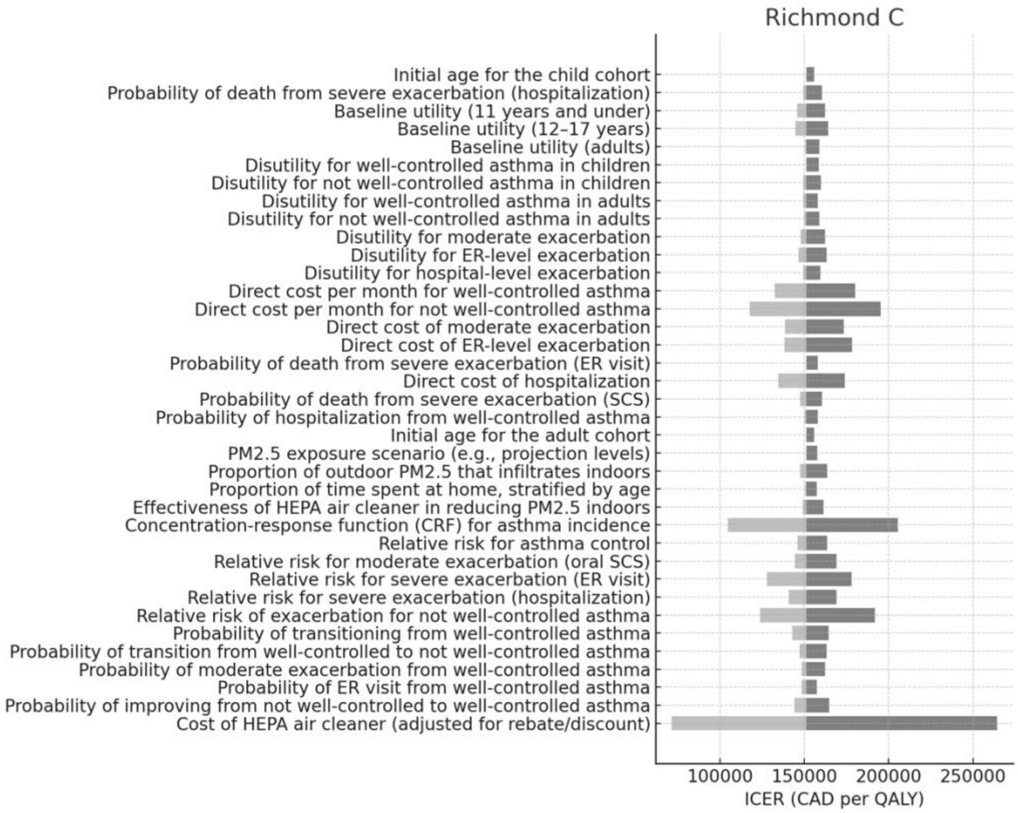
Richmond C
Initial age for the child cohort
Probability of death from severe exacerbation (hospitalization)
Baseline utility (11 years and under)
Baseline utility (12–17 years)
Baseline utility (adults)
Disutility for well-controlled asthma in children
Disutility for not well-controlled asthma in children
Disutility for well-controlled asthma in adults
Disutility for not well-controlled asthma in adults
Disutility for moderate exacerbation
Disutility for ER-level exacerbation
Disutility for hospital-level exacerbation
Direct cost per month for well-controlled asthma
Direct cost per month for not well-controlled asthma
Direct cost of moderate exacerbation
Direct cost of ER-level exacerbation
Probability of death from severe exacerbation (ER visit)
Direct cost of hospitalization
Probability of death from severe exacerbation (SCS)
Probability of hospitalization from well-controlled asthma
Initial age for the adult cohort
PM2.5 exposure scenario (e.g., projection levels)
Proportion of outdoor PM2.5 that infiltrates indoors
Proportion of time spent at home, stratified by age
Effectiveness of HEPA air cleaner in reducing PM2.5 indoors
Concentration-response function (CRF) for asthma incidence
Relative risk for asthma control
Relative risk for moderate exacerbation (oral SCS)
Relative risk for severe exacerbation (ER visit)
Relative risk for severe exacerbation (hospitalization)
Relative risk of exacerbation for not well-controlled asthma
Probability of transitioning from well-controlled asthma
Probability of transition from well-controlled to not well-controlled asthma
Probability of moderate exacerbation from well-controlled asthma
Probability of ER visit from well-controlled asthma
Probability of improving from not well-controlled to well-controlled asthma
Cost of HEPA air cleaner (adjusted for rebate/discount)
100000
150000
200000
250000
ICER (CAD per QALY)

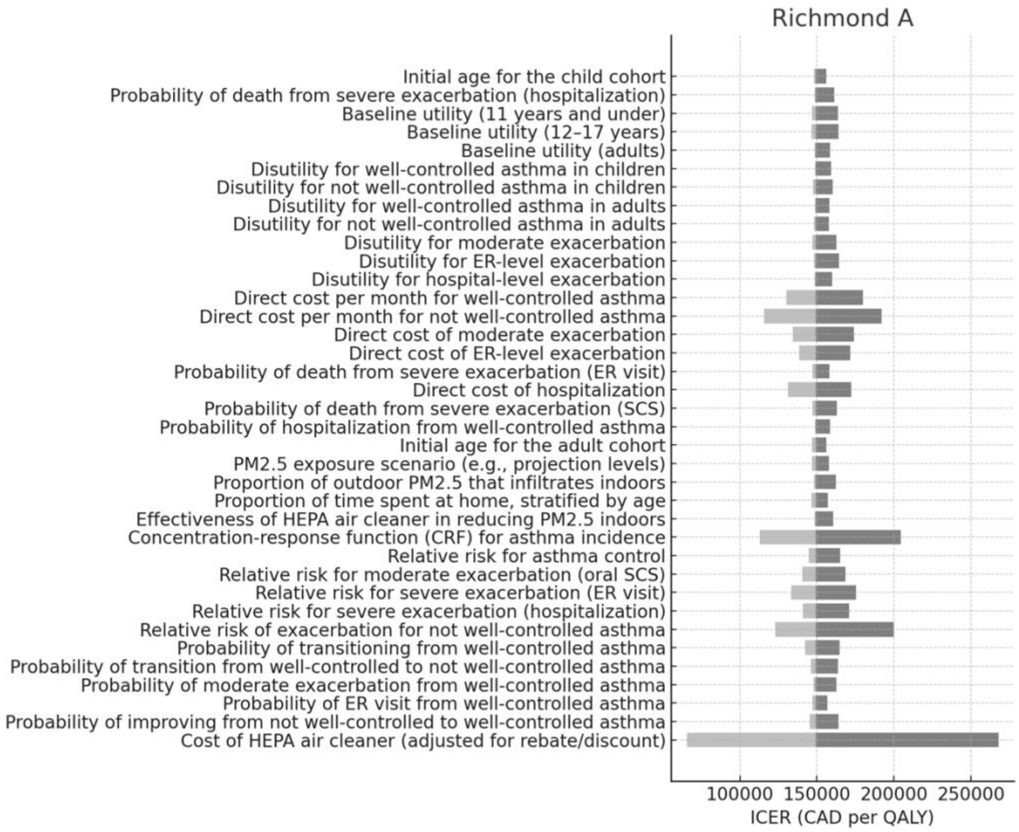
Richmond A
Initial age for the child cohort
Probability of death from severe exacerbation (hospitalization)
Baseline utility (11 years and under)
Baseline utility (12–17 years)
Baseline utility (adults)
Disutility for well-controlled asthma in children
Disutility for not well-controlled asthma in children
Disutility for well-controlled asthma in adults
Disutility for not well-controlled asthma in adults
Disutility for moderate exacerbation
Disutility for ER-level exacerbation
Disutility for hospital-level exacerbation
Direct cost per month for well-controlled asthma
Direct cost per month for not well-controlled asthma
Direct cost of moderate exacerbation
Direct cost of ER-level exacerbation
Probability of death from severe exacerbation (ER visit)
Direct cost of hospitalization
Probability of death from severe exacerbation (SCS)
Probability of hospitalization from well-controlled asthma
Initial age for the adult cohort
PM2.5 exposure scenario (e.g., projection levels)
Proportion of outdoor PM2.5 that infiltrates indoors
Proportion of time spent at home, stratified by age
Effectiveness of HEPA air cleaner in reducing PM2.5 indoors
Concentration-response function (CRF) for asthma incidence
Relative risk for asthma control
Relative risk for moderate exacerbation (oral SCS)
Relative risk for severe exacerbation (ER visit)
Relative risk for severe exacerbation (hospitalization)
Relative risk of exacerbation for not well-controlled asthma
Probability of transitioning from well-controlled asthma
Probability of transition from well-controlled to not well-controlled asthma
Probability of moderate exacerbation from well-controlled asthma
Probability of ER visit from well-controlled asthma
Probability of improving from not well-controlled to well-controlled asthma
Cost of HEPA air cleaner (adjusted for rebate/discount)
100000
150000
200000
250000
ICER (CAD per QALY)

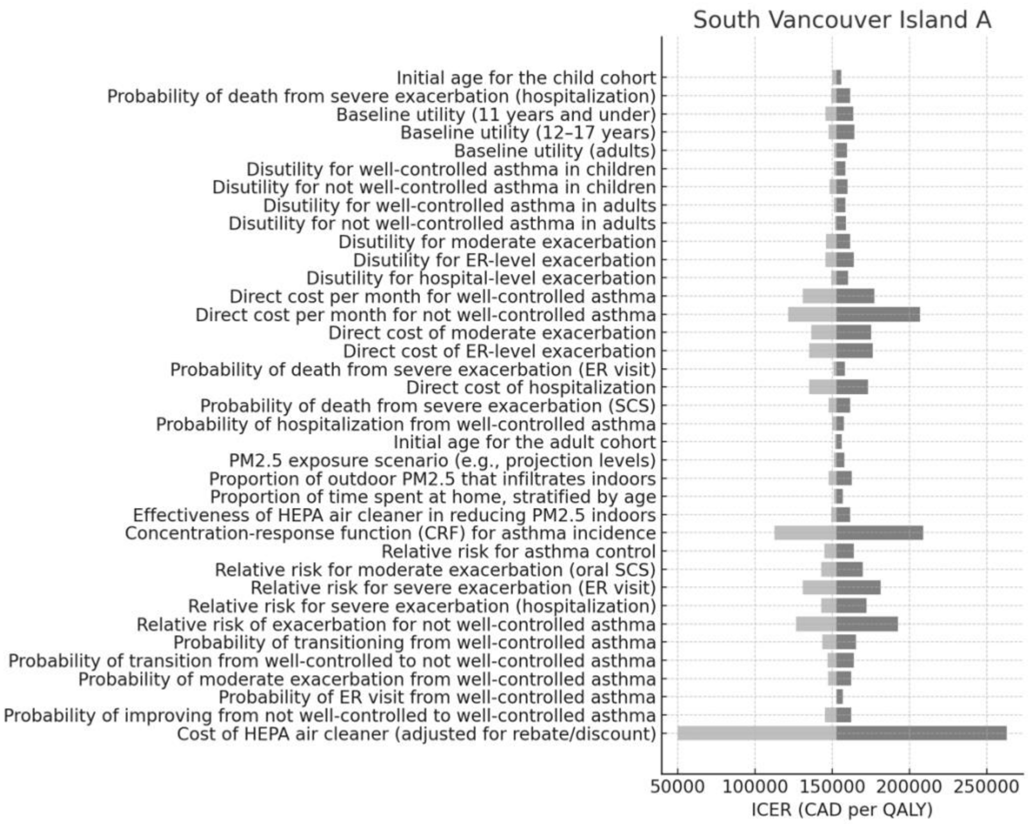
South Vancouver Island A
Initial age for the child cohort
Probability of death from severe exacerbation (hospitalization)
Baseline utility (11 years and under)
Baseline utility (12–17 years)
Baseline utility (adults)
Disutility for well-controlled asthma in children
Disutility for not well-controlled asthma in children
Disutility for well-controlled asthma in adults
Disutility for not well-controlled asthma in adults
Disutility for moderate exacerbation
Disutility for ER-level exacerbation
Disutility for hospital-level exacerbation
Direct cost per month for well-controlled asthma
Direct cost per month for not well-controlled asthma
Direct cost of moderate exacerbation
Direct cost of ER-level exacerbation
Probability of death from severe exacerbation (ER visit)
Direct cost of hospitalization
Probability of death from severe exacerbation (SCS)
Probability of hospitalization from well-controlled asthma
Initial age for the adult cohort
PM2.5 exposure scenario (e.g., projection levels)
Proportion of outdoor PM2.5 that infiltrates indoors
Proportion of time spent at home, stratified by age
Effectiveness of HEPA air cleaner in reducing PM2.5 indoors
Concentration-response function (CRF) for asthma incidence
Relative risk for asthma control
Relative risk for moderate exacerbation (oral SCS)
Relative risk for severe exacerbation (ER visit)
Relative risk for severe exacerbation (hospitalization)
Relative risk of exacerbation for not well-controlled asthma
Probability of transitioning from well-controlled asthma
Probability of transition from well-controlled to not well-controlled asthma
Probability of moderate exacerbation from well-controlled asthma
Probability of ER visit from well-controlled asthma
Probability of improving from not well-controlled to well-controlled asthma
Cost of HEPA air cleaner (adjusted for rebate/discount)
50000
100000
150000
200000
250000
ICER (CAD per QALY)

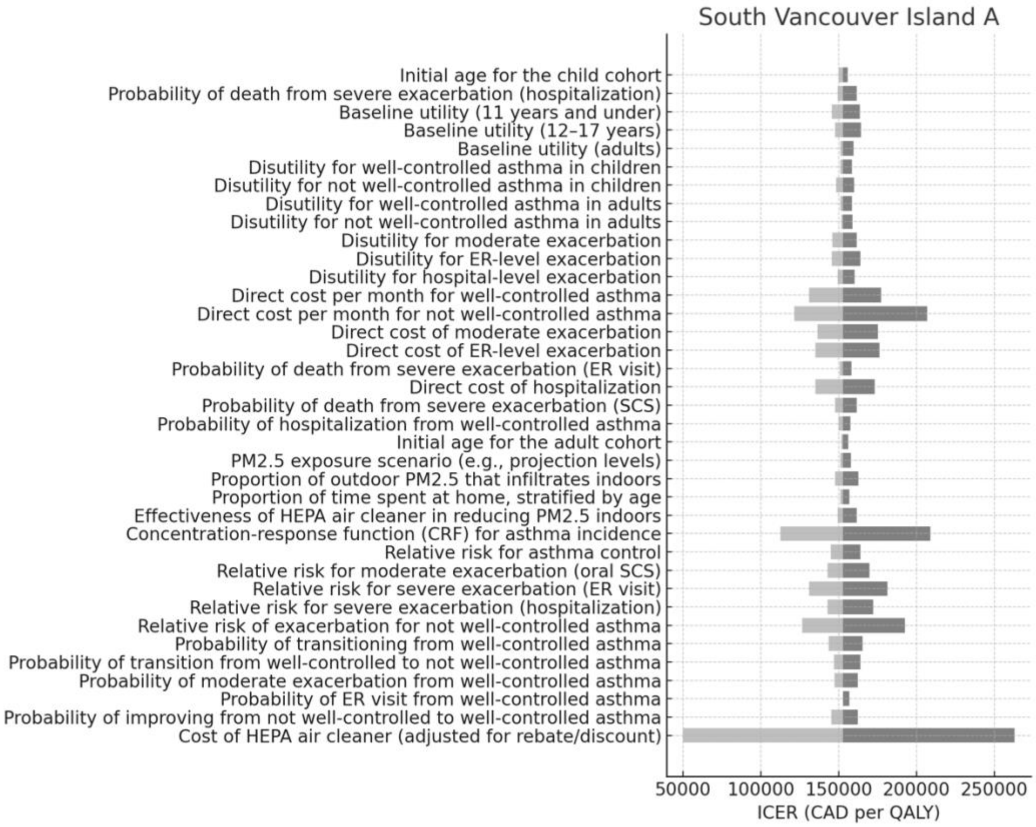
South Vancouver Island A
Initial age for the child cohort
Probability of death from severe exacerbation (hospitalization)
Baseline utility (11 years and under)
Baseline utility (12–17 years)
Baseline utility (adults)
Disutility for well-controlled asthma in children
Disutility for not well-controlled asthma in children
Disutility for well-controlled asthma in adults
Disutility for not well-controlled asthma in adults
Disutility for moderate exacerbation
Disutility for ER-level exacerbation
Disutility for hospital-level exacerbation
Direct cost per month for well-controlled asthma
Direct cost per month for not well-controlled asthma
Direct cost of moderate exacerbation
Direct cost of ER-level exacerbation
Probability of death from severe exacerbation (ER visit)
Direct cost of hospitalization
Probability of death from severe exacerbation (SCS)
Probability of hospitalization from well-controlled asthma
Initial age for the adult cohort
PM2.5 exposure scenario (e.g., projection levels)
Proportion of outdoor PM2.5 that infiltrates indoors
Proportion of time spent at home, stratified by age
Effectiveness of HEPA air cleaner in reducing PM2.5 indoors
Concentration-response function (CRF) for asthma incidence
Relative risk for asthma control
Relative risk for moderate exacerbation (oral SCS)
Relative risk for severe exacerbation (ER visit)
Relative risk for severe exacerbation (hospitalization)
Relative risk of exacerbation for not well-controlled asthma
Probability of transitioning from well-controlled asthma
Probability of transition from well-controlled to not well-controlled asthma
Probability of moderate exacerbation from well-controlled asthma
Probability of ER visit from well-controlled asthma
Probability of improving from not well-controlled to well-controlled asthma
Cost of HEPA air cleaner (adjusted for rebate/discount)
50000
100000
150000
200000
250000
ICER (CAD per QALY)

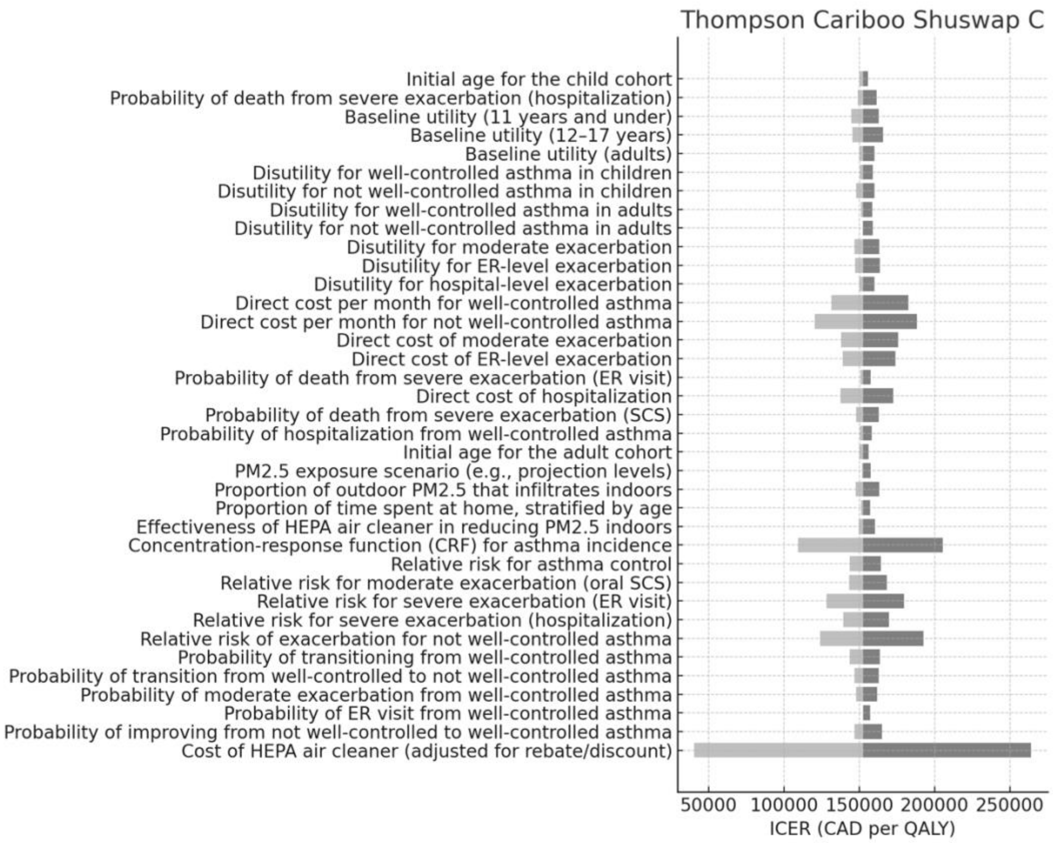
Thompson Cariboo Shuswap C
Initial age for the child cohort
Probability of death from severe exacerbation (hospitalization)
Baseline utility (11 years and under)
Baseline utility (12–17 years)
Baseline utility (adults)
Disutility for well-controlled asthma in children
Disutility for not well-controlled asthma in children
Disutility for well-controlled asthma in adults
Disutility for not well-controlled asthma in adults
Disutility for moderate exacerbation
Disutility for ER-level exacerbation
Disutility for hospital-level exacerbation
Direct cost per month for well-controlled asthma
Direct cost per month for not well-controlled asthma
Direct cost of moderate exacerbation
Direct cost of ER-level exacerbation
Probability of death from severe exacerbation (ER visit)
Direct cost of hospitalization
Probability of death from severe exacerbation (SCS)
Probability of hospitalization from well-controlled asthma
Initial age for the adult cohort
PM2.5 exposure scenario (e.g., projection levels)
Proportion of outdoor PM2.5 that infiltrates indoors
Proportion of time spent at home, stratified by age
Effectiveness of HEPA air cleaner in reducing PM2.5 indoors
Concentration-response function (CRF) for asthma incidence
Relative risk for asthma control
Relative risk for moderate exacerbation (oral SCS)
Relative risk for severe exacerbation (ER visit)
Relative risk for severe exacerbation (hospitalization)
Relative risk of exacerbation for not well-controlled asthma
Probability of transitioning from well-controlled asthma
Probability of transition from well-controlled to not well-controlled asthma
Probability of moderate exacerbation from well-controlled asthma
Probability of ER visit from well-controlled asthma
Probability of improving from not well-controlled to well-controlled asthma
Cost of HEPA air cleaner (adjusted for rebate/discount)
50000 100000 150000 200000 250000
ICER (CAD per QALY)

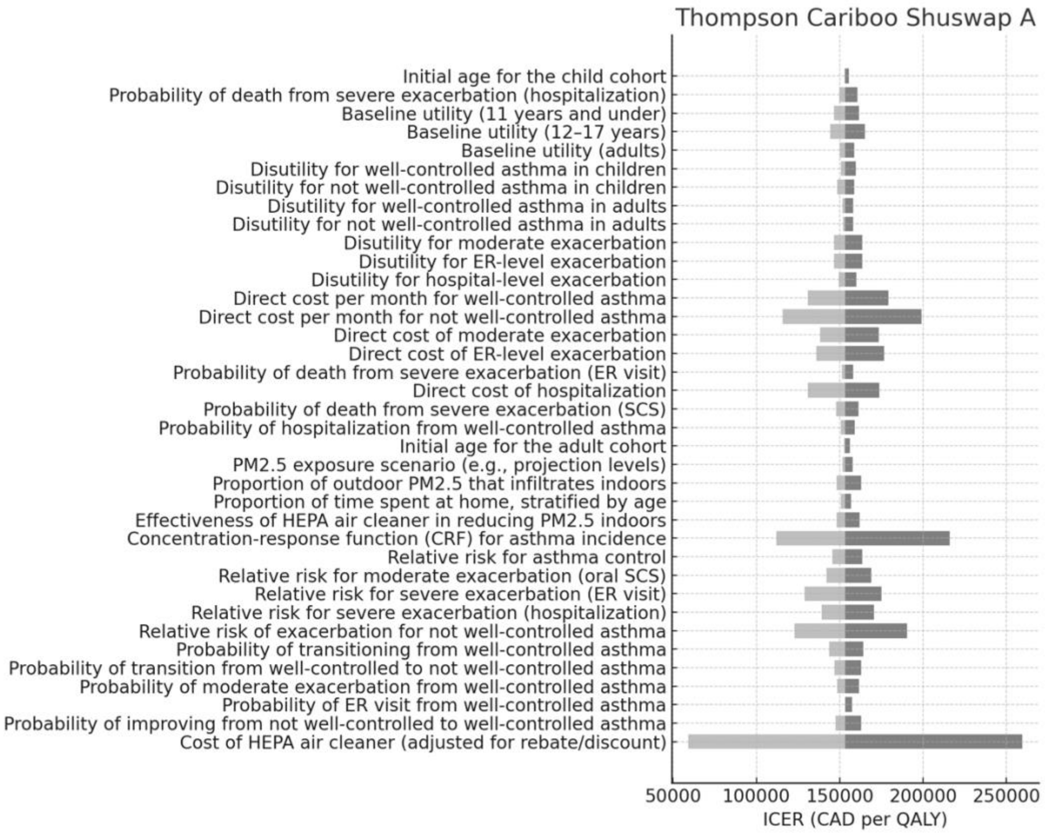
Thompson Cariboo Shuswap A
Initial age for the child cohort
Probability of death from severe exacerbation (hospitalization)
Baseline utility (11 years and under)
Baseline utility (12–17 years)
Baseline utility (adults)
Disutility for well-controlled asthma in children
Disutility for not well-controlled asthma in children
Disutility for well-controlled asthma in adults
Disutility for not well-controlled asthma in adults
Disutility for moderate exacerbation
Disutility for ER-level exacerbation
Disutility for hospital-level exacerbation
Direct cost per month for well-controlled asthma
Direct cost per month for not well-controlled asthma
Direct cost of moderate exacerbation
Direct cost of ER-level exacerbation
Probability of death from severe exacerbation (ER visit)
Direct cost of hospitalization
Probability of death from severe exacerbation (SCS)
Probability of hospitalization from well-controlled asthma
Initial age for the adult cohort
PM2.5 exposure scenario (e.g., projection levels)
Proportion of outdoor PM2.5 that infiltrates indoors
Proportion of time spent at home, stratified by age
Effectiveness of HEPA air cleaner in reducing PM2.5 indoors
Concentration-response function (CRF) for asthma incidence
Relative risk for asthma control
Relative risk for moderate exacerbation (oral SCS)
Relative risk for severe exacerbation (ER visit)
Relative risk for severe exacerbation (hospitalization)
Relative risk of exacerbation for not well-controlled asthma
Probability of transitioning from well-controlled asthma
Probability of transition from well-controlled to not well-controlled asthma
Probability of moderate exacerbation from well-controlled asthma
Probability of ER visit from well-controlled asthma
Probability of improving from not well-controlled to well-controlled asthma
Cost of HEPA air cleaner (adjusted for rebate/discount)
50000 100000 150000 200000 250000
ICER (CAD per QALY)

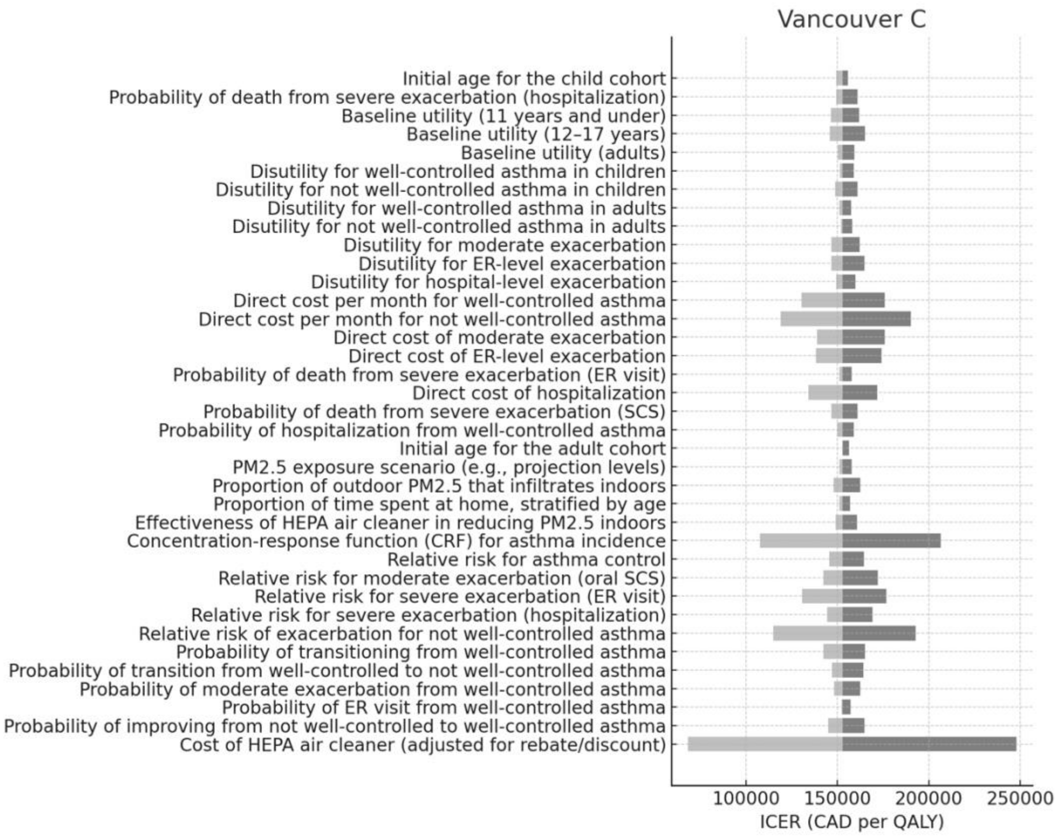
Vancouver C
Initial age for the child cohort
Probability of death from severe exacerbation (hospitalization)
Baseline utility (11 years and under)
Baseline utility (12–17 years)
Baseline utility (adults)
Disutility for well-controlled asthma in children
Disutility for not well-controlled asthma in children
Disutility for well-controlled asthma in adults
Disutility for not well-controlled asthma in adults
Disutility for moderate exacerbation
Disutility for ER-level exacerbation
Disutility for hospital-level exacerbation
Direct cost per month for well-controlled asthma
Direct cost per month for not well-controlled asthma
Direct cost of moderate exacerbation
Direct cost of ER-level exacerbation
Probability of death from severe exacerbation (ER visit)
Direct cost of hospitalization
Probability of death from severe exacerbation (SCS)
Probability of hospitalization from well-controlled asthma
Initial age for the adult cohort
PM2.5 exposure scenario (e.g., projection levels)
Proportion of outdoor PM2.5 that infiltrates indoors
Proportion of time spent at home, stratified by age
Effectiveness of HEPA air cleaner in reducing PM2.5 indoors
Concentration-response function (CRF) for asthma incidence
Relative risk for asthma control
Relative risk for moderate exacerbation (oral SCS)
Relative risk for severe exacerbation (ER visit)
Relative risk for severe exacerbation (hospitalization)
Relative risk of exacerbation for not well-controlled asthma
Probability of transitioning from well-controlled asthma
Probability of transition from well-controlled to not well-controlled asthma
Probability of moderate exacerbation from well-controlled asthma
Probability of ER visit from well-controlled asthma
Probability of improving from not well-controlled to well-controlled asthma
Cost of HEPA air cleaner (adjusted for rebate/discount)
100000
150000
200000
250000
ICER (CAD per QALY)

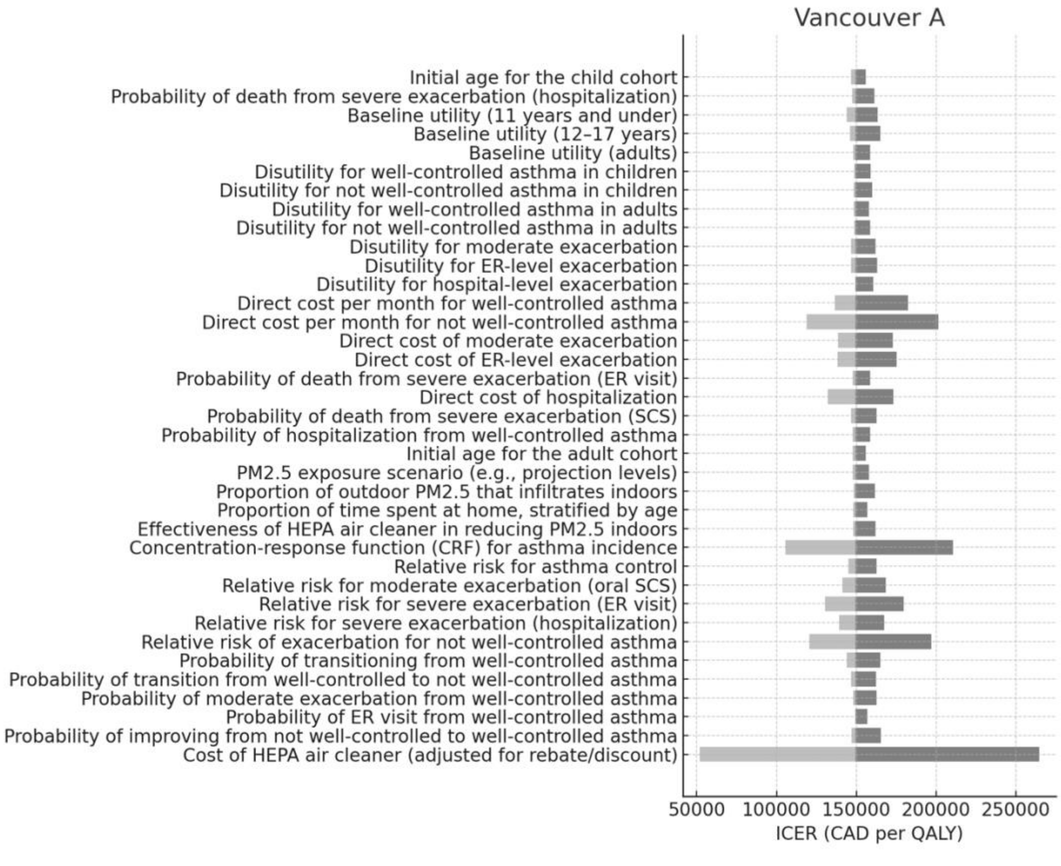
Vancouver A
Initial age for the child cohort
Probability of death from severe exacerbation (hospitalization)
Baseline utility (11 years and under)
Baseline utility (12–17 years)
Baseline utility (adults)
Disutility for well-controlled asthma in children
Disutility for not well-controlled asthma in children
Disutility for well-controlled asthma in adults
Disutility for not well-controlled asthma in adults
Disutility for moderate exacerbation
Disutility for ER-level exacerbation
Disutility for hospital-level exacerbation
Direct cost per month for well-controlled asthma
Direct cost per month for not well-controlled asthma
Direct cost of moderate exacerbation
Direct cost of ER-level exacerbation
Probability of death from severe exacerbation (ER visit)
Direct cost of hospitalization
Probability of death from severe exacerbation (SCS)
Probability of hospitalization from well-controlled asthma
Initial age for the adult cohort
PM2.5 exposure scenario (e.g., projection levels)
Proportion of outdoor PM2.5 that infiltrates indoors
Proportion of time spent at home, stratified by age
Effectiveness of HEPA air cleaner in reducing PM2.5 indoors
Concentration-response function (CRF) for asthma incidence
Relative risk for asthma control
Relative risk for moderate exacerbation (oral SCS)
Relative risk for severe exacerbation (ER visit)
Relative risk for severe exacerbation (hospitalization)
Relative risk of exacerbation for not well-controlled asthma
Probability of transitioning from well-controlled asthma
Probability of transition from well-controlled to not well-controlled asthma
Probability of moderate exacerbation from well-controlled asthma
Probability of ER visit from well-controlled asthma
Probability of improving from not well-controlled to well-controlled asthma
Cost of HEPA air cleaner (adjusted for rebate/discount)
50000
100000
150000
200000
250000
ICER (CAD per QALY)

**Appendix 8.** Probability of cost-effectiveness in probabilistic sensitivity analysis (PSA) with $50,000/Quality adjusted life-year (QALY), $100,000/QALY, and $150,000/QALY willingness-to-pay thresholds.

| | **ICER threshold and cohort** | | | | | |
|---|---|---|---|---|---|---|
| | **$50,000/QALY** | | **$100,000/QALY** | | **$150,000/QALY** | |
| **HSDA** | **Age 5** | **Age 25** | **Age 5** | **Age 25** | **Age 5** | **Age 25** |
| Central Vancouver Island | 0.022% | 0.023% | 3.32% | 3.42% | 53.31% | 54.54% |
| East Kootenay | 0.024% | 0.025% | 3.60% | 3.69% | 57.63% | 59.42% |
| Fraser East | 0.024% | 0.025% | 3.60% | 3.69% | 57.54% | 58.84% |
| Fraser North | 0.024% | 0.025% | 3.60% | 3.69% | 58.30% | 59.27% |
| Fraser South | 0.023% | 0.023% | 3.42% | 3.51% | 54.83% | 55.94% |
| Kootenay Boundary | 0.023% | 0.024% | 3.47% | 3.56% | 55.63% | 56.52% |
| North Shore/Coast Garibaldi | 0.024% | 0.025% | 3.66% | 3.74% | 58.20% | 59.65% |
| North Vancouver Island | 0.024% | 0.029% | 3.60% | 4.30% | 57.67% | 69.00% |
| Northeast | 0.024% | 0.025% | 3.59% | 3.68% | 57.83% | 58.54% |
| Northern Interior | 0.022% | 0.023% | 3.34% | 3.42% | 53.01% | 54.34% |
| Northwest | 0.027% | 0.027% | 4.02% | 4.12% | 64.24% | 65.48% |
| Okanagan | 0.028% | 0.026% | 4.14% | 3.87% | 66.00% | 62.20% |
| Richmond | 0.027% | 0.027% | 3.98% | 4.08% | 63.48% | 64.72% |
| South Vancouver Island | 0.028% | 0.028% | 4.16% | 4.26% | 66.47% | 68.23% |
| Thompson Cariboo Shuswap | 0.023% | 0.023% | 3.41% | 3.49% | 54.65% | 55.58% |
| Vancouver | 0.025% | 0.026% | 3.82% | 3.91% | 60.78% | 62.58% |

**Appendix 9.** Scenario analysis from societal perspective.

| ` | Age 5 (to 30) cohort | | | Age 25 (to 50) cohort | | |
|---|---|---|---|---|---|---|
| **HSDA** | **Δ Cost** | **Δ QALY** | **ICER** | **Δ Cost** | **Δ QALY** | **ICER** |
| Central Vancouver Island | 361.91 | 0.00060 | 427,756 | 308.58 | 0.00074 | 550,956 |
| East Kootenay | 99.41 | 0.02145 | 397,879 | 125.26 | 0.00025 | 479,193 |
| Fraser East | 399.20 | 0.00910 | 468,543 | 479.73 | 0.00104 | 436,981 |
| Fraser North | 771.78 | 0.01615 | 341,403 | 557.21 | 0.00265 | 410,813 |
| Fraser South | 1,177.89 | 0.02257 | 491,390 | 1,226.03 | 0.00275 | 504,658 |
| Kootenay Boundary | 103.08 | 0.00203 | 491,078 | 86.26 | 0.00019 | 540,445 |
| North Shore/Coast Garibaldi | 287.21 | 0.00703 | 539,737 | 370.15 | 0.00093 | 463,314 |
| North Vancouver Island | 143.58 | 0.00312 | 508,727 | 156.38 | 0.00032 | 520,746 |
| Northeast | 94.55 | 0.00245 | 409,876 | 182.85 | 0.00033 | 487,970 |
| Northern Interior | 167.42 | 0.00447 | 476,253 | 279.22 | 0.00051 | 432,977 |
| Northwest | 137.03 | 0.00244 | 344,772 | 118.76 | 0.00022 | 525,848 |
| Okanagan | 411.10 | 0.00805 | 499,667 | 451.99 | 0.00115 | 513,871 |
| Richmond | 229.78 | 0.00509 | 488,067 | 349.55 | 0.00083 | 440,469 |
| South Vancouver Island | 451.95 | 0.00834 | 462,409 | 574.12 | 0.00151 | 421,735 |
| Thompson Cariboo Shuswap | 259.75 | 0.00576 | 313,391 | 384.20 | 0.00073 | 264,255 |
| Vancouver | 505.78 | 0.01298 | 476,240 | 1,919.72 | 0.00369 | 425,300 |
| **BC overall** | 410.39 | 0.01157 | 445,907 | 958.80 | 0.00202 | 418,306 |

**Appendix 10.** Scenario analysis at 0% discounting rate.

| ` | Age 5 (to 30) cohort | | | Age 25 (to 50) cohort | | |
|---|---|---|---|---|---|---|
| **HSDA** | **Δ Cost** | **Δ QALY** | **ICER** | **Δ Cost** | **Δ QALY** | **ICER** |
| Central Vancouver Island | 89.35 | 0.00060 | 147,951 | 110.52 | 0.00074 | 148,853 |
| East Kootenay | 30.91 | 0.02145 | 144,094 | 35.11 | 0.00025 | 141,867 |
| Fraser East | 136.00 | 0.00910 | 149,484 | 152.77 | 0.00104 | 146,964 |
| Fraser North | 235.98 | 0.01615 | 146,146 | 383.28 | 0.00265 | 144,459 |
| Fraser South | 335.84 | 0.02257 | 148,788 | 402.47 | 0.00275 | 146,235 |
| Kootenay Boundary | 29.60 | 0.00203 | 145,859 | 26.95 | 0.00019 | 143,283 |
| North Shore/Coast Garibaldi | 102.09 | 0.00703 | 145,237 | 134.55 | 0.00093 | 144,583 |
| North Vancouver Island | 46.12 | 0.00312 | 147,900 | 46.50 | 0.00032 | 146,788 |
| Northeast | 35.34 | 0.00245 | 144,522 | 47.70 | 0.00033 | 146,004 |
| Northern Interior | 66.12 | 0.00447 | 147,761 | 75.91 | 0.00051 | 147,464 |
| Northwest | 35.60 | 0.00244 | 146,163 | 31.90 | 0.00022 | 146,469 |
| Okanagan | 117.93 | 0.00805 | 146,524 | 168.35 | 0.00115 | 146,595 |
| Richmond | 73.88 | 0.00509 | 145,190 | 118.47 | 0.00083 | 142,454 |
| South Vancouver Island | 122.12 | 0.00834 | 146,504 | 222.12 | 0.00151 | 146,643 |
| Thompson Cariboo Shuswap | 84.39 | 0.00576 | 146,462 | 107.19 | 0.00073 | 146,312 |
| Vancouver | 190.28 | 0.01298 | 146,604 | 528.84 | 0.00369 | 143,211 |
| **BC overall** | 169.73 | 0.01157 | 146,656 | 292.52 | 0.00202 | 144,843 |

**Appendix 11.** Scenario analysis at 3% discounting rate.

| ` | Age 5 (to 30) cohort | | | Age 25 (to 50) cohort | | |
|---|---|---|---|---|---|---|
| **HSDA** | **Δ Cost** | **Δ QALY** | **ICER** | **Δ Cost** | **Δ QALY** | **ICER** |
| Central Vancouver Island | $98.75 | 0.00062 | 160,286 | $122.16 | 0.00076 | 161,263 |
| East Kootenay | $34.17 | 0.02189 | 156,109 | $38.81 | 0.00025 | 153,695 |
| Fraser East | $150.32 | 0.00928 | 161,947 | $168.85 | 0.00106 | 159,218 |
| Fraser North | $260.82 | 0.01647 | 158,331 | $423.62 | 0.00271 | 156,503 |
| Fraser South | $371.20 | 0.02303 | 161,193 | $444.83 | 0.00281 | 158,427 |
| Kootenay Boundary | $32.72 | 0.00207 | 158,020 | $29.79 | 0.00019 | 155,229 |
| North Shore/Coast Garibaldi | $112.83 | 0.00717 | 157,346 | $148.71 | 0.00095 | 156,637 |
| North Vancouver Island | $50.98 | 0.00318 | 160,231 | $51.40 | 0.00032 | 159,027 |
| Northeast | $39.06 | 0.00249 | 156,572 | $52.72 | 0.00033 | 158,177 |
| Northern Interior | $73.08 | 0.00457 | 160,081 | $83.91 | 0.00053 | 159,759 |
| Northwest | $39.34 | 0.00248 | 158,349 | $35.26 | 0.00022 | 158,681 |
| Okanagan | $130.35 | 0.00821 | 158,741 | $186.07 | 0.00117 | 158,817 |
| Richmond | $81.66 | 0.00519 | 157,296 | $130.94 | 0.00085 | 154,332 |
| South Vancouver Island | $134.98 | 0.00850 | 158,719 | $245.50 | 0.00155 | 158,869 |
| Thompson Cariboo Shuswap | $93.27 | 0.00588 | 158,674 | $118.47 | 0.00075 | 158,512 |
| Vancouver | $210.30 | 0.01324 | 158,827 | $584.50 | 0.00377 | 155,152 |
| **BC overall** | $187.59 | 0.01181 | 158,884 | $323.32 | 0.00206 | 156,919 |

**Appendix 12.** Scenario analysis varying rebate levels. A) $50 air cleaner rebate; B) $100 rebate; C) $200 rebate.

| | Age 5 (to 30) cohort | | | Age 25 (to 50) cohort | | |
|---|---|---|---|---|---|---|
| **HSDA** | **Δ Cost** | **Δ QALY** | **ICER** | **Δ Cost** | **Δ QALY** | **ICER** |
| Central Vancouver Island | 33.09 | 0.00061 | 32,821 | 35.80 | 0.0008 | 31,358 |
| East Kootenay | 9.09 | 0.02167 | 30,920 | 14.53 | 0.0003 | 32,318 |
| Fraser East | 36.50 | 0.00919 | 29,817 | 55.65 | 0.0012 | 29,471 |
| Fraser North | 70.56 | 0.01631 | 32,926 | 64.64 | 0.0030 | 33,506 |
| Fraser South | 107.69 | 0.0228 | 31,270 | 142.22 | 0.0031 | 34,035 |
| Kootenay Boundary | 9.42 | 0.00205 | 25,650 | 10.01 | 0.0002 | 24,848 |
| North Shore/Coast Garibaldi | 26.26 | 0.0071 | 34,347 | 42.94 | 0.0011 | 31,247 |
| North Vancouver Island | 13.13 | 0.00315 | 32,374 | 18.14 | 0.0004 | 35,120 |
| Northeast | 8.64 | 0.00247 | 27,763 | 21.21 | 0.0004 | 27,110 |
| Northern Interior | 15.31 | 0.00452 | 30,307 | 32.39 | 0.0006 | 29,201 |
| Northwest | 12.53 | 0.00246 | 33,140 | 13.78 | 0.0002 | 23,864 |
| Okanagan | 37.59 | 0.00813 | 31,797 | 52.43 | 0.0013 | 34,656 |
| Richmond | 21.01 | 0.00514 | 31,059 | 40.55 | 0.0009 | 29,706 |
| South Vancouver Island | 41.32 | 0.00842 | 29,426 | 66.60 | 0.0017 | 28,443 |
| Thompson Cariboo Shuswap | 23.75 | 0.00582 | 25,543 | 44.57 | 0.0008 | 28,262 |
| Vancouver | 46.24 | 0.01311 | 30,306 | 222.69 | 0.0042 | 28,683 |
| **BC overall** | **37.52** | **0.01169** | **31,176** | **111.22** | **0.0023** | **33,431** |

B) $100 rebate

| ` | **Age 5 (to 30) cohort** | | | **Age 25 (to 50) cohort** | | |
|---|---|---|---|---|---|---|
| **HSDA** | **Δ Cost** | **Δ QALY** | **ICER** | **Δ Cost** | **Δ QALY** | **ICER** |

| | | | | | | |
|---|---|---|---|---|---|---|
| Central Vancouver Island | 98.44 | 0.00061 | 55,679 | 97.92 | 0.0008 | 51,362 |
| East Kootenay | 27.04 | 0.02167 | 52,453 | 13.83 | 0.0003 | 52,934 |
| Fraser East | 108.59 | 0.00919 | 50,582 | 52.98 | 0.0012 | 48,271 |
| Fraser North | 209.93 | 0.01631 | 55,856 | 61.53 | 0.0030 | 54,881 |
| Fraser South | 320.39 | 0.0228 | 53,048 | 135.39 | 0.0031 | 55,747 |
| Kootenay Boundary | 28.04 | 0.00205 | 43,514 | 9.52 | 0.0002 | 40,700 |
| North Shore/Coast Garibaldi | 78.12 | 0.0071 | 58,267 | 40.88 | 0.0011 | 51,180 |
| North Vancouver Island | 39.05 | 0.00315 | 54,920 | 17.27 | 0.0004 | 57,524 |
| Northeast | 25.71 | 0.00247 | 47,098 | 20.19 | 0.0004 | 44,404 |
| Northern Interior | 45.53 | 0.00452 | 51,414 | 30.83 | 0.0006 | 47,829 |
| Northwest | 37.27 | 0.00246 | 56,220 | 13.12 | 0.0002 | 59,088 |
| Okanagan | 111.82 | 0.00813 | 53,941 | 49.91 | 0.0013 | 56,764 |
| Richmond | 62.50 | 0.00514 | 52,689 | 38.60 | 0.0009 | 52,656 |
| South Vancouver Island | 122.93 | 0.00842 | 51,919 | 63.40 | 0.0017 | 56,587 |
| Thompson Cariboo Shuswap | 70.65 | 0.00582 | 43,332 | 42.43 | 0.0008 | 46,291 |
| Vancouver | 137.57 | 0.01311 | 51,412 | 212.00 | 0.0042 | 56,980 |
| **BC overall** | **111.62** | **0.01169** | **52,887** | **105.88** | **0.0023** | **54,758** |

$200 rebate

| ` | **Age 5 (to 30) cohort** | | | **Age 25 (to 50) cohort** | | |
|---|---|---|---|---|---|---|
| **HSDA** | **Δ Cost** | **Δ QALY** | **ICER** | **Δ Cost** | **Δ QALY** | **ICER** |
| Central Vancouver Island | 423.94 | 0.00061 | 263,741 | 430.98 | 0.0008 | 232,204 |
| East Kootenay | 116.44 | 0.02167 | 248,463 | 174.93 | 0.0003 | 236,251 |
| Fraser East | 467.65 | 0.00919 | 239,598 | 670.02 | 0.0012 | 224,250 |
| Fraser North | 904.09 | 0.01631 | 264,582 | 778.25 | 0.0030 | 241,261 |
| Fraser South | 1379.81 | 0.0228 | 251,280 | 1712.37 | 0.0031 | 243,491 |
| Kootenay Boundary | 120.75 | 0.00205 | 206,118 | 120.46 | 0.0002 | 204,761 |
| North Shore/Coast Garibaldi | 336.45 | 0.0071 | 276,003 | 516.96 | 0.0011 | 231,737 |
| North Vancouver Island | 168.18 | 0.00315 | 260,145 | 218.42 | 0.0004 | 248,067 |
| Northeast | 110.74 | 0.00247 | 223,097 | 255.38 | 0.0004 | 214,295 |
| Northern Interior | 196.10 | 0.00452 | 243,540 | 389.96 | 0.0006 | 223,110 |
| Northwest | 160.52 | 0.00246 | 266,306 | 165.88 | 0.0002 | 200,611 |
| Okanagan | 481.59 | 0.00813 | 255,510 | 631.27 | 0.0013 | 246,110 |
| Richmond | 269.17 | 0.00514 | 249,579 | 488.22 | 0.0009 | 225,240 |
| South Vancouver Island | 529.43 | 0.00842 | 236,457 | 801.84 | 0.0017 | 219,914 |
| Thompson Cariboo Shuswap | 304.26 | 0.00582 | 205,259 | 536.61 | 0.0008 | 219,151 |
| Vancouver | 592.49 | 0.01311 | 243,531 | 2681.22 | 0.0042 | 220,926 |
| **BC overall** | **480.73** | **0.01169** | **250,520** | **1339.12** | **0.0023** | **240,946** |

**Appendix 13.** Scenario analysis of alternative infiltration efficiency of 0.28.

| | Age 5-30 cohort | | | Age 25-50 cohort | | |
|---|---|---|---|---|---|---|
| **HSDA** | **Δ Cost** | **Δ QALY** | **ICER** | **Δ Cost** | **Δ QALY** | **ICER** |
| Central Vancouver Island | 296.76 | 0.00049 | $184,618 | 344.79 | 0.00076 | $158,645 |
| East Kootenay | 46.58 | 0.01734 | $173,924 | 139.95 | 0.00025 | $163,501 |
| Fraser East | 187.06 | 0.00735 | $167,719 | 536.02 | 0.00106 | $149,099 |
| Fraser North | 361.64 | 0.01305 | $185,207 | 622.60 | 0.00270 | $169,514 |
| Fraser South | 551.93 | 0.01824 | $175,896 | 1369.90 | 0.00280 | $172,190 |
| Kootenay Boundary | 48.30 | 0.00164 | $144,283 | 96.37 | 0.00019 | $125,713 |
| North Shore/Coast Garibaldi | 134.58 | 0.00568 | $193,202 | 413.57 | 0.00095 | $158,084 |
| North Vancouver Island | 67.27 | 0.00252 | $182,102 | 174.74 | 0.00032 | $177,680 |
| Northeast | 44.30 | 0.00198 | $156,168 | 204.30 | 0.00033 | $137,154 |
| Northern Interior | 78.44 | 0.00362 | $170,478 | 311.97 | 0.00052 | $147,732 |
| Northwest | 64.21 | 0.00197 | $186,414 | 132.71 | 0.00022 | $120,733 |
| Okanagan | 192.63 | 0.00650 | $178,857 | 505.01 | 0.00117 | $175,332 |
| Richmond | 107.67 | 0.00411 | $174,705 | 390.58 | 0.00085 | $150,288 |
| South Vancouver Island | 211.77 | 0.00674 | $165,520 | 641.47 | 0.00154 | $143,897 |
| Thompson Cariboo Shuswap | 121.70 | 0.00466 | $143,681 | 429.29 | 0.00075 | $142,981 |
| Vancouver | 237.00 | 0.01049 | $170,472 | 2144.98 | 0.00376 | $145,112 |
| **BC overall** | **192.29** | **0.00935** | **$175,364** | **1071.30** | **0.00206** | **$169,135** |
